# Myocarditis and Pericarditis following COVID-19 Vaccination: Rapid Systematic Review of Incidence, Risk Factors, and Clinical Course

**DOI:** 10.1101/2021.11.19.21266605

**Authors:** Jennifer Pillay, Liza Bialy, Lindsay Gaudet, Aireen Wingert, Andrew S. Mackie, D. Ian Paterson, Lisa Hartling

## Abstract

**Objectives:** Myocarditis and pericarditis are adverse events of special interest after vaccination with mRNA vaccines. This rapid systematic review examined incidence rates of myocarditis and pericarditis after COVID-19 vaccination, and the presentation and clinical course of cases.

**Design:** Rapid systematic review

**Data sources:** Medline, Embase and the Cochrane Library were searched from October 2020 to October 6, 2021; reference lists and grey literature (to Oct 21, 2021).

**Review methods:** Randomized controlled trials (RCTs) and large population-based/multisite observational studies and surveillance data reporting on myocarditis or pericarditis in people of any age after receiving any COVID-19 vaccine; systematic reviews of case series. A single reviewer completed screening and another verified 50% of exclusions, using a machine-learning program to prioritize records. A second reviewer verified all exclusions at full text, data extractions, and (for incidence) risk of bias assessments using Cochrane Risk of Bias 2.0 and Joanna Briggs Institute tools. Certainty of evidence ratings for incidence were based on team consensus using GRADE. Patient partners provided key messages from their interpretations of the findings.

**Results:** 3457 titles/abstracts and 159 full texts were screened. For incidence rates we included 7 RCTs (n=3732 to 44,325) and 22 large observational studies/data sources using passive (n=10) and active (n=12) surveillance; for case presentation, we included 11 case series published as articles and three based on publicly available websites (n=12,636 cases). Mainly due to imprecision, the RCTs provided very low certainty evidence for incidence of myocarditis or pericarditis. From observational data, the incidence of myocarditis following mRNA vaccines is low but probably highest in males 12-17 years (55 [7-day risk] to 134 [30-day risk] cases per million; specific to Pfizer) and 18-29 years (40 [7-day risk] to 99 [21-30 day risk]) cases per million) (Moderate certainty evidence). Incidence is lower (<20 per million) or little-to-none in older ages and across all ages of females (Low certainty). Evidence for pericarditis was of very low certainty. Among adult males under 40 years, Moderna compared with Pfizer vaccine may be associated with a small increase (<20 per million) in risk for myocarditis or (one of) myocarditis or pericarditis following vaccination (Low certainty); the evidence for youth under 18 years was very uncertain. No study examined differences in incidence based on pre-existing condition(s) or risk factors apart from age and sex. The majority of myocarditis cases involved males (often >90%) in their 20s, with a short symptom onset of 2 to 4 days after a second dose (71-100%). The majority of cases presented with chest pain/pressure and troponin elevation; a minority (<30%) had left ventricular dysfunction. Most were hospitalized (≥84%), without stays in intensive care units, for a short duration (2-4 d) and treated with anti-inflammatory and/or other supportive therapies. Almost all reports of death are from unverified cases and of unclear cause. Most cases of pericarditis were unconfirmed; for this outcome there appears to be more variation in age, sex, onset timing and rate of hospitalization.

**Conclusions:** Incidence of myocarditis following mRNA vaccines is low but probably highest in males 12-29 years old. Existing evidence does not strongly support preference of one mRNA vaccine, even in young males. Continued active surveillance of myocarditis incidence out to 30 days from dosing is recommended with respect to i) new populations (i.e., children <12y), ii) third and subsequent doses, and iii) affected individuals receiving subsequent mRNA vaccine doses. Future research is needed to examine other risk factors and long-term effects.

**Funding and Registration no:** This project was funded in part by the Canadian Institutes of Health Research (CIHR) through the COVID-19 Evidence Network to support Decision-making (COVID-END) at McMaster University. Not registered.

Summary box

What is already known about this topic?
Case reports and surveillance signals of myocarditis (inflammation of the heart muscle) and pericarditis (inflammation of the two-layered sac surrounding the heart) after COVID-19 vaccination appeared as early as April 2021.
These have prompted ongoing surveillance and research of these complications to investigate their incidence, possible attribution to the vaccines, and clinical course.

What this study adds
This review critically appraises and synthesizes the available evidence to-date on the incidence of myocarditis and pericarditis after COVID-19 vaccination in multiple countries. It summarizes the presentation and clinical course of over 12,000 reported cases.
Though low, the incidence in young males 12-29 years of age is probably the highest and appears to be similar across mRNA vaccines. Most cases present with chest pain and are mild and self-limiting. Continued active surveillance is warranted especially with vaccine rollout to young children and use of third doses, and to learn of any long-term consequences.

## Introduction

Case reports and surveillance signals^1-4^ of myocarditis (including myopericarditis) and pericarditis after COVID-19 vaccination appeared as early as April 2021. This led to the surveillance of adverse events of special interest following vaccination with messenger RNA (mRNA) vaccines manufactured by Pfizer/BioNTech and Moderna. Estimated rates of myocarditis in the United Kingdom (UK)^5^ and the United States (US)^6^ are 11 and 1.3 cases per 100,000 person-years, respectively, regardless of age. Estimated background/expected rates of myocarditis and pericarditis following COVID-19 vaccination in the US, adjusted for a 7-day risk period where most cases appear, are 0.2 and 1.4 per 1 million people, respectively.^6^ Historically, myocarditis has been more prevalent in males than females, from childhood through young adulthood;^7^ this trend has been evident in early case series of post-COVID-19 vaccinations.^8 9^

Symptoms indicative of myocarditis or pericarditis typically include new onset and persisting chest pain, shortness of breath, and/or palpitations.^10^ Diagnosis of a probable case of myocarditis usually requires elevated troponin levels and/or findings from imaging (e.g., echocardiography, magnetic resonance imaging) or other testing (e.g., ECG); histopathology to provide definitive diagnosis is not usually performed.^11^ Differential diagnoses, including COVID-19 infection and community-acquired myocarditis, should be considered and ruled out.^11 12^ Most people experiencing these conditions will fully recover, however, in rare cases, myocarditis can lead to heart failure or asymptomatic left ventricular dysfunction. Long-term consequences associated with pericarditis include one or more recurrences and, rarely, thickening of the pericardium and constrictive heart filling.^13^

This rapid systematic review aimed to provide estimates of the incidence rates for these harms after vaccination for COVID -19, including whether these vary by patient and/or vaccine characteristics, and to describe the clinical course of myocarditis and pericarditis after COVID-19 vaccination.

### Research Questions

1. What is the incidence of myocarditis and pericarditis following COVID-19 vaccination, and does the incidence vary by patient (e.g., age, sex, race/ethnicity, pre-existing conditions [e.g. cardiac diseases] or infections [e.g., COVID-19]) and vaccine factors (e.g. vaccine type, dose, interval)?
2. What are the characteristics and short-term clinical course in patients with myocarditis and pericarditis after COVID-19 vaccination?

## Methods

We conducted this rapid systematic review following a protocol developed before starting to screen the literature. Due to the rapid development and conduct of this review we did not register the protocol. We followed guidance for systematic reviews when conducting^14^ and reporting^15^ this review.

### Literature Search

We worked with an experienced medical information specialist to develop the search strategy, which was peer-reviewed^16^ (**Appendix 1**). Searches combined concepts for COVID-19, vaccines, and myocarditis/ pericarditis/cardiovascular manifestations/adverse events/surveillance. We limited our search to articles published since October 2020. We did not add limits for language, country, or study design, but had limits for human (not animal only) studies and commentaries. Using the multifile option as well as the deduping tool in Ovid, we searched Ovid MEDLINE(R) and Epub Ahead of Print, In-Process, In-Data-Review & Other Non-Indexed Citations and Daily <1946 to October 05, 2021> and Embase <1974 to 2021 October 05>. We also searched the Cochrane Library and medRxiv (last 2 months). These searches were run on October 6, 2021. We searched grey literature by scanning 20 key national websites (e.g., Public Health Agency of Canada, UK’s Medicines & Healthcare Products Regulatory Agency, Centers for Disease Control and Prevention) to identify unpublished data. We also scanned reference lists of included studies.

### Eligibility Criteria

Our inclusion criteria are outlined in **Table 1**.

**Table 1.** Eligibility Criteria for Each Research Question.

|  |  |
| --- | --- |
| **Population** | 1. People of any age<br>2. People of any age experiencing myocarditis, pericarditis, or myopericarditis after receiving a COVID-19 vaccine |
| **Intervention/**<br>**Exposure** | 1. BNT162b2 mRNA/PfizerBioNTech/Comirnaty, mRNA-1273/Moderna Spikevax, AZD1222 (ChAdOx1 nCoV-19)/AstraZeneca Vaxzevria, Ad26.COV2.S/Janssen (alternative manufacturers of same vaccine are eligible)<br>2. Same as 1. |
| **Control/**<br>**Comparator** | 1. Comparators of previously vaccinated people (no longer at risk for outcome) or unvaccinated people; or no comparator. Risk factors include; age, sex, race/ethnicity, preexisting chronic conditions [e.g. cardiac diseases, immunocompromise] or infections (e.g., recent/past COVID-19), vaccine type (e.g., mRNA vs vector)/dose/interval<br>2. No comparator |
| **Outcomes** | 1. Incidence rate/cumulative risk of myocarditis (or myopericarditis) and/or pericarditis after receipt of vaccine (any dose).<br>2. Characteristics of the patients (e.g., age, sex, pre-existing conditions [e.g. cardiac diseases] and infections [e.g. recent/past COVID-19], race/ethnicity) and case presentation (e.g., timing/dose/type of vaccine, diagnostics, illness severity, treatments provided, short-term outcomes, disposition) |
| **Setting** | Any setting and country |
| **Study designs** | 1. Phase 3/4 RCTs; large (>1000 vaccinated people) population/multisite/health system-based observational studies and surveillance data<br>2. Systematic/rapid reviews and data on cases from studies included in question 1 |
| **Publication**<br>**Language** | 1. English full texts<br>2. English full texts |
| **Publication Date** | 1. Oct 2020-onwards (vaccines were authorized mid-Sept 2020)<br>2. Oct 2020-onwards |

### Study Selection

All reviewers undertaking screening conducted a pilot round in Excel using 300 records. We then conducted screening and selection in DistillerSR (Evidence Partners) using structured forms. For title and abstract review, we applied the machine learning program Daisy AI which continually reprioritizes records during screening.^17^ A single reviewer screened all titles/abstracts and another reviewer verified exclusions for the first 50% of records. For full text selection, a single reviewer reviewed all records, with all exclusions verified by another reviewer. Studies were further verified for inclusion during data extraction.

For research question 1, when reports collected data from similar/overlapping populations (e.g., US national surveillance data), we first included the study with the most recent data and then included additional studies if they differed in their methods (e.g., of verifying cases, differing risk intervals [period after the vaccine where case are counted]), differing age and/or sex stratification, use of unvaccinated or similar controls). For randomized controlled trials (RCTs), we used the report of the largest population as the primary (included) publication and cited associated papers used for data extraction; sub-studies within RCTs, for example of different ages, were considered different studies.

For research question 2, we mapped the case series and case reports contained within the relevant reviews. This served as a baseline for identifying the most recent and comprehensive data source for each geographic region (i.e., US, Canada, UK, Europe, and Israel). When more recent data were reported in studies included in research question 1, these were included in place of studies identified through the reviews. For regions with multiple reports, we prioritized data that was most recent, most comprehensive (largest numbers), and that verified cases. Our goal was to avoid overlap in data, i.e., the same cases reported in different sources. However, we did include sources that reported on specific sub-groups (e.g., 12 to 17 year-olds) to present more details on these groups of interest. We have noted in the evidence tables where there may be overlap in cases between reports.

### Data Extraction

We extracted all data into structured Word forms and conducted a pilot exercise with two studies. Thereafter, one reviewer extracted all data and another verified eligibility and all data. Discrepancies were resolved by discussion or by a review lead.

For research question 1, extracted data related to all elements of the eligibility criteria (Table 1) and data used for risk of bias assessment, focusing on methods for identifying cases (i.e., passive surveillance versus active surveillance/registry data), outcome ascertainment and confirmation/adjudication (including criteria for case definitions and classification), as well as the risk interval for which the events were captured. We distinguished between estimates of incidence compared with an unexposed group (excess incidence/risk differences) versus without a control, and extracted data on incident rates per person-years and per doses of vaccine/people vaccinated (after either dose, dose 1, or dose 2). As available, we extracted data on any stratified or subgroup analyses based on age, sex, pre-existing conditions, race/ethnicity, different vaccine types or doses (events captured after any dose, dose 1, or dose 2), and different risk intervals (0-7d vs. 8-28d vs. longer). Effect measures included: incidence rate/cumulative risk (including excess risk [risk difference] when using a control group) and relative effects between groups (e.g., incidence rate ratio (IRR)), adjusted for key confounders (i.e., age, sex, infection status, cardiac and immunodeficiency/autoimmune conditions) when reported.

For research question 2, we planned to rely on data from the relevant systematic reviews identified through our search. We used the evidence table presented in an existing review^8^ as the foundation for data extraction and presentation of findings as it covered most of the items specified under our outcomes in Table 1. We added the following items: criteria for confirmation of cases, breakdown of cases by diagnosis (myocarditis, pericarditis, myopericarditis), case source, age included, percent with pre-existing conditions, percent admitted to an intensive care unit (ICU), percent hospitalized, number of fatalities. Most of the studies included in the existing reviews were superseded by more recent publications/data; therefore, we extracted data directly from the relevant case series/data sources.

### Risk of Bias Assessment

For research question 1, all reviewers involved in the risk of bias assessments piloted each risk of bias tool with two papers. We used the Cochrane Collaboration ROB 2.0 tool^18^ for RCTs, and the JBI (formally Joanna Briggs Institute) checklist for cohort studies^19^ for observational studies/surveillance data. We focused on valid and reliable case finding and outcome ascertainment methods and, in observational studies, accounting for key confounders including age, sex, existing cardiac diseases/conditions, and prior COVID-19 exposure. We used the findings of the risk of bias assessments when undertaking Grading of Recommendations, Assessment, Development and Evaluation (GRADE) assessments of the certainty of the evidence.^20 21^ Assessments were completed by one reviewer and verified by another. Discrepancies were resolved by discussion or by a review lead.

For research question 2, we did not assess the risk of bias of the included systematic reviews because we did not rely on their reporting or synthesis in any way. We also did not rate the case series data, either published or derived from grey literature or publicly available data sources (e.g., Yellow Card, UK). The intent of question 2 was to characterize these patients and their clinical course without providing quantitative estimates, e.g., of incidence or effectiveness. The main differentiating feature between case series was whether they only reported on cases verified by clinicians; we extracted this information and considered this factor when summarizing the results.

### Data Synthesis

For research question 1, we did not pool results due to heterogeneity in case finding (passive vs. active surveillance), dosing and risk intervals, case ascertainment, populations (age and sex), and the large amount of overlap in some study populations. We tabulated all results and compared and contrasted findings between studies based on the major differentiating population, vaccine and methodologic variables. We reached consensus on a best estimate of the incidence (or a range/upper limit based on different risk intervals or other variables), relying when possible on studies that verified cases and used active surveillance. Based on clinical input we developed primary age categories (12-17y, 18-29y, 30-39y, ≥40y) to rely on when possible. If a study contributed more than one result within these (e.g., 20-24y and 25-29y, results for each mRNA vaccine) we took the average of the incident rates. When a study reported an incidence rate (or data to calculate this) and an IRR compared with a control/background rate, but not the difference in incidence (excess incidence over background rate), we calculated the excess incidence (i.e., crude incidence – [crude incidence/IRR]). We assessed the certainty for each of our conclusion statements using GRADE.^20 21^ RCTs started at high certainty (i.e., indicating we are very confident that the true effect lies close to that of the estimate of the effect) and observational studies started at low certainty (i.e., our confidence in the effect estimate is limited; the true effect may be substantially different from the estimate of the effect). We rated down based on serious concerns about risk of bias, inconsistency, indirectness, imprecision, and/or reporting biases. We did not apply a threshold for an important effect, but in general considered 10-20 or fewer cases per million doses to be less meaningful for decision-making. We downrated for risk of bias when studies using passive surveillance (assuming some under ascertainment) and unverified cases contributed to a synthesis, and for indirectness for comparisons across all ages and both sexes, due to the large heterogeneity in incidence rates across ages (for males) and sexes. We considered uprating for observational studies due to large incidence rates when no other major limitations were evident.

For research question 2, we present the details for each case series in an evidence table and provide a descriptive summary.

### Patient and Public Involvement

Two patient/public partners joined the research team for this review (see Acknowledgements). They were not involved in decisions about the research questions or outcomes, which were determined by the funders. The research leads met with these partners to present and discuss the findings and their implications. The partners co-developed with the leads key messages from their perspective.

## Results

### Study Selection

After deduplication, our database searches retrieved 3439 citations and other sources identified 18 citations (**Appendix 2**). After title/abstract screening, we retrieved and screened at full text 159 citations. For question 1 we included 29 studies reported in 32 publications, and for question 2 we included data for 14 case series (9 were reported in studies included in question 1). **Appendix 3** contains descriptions of the various national surveillance systems from which reports were collated or data collected. Tables of study characteristics, with all data on results, and risk of bias ratings for question 1 are in **Appendix 4**.

### Findings

#### Research Question 1: Incidence of Myocarditis and Pericarditis Following COVID-19 Vaccination

We included seven RCTs (n=3732 to 44,325)^22-28^ reported in 10 papers^29-31^, and 22 large observational studies/data sources (reported in publications, presentations, online reports, accessible data; all are considered here as “studies” for simplicity), with identification of cases based on passive surveillance systems (n=10) ^9 32-40^ or on active surveillance/registry data (n=12).^41-52^ Observational data came from the US, Canada, the UK, Israel, and the European Economic Area (EEA). Several data sources were used in multiple studies (e.g., 5 relied on data from the US VAERS) with different methods (e.g., risk interval used, whether cases were confirmed/verified).

Nine of the 12 studies using active surveillance included a comparator group to estimate excess incidence. Eleven studies examined age and/or sex subgroups and seven provided data by specific vaccine. None assessed differences in incidence based on pre-existing condition(s), or race/ethnicity. Overall, risk of bias for adverse events from the RCTs was low to “some concerns”. Risk of bias was low to “some concerns” for data from active surveillance systems and high for passive surveillance data.

**Tables 2** and **3** include the Summary of Findings tables for evidence from the observational data, including the GRADE certainty assessments. We refer to studies by their source of adverse event data and the date of data collection; references for each studies using this format are in **Appendix 3. Appendix 4** includes the Summary of Findings table for evidence from the RCTs, from which there was low certainty evidence mainly due to very serious concerns about imprecision from inadequate sample sizes for these outcomes. Two observational studies using one cohort of nursing home residents (n∼21,000) reported no events^42 43^

**Table 2:** Summary of Findings for Incident Rates after Receipt of Either mRNA Vaccine. <u>Note</u>: Shading represents the data used to create lower and upper limits of incidence rates, when used in conclusions. See Appendix 3 for references.

| Sex | Age | Studies (data source and date) | Risk interval; Confirmed cases (Y/N) | Incidence rates per million doses after dose 2 of either mRNA vaccine unless otherwise stated<br>Ranges represent each vaccine and square brackets indicate either an average across vaccines/ages or our calculation of excess incidence* | Conclusions | Certainty about conclusions using GRADE |
| --- | --- | --- | --- | --- | --- | --- |
| **Myocarditis** |  |  |  |  |  |  |
| Both sexes | All ages (≥12y unless otherwise stated) | VAERS Aug 6 | None; N | 3.65-6.47 [5.1] | Across all ages, we are very uncertain about the incidence of myocarditis after vaccination with mRNA vaccines | Very low<sup>a</sup> |
|  |  | Yellow Card Oct 13 | NR; N | 8-32 [20] |  |  |
|  |  | Israel MOH May 30 | 30 d; N | 24 (Pfizer) (≥16y) |  |  |
|  |  | EudraVigilance Oct 19 | NR; N | 8.5-25.8 (either dose) [7.2] |  |  |
|  |  | Israel MOH May 31 | 30 d; N | 22.8 (Pfizer) (≥16y) |  |  |
|  |  | Clalit Health May 24a | 42 d after dose 1; Y | 21.3 (Pfizer) (≥16y) |  |  |
|  |  | Clalit Health May 24b | 42 d after dose 1; N | 27 (Pfizer) (≥16y) |  |  |
|  |  | KPSC July 20 | 10 d; Y | 5.8 (>18y) |  |  |
|  |  | Providence Health May 25 | 30d; Y | 10 |  |  |
| Both sexes | <18 y | EudraVigilance Oct 19 | NR; N | 41.6-60.3 (either dose) [50.9] | Among youth of both sexes, we are very uncertain about the incidence of myocarditis after vaccination with the Pfizer vaccine | Very low<sup>b</sup> |
| M | All ages | Israel MOH May 31 | 30 d; N | 38.3 (Pfizer) (≥16y) | Across all ages of males, we are very uncertain about the incidence of myocarditis after vaccination with the Pfizer vaccine | Very low<sup>a</sup> |
|  |  | Clalit Health May 24a | 42 d after dose 1; Y | 41.2 (Pfizer; either dose) (≥16y) |  |  |
| F | All ages | Israel MOH May 31 | 30 d; N | 4.6 (Pfizer) (≥16y) | Across all ages of females, there may be no excess risk of myocarditis after vaccination with the Pfizer vaccine | Low |
|  |  | Clalit Health May 24a | 42 d after dose 1; Y | 2.3 (Pfizer; either dose) (≥16y) |  |  |
| M | 12-17y | VAERS Jun 18a | Any; Y | 162.2 (Pfizer) (12-15y)<br>94.02 (Pfizer) (16-17y) | Among 12-17 year old males, the incidence of presenting with myocarditis after vaccination with the Pfizer vaccine is probably between 55 (7 d risk) and 134 (30 d risk) cases per million | Moderate<sup>c</sup> |
|  |  | VAERS Oct 6 | 7 d; Y | 39.9 (Pfizer) (12-15y)<br>69.1 (Pfizer) (16-17y) [55] |  |  |
|  |  | Israel MOH May 31 | 30 d; Y | 150.7 (Pfizer) (16-19y) [134]* |  |  |
|  | 18-29y | VAERS Jun 30 | NR; N | 22-27 | Among 18-29 year old males, the incidence of presenting with myocarditis after vaccination with the mRNA vaccines is probably between 40 (≤7 d risk) and 99 (21-30 d risk) cases per million | Moderate<sup>c</sup> |
|  |  | VAERS Oct 6 | 7 d; Y | 36.8-38.5 (18-24y) [37.6]<br>10.8-17.2 (25-29y) [14]<br>[25.8 across ages] |  |  |
|  |  | Israel Defense Forces Mar 7 | 7d; Y | 50.7 (Pfizer; all cases males; 18-24 y) |  |  |
|  |  | US Military Apr 30 | 4 d (all cases); Y | 44 (median 25 y [20-51]) |  |  |
|  |  | Israel MOH May 31 | 30 d; Y | 150.7 (Pfizer) (16-19y) [134]<br>108.6 (Pfizer) (20-24y) [90.9]<br>69.9 (Pfizer) (25-29y) [50.4]<br>[average 91.8] |  |  |
|  |  | Clalit Health May 24a | 42 d after dose 1; Y | 106.9 (Pfizer; either dose; 94% had 2 doses) (16-29y) |  |  |
|  | 30-39y | VAERS Oct 6 | 7 d; Y | 5.2-6.7 [6.0] | Among 30-39 year old males, the incidence of presenting with myocarditis after vaccination with the mRNA vaccines may be between 6 (7 d risk) and 21 (30 d risk) cases per million | Low |
|  |  | Israel MOH May 31 | 30 d; N | 36.9 (Pfizer) [20.9] |  |  |
|  | ≥40y | VAERS Oct 6 | 7 d; Y | 0.1-2.9 [1.5] | Among males 40 and older, the incidence of presenting with myocarditis after vaccination with the mRNA vaccines may be fewer than 20 cases per million | Low |
|  |  | VAERS Jun 11 | 7 d; N | 2.4 (≥30y) |  |  |
|  |  | Israel MOH May 31 | 30 d; N | 6.8 (Pfizer) (average among 40-49y and ≥50y) |  |  |
|  |  | Clalit Health May 24a | 42 d after dose 1; Y | 21.1 (Pfizer; either dose; 94% had 2 doses) (≥30y) |  |  |
| F | 12-17y | VAERS Jun 18a | Any; Y | 13.0 (Pfizer) (12-15y)<br>13.4 (Pfizer) (16-17y) [13.2] | Among 12-17 year old females, the incidence of presenting with myocarditis after vaccination with the Pfizer vaccine may be fewer than 10 cases per million | Low |
|  |  | VAERS Oct 6 | 7 d; Y | 3.9 (Pfizer) (12-15y)<br>7.9 (Pfizer) (16-17y) [11.8] |  |  |
|  | 18-29y | VAERS Jun 30 | NR; N | 3-4 |  | Low |
|  |  | VAERS Oct 6 | 7 d; Y | 1.2-5.7 (25-29y) [3.5]<br>2.5-5.3 (18-24y) [4.0] | Among 18-29 year old females, the incidence of presenting with myocarditis after vaccination with the mRNA vaccines may be fewer than 10 cases per million |  |
|  |  | Israel MOH May 31 | 30 d; Y | 10.0 (Pfizer) (16-19y) [6.7]<br>21.6 (Pfizer) (20-24y) [18.7]<br>0 (Pfizer) (25-29y) [average 8.5] |  |  |
|  |  | Clalit Health May 24a | 42 d after dose 1; Y | 3.4 (Pfizer; either dose) (16-29y) |  |  |
|  | 30-39y | VAERS Oct 6 | 7 d; Y | 0.4-0.7 [0.55] | Among 30-39 year old females, there may be no excess risk of myocarditis after vaccination with the mRNA vaccines | Low |
|  |  | Israel MOH May 31 | 30 d; N | 2.2 (Pfizer) [2.2] |  |  |
|  | ≥40 | VAERS Oct 6 | 7 d; Y | 0.3-1.4 [0.85] | Among females 40 and older, there may be no excess risk of myocarditis after vaccination with the mRNA vaccines | Low |
|  |  | VAERS Jun 11 | 7 d; Y | 1.0 (≥30 y) |  |  |
|  |  | Israel MOH May 31 | 30 d; N | 1.9-4.5 (Pfizer) [3.2] |  |  |
|  |  | Clalit Health May 24a | 42 d after dose 1; Y | 2.0 (Pfizer; either dose) (≥30y) |  |  |
| Pericarditis |  |  |  |  |  |  |
| Both sexes | All ages | EudraVigilance Oct 19 | NR; N | 6.8-13.9 (either dose) [10.4] | Across all ages, we are very uncertain about the incidence of pericarditis after vaccination with mRNA vaccines | Very low<sup>a</sup> |
|  |  | Yellow Card Oct 13 | NR; N | 6-19 (either dose) [12.5] |  |  |
|  |  | Clalit Health May 24b | 42 d after dose 1; N | 10 (Pfizer; either dose) (≥16y) |  |  |
|  |  | Providence Health May 25 | 30d; Y | 18 (either dose) |  |  |
| Myocarditis/pericarditis |  |  |  |  |  |  |
| Both sexes | All ages | CAEFISS & CVP | None; Y (not all probable though) | 13.7-25.1 (either dose) [19.4] | Across all ages, we are very uncertain about the incidence of myocarditis or pericarditis after vaccination with mRNA vaccines | Very low<sup>a</sup> |
|  |  | VSD Oct 9 | 21 d; N | 9.7 (either dose) |  |  |
| Both sexes | 12-17y | VSD Oct 9 | 7 d; Y | 54 (Pfizer) | Among youth 12-17 years, the incidence of presenting with myocarditis or pericarditis after vaccination with the Pfizer vaccine may be approximately 50 cases (mostly males)** per million | Low |
|  |  | VSD Oct 9 | 21 d; Y | 56.7 (Pfizer) |  |  |
|  |  | Yellow Card Oct 13 | NR; N | 0-10 (either dose) |  |  |
| Both sexes | 18-39y | VSD Oct 9 | 7 d; Y | 21.5 (Pfizer) (12-39y)<br>13.1 (18-39y) | Among adults under 40, fewer than 20 cases (mostly males)** per million may present with myocarditis or pericarditis after vaccination with mRNA vaccines | Low |
|  |  | VSD Aug 21 | 21 d; Y | 13.6 (Pfizer) (12-39y) |  |  |
|  |  | Yellow Card Oct 13 | NR; N | 18-44 (either dose) (18-49y) [31] |  |  |

\*Crude incident rates were converted to excess incidence rates using the estimated aIRR from the study (excess=crude incidence –(crude incidence / aIRR); for males: aIRR 16-19 y 8.96 (95% CI, 4.50 to 17.83); 20-24 y 6.13 (95% CI 3.16 to 11.88); 25-29 y 3.58 (95% CI 1.82 to 7.01); ≥30 y 1.00 (95% CI, 0.61 to 1.64) (note: for the 30-39y old data we used an average of the 25-29 and ≥30y aIRRs); for females: 16-19y 2.95 (0.42–20.91), 20-24 y 7.56 (1.47–38.96), 25-29y 0, ≥30y 0.82 (0.33–2.02)(not used)
\*\* Case series data from the VSD Aug 21 found 56% pericarditis and 84% males (see Table 4)

**Table 3.** Summary of Findings for Comparison of Data in Studies Reporting on Moderna and Pfizer. Note: Shading represents the data used to create lower and upper limits for incidence rates, when used in conclusions. See Appendix 3 for references.

| Sex | Age | Study (by data source & date) | Risk interval; confirmed cases (Y/N) | Incidence/reporting rates per million doses by vaccine after dose 2, unless otherwise stated | Conclusions | Certainty about conclusions using GRADE |
| --- | --- | --- | --- | --- | --- | --- |
| **Myocarditis** |  |  |  |  |  |  |
| Both | All ages (≥12 unless stated) | VAERS Aug 6 | None applied; N | Moderna: 3.65 vs. Pfizer: 6.47 | Across all ages, we are very uncertain about whether there is an association between mRNA vaccine type and incidence of myocarditis | Very low<sup>a,b</sup> |
|  |  | Yellow Card Oct 13 | NR; N | Moderna: 32 (either dose) vs. Pfizer: 8 (either dose) |  |  |
|  |  | EudraVigilance Oct 19 | NR; N | Moderna: 25.8 (either dose) vs. Pfizer: 8.5 (either dose) |  |  |
| Both | <18 y | EudraVigilance Oct 19 | NR; N | Moderna: 60.3 (either dose) events per million vs. Pfizer: 41.6 (either dose) events per million | Among youth under 18 years, we are very uncertain about whether there is an association between mRNA vaccine type and incidence of myocarditis. | Very low<sup>b</sup> |
| Both | 18-29y | VSD Oct 9 | 7d; Y | Moderna vs. Pfizer (risk difference) 13.3 (aRR: 2.28 [95% CI, 1.25 to 6.05]) (18-39y) | Among 18-29 year old adults, we are very uncertain about whether there is an association between mRNA vaccine type and incidence of myocarditis. | Very low<sup>a</sup> |
|  |  | VSD Oct 9 | 21d; Y | Moderna vs. Pfizer (risk difference) 9.4 (aRR: 2.19 [95% CI, 0.98 to 4.97]) (18-39y) |  |  |
| M | 18-29y | VAERS Oct 6 | 7d; Y | Moderna: 38.5 vs. Pfizer: 36.8 (18-24y)<br>Moderna: 17.2 vs. Pfizer: 10.8 (25-29y) | Among 18-29 year old males, Moderna compared with Pfizer may be associated with a small increase (4-20 cases per million) in risk for myocarditis following vaccination | Low |
|  |  | VSD Oct 9 | 7d; Y | Moderna vs. Pfizer (risk difference) 19.1 (aRR: 2.14 [95% CI, 0.93 to 4.98]) (18-39y) |  |  |
|  | 30-39y | VAERS Oct 6 | 7d; Y | Moderna: 6.7 vs. Pfizer: 5.2 | Among 30-39 year old males, there may be little-to-no difference in risk of myocarditis after vaccination with Moderna compared with Pfizer | Low |
|  | ≥40y | VAERS Oct 6 | 7d; Y | Moderna: 2.9 vs. Pfizer: 2.0 (40-49 y) | Among 30-39 year old males, there may be little-to-no difference in risk of myocarditis after vaccination with Moderna compared with Pfizer | Low |
| F | 18-29y | VAERS Oct 6 | 7d; Y | Moderna: 5.3 vs. Pfizer: 2.5 (18-24y)<br>Moderna: 5.7 vs. Pfizer: 1.2 (25-29y) | Among 18-29 year old females, there may be little-to-no difference in risk of myocarditis after vaccination with Moderna compared with Pfizer | Low |
|  | 30-39y | VAERS Oct 6 | 7d; Y | Moderna: 0.4 vs. Pfizer: 0.7 | Among 30-39 year old females, there may be little-to-no difference in risk of myocarditis after vaccination with Moderna compared with Pfizer | Low |
|  | 40-49y | VAERS Oct 6 | 7d; Y | Moderna: 1.4 vs. Pfizer: 1.1 | Among 40-49 year old females, there may be little-to-no difference in risk of myocarditis after vaccination with Moderna compared with Pfizer | Low |
| Pericarditis |  |  |  |  |  |  |
| Both sexes | All ages | EEA Oct 19 | NR; N | Moderna: 13.9 vs. Pfizer: 6.8 | Across all ages, we are very uncertain about whether there is an association between mRNA vaccine type and incidence of myocarditis | Very low<sup>b</sup> |
|  |  | Yellow Card Oct 13 | NR; N | Moderna: 19 vs. Pfizer: 6 |  |  |
| Myocarditis/pericarditis |  |  |  |  |  |  |
| Both sexes | All ages | CAEFISS & CVP | None; Y (suspicious cases included) | Moderna: 25.1 vs. Pfizer: 13.7 | Across all ages, we are very uncertain about whether there is an association between mRNA vaccine type and incidence of myocarditis or pericarditis | Very low<sup>a,b</sup> |
| Both sexes | 12-17y | Yellow Card Oct 13 | NR; N | Moderna: 0 vs. Pfizer: 10 | Among youth 12-17 years old, we are very uncertain about whether there is an association between mRNA vaccine type and incidence of myocarditis or pericarditis | Very low<sup>a,b</sup> |
| Both sexes | 18-39y | VSD Oct 9 | 7d; Y | Moderna: 21.0 vs. Pfizer: 8.5; risk difference 13.3 (aRR: 2.72 [95% CI, 1.25 to 6.05]) (18-39y) | Among 18-39 year old adults, Moderna compared with Pfizer may be associated with a small increase (about 10 cases per million) in risk (mostly in males)* for myocarditis or pericarditis following vaccination | Low |
|  |  | VSD Oct 9 | 21d; Y | Moderna vs. Pfizer risk difference 11.5 (aRR: 2.19 [95% CI, 1.05 to 4.65]) (18-39y) |  |  |
|  |  | Yellow Card Oct 13 | NR; N | Moderna: 44 vs. Pfizer: 18 (18-49y) |  |  |

\* Case series data from the VSD studies (Aug) found 56% pericarditis and 84% males (see Table 4).

#### Research Question 2. Case Characteristics and Short-term Clinical Course

We included 11 case series reported in articles (either published, pre-proofs, or pre-prints)^9 34 44 47-50 53-56^ and three case series compiled from publicly available websites (Public Health Agency of Canada, PHAC; UK’s Medicines & Healthcare Products Regulatory Agency [Yellow Card database], UK; EudraVigilance, EEA).^32 37 38^ For one of the research articles (Klein, 2021),^47^ more recent data on some variables were provided in a publicly available presentation^46^ and information from both sources was used.

The 14 reports included a total of 12,636 cases (median 57, IQR 9-279), although it is important to note that there may be some overlap. All reports included clinician-confirmed cases except PHAC, Yellow Card, and EudraVigilance. **Table 4** (myocarditis or myopericarditis) and **Table 5** (pericarditis and mixed diagnosis [i.e., myocarditis, pericarditis and/or myopericarditis]) include data from the 14 case series.

**Table 4.**
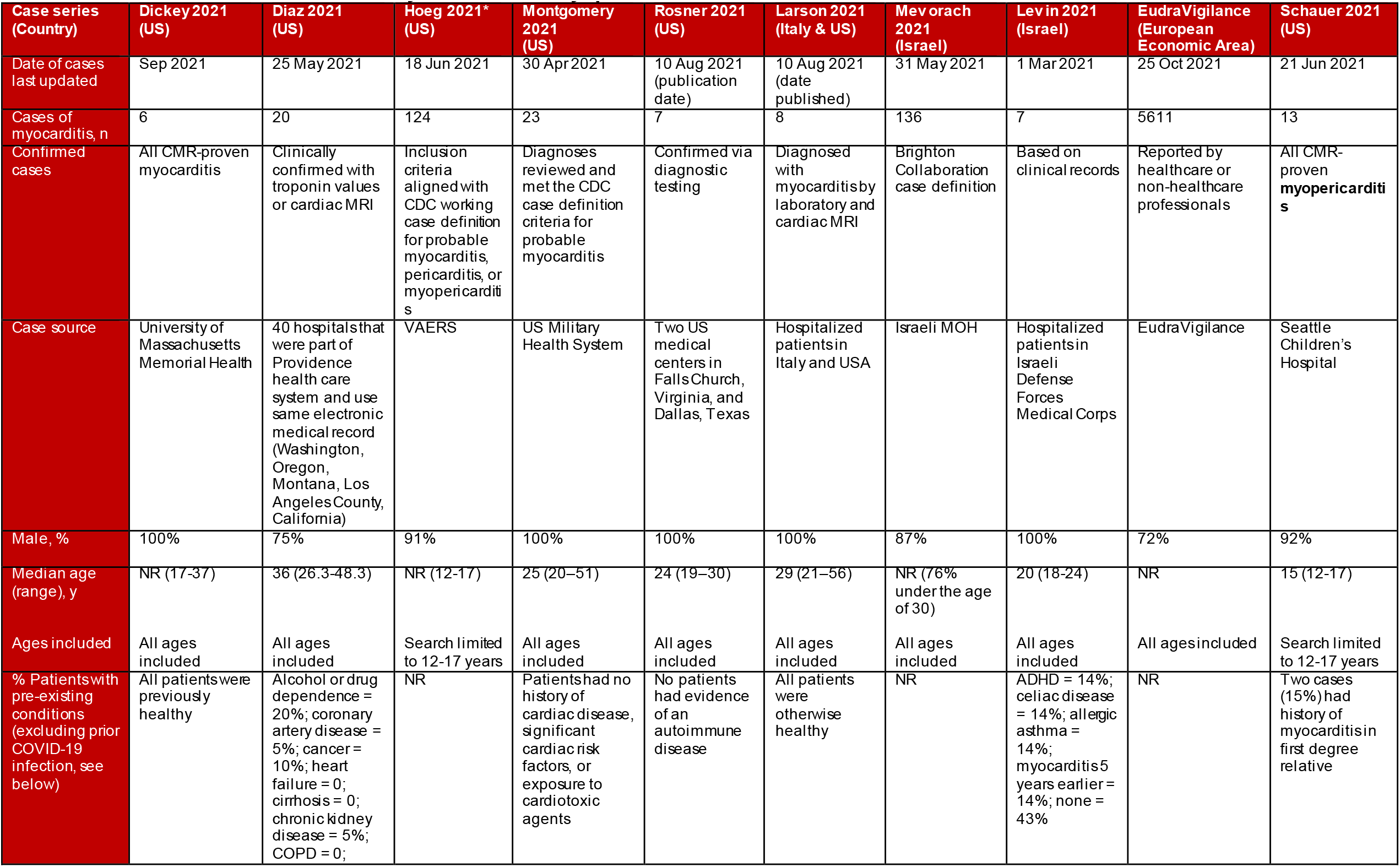
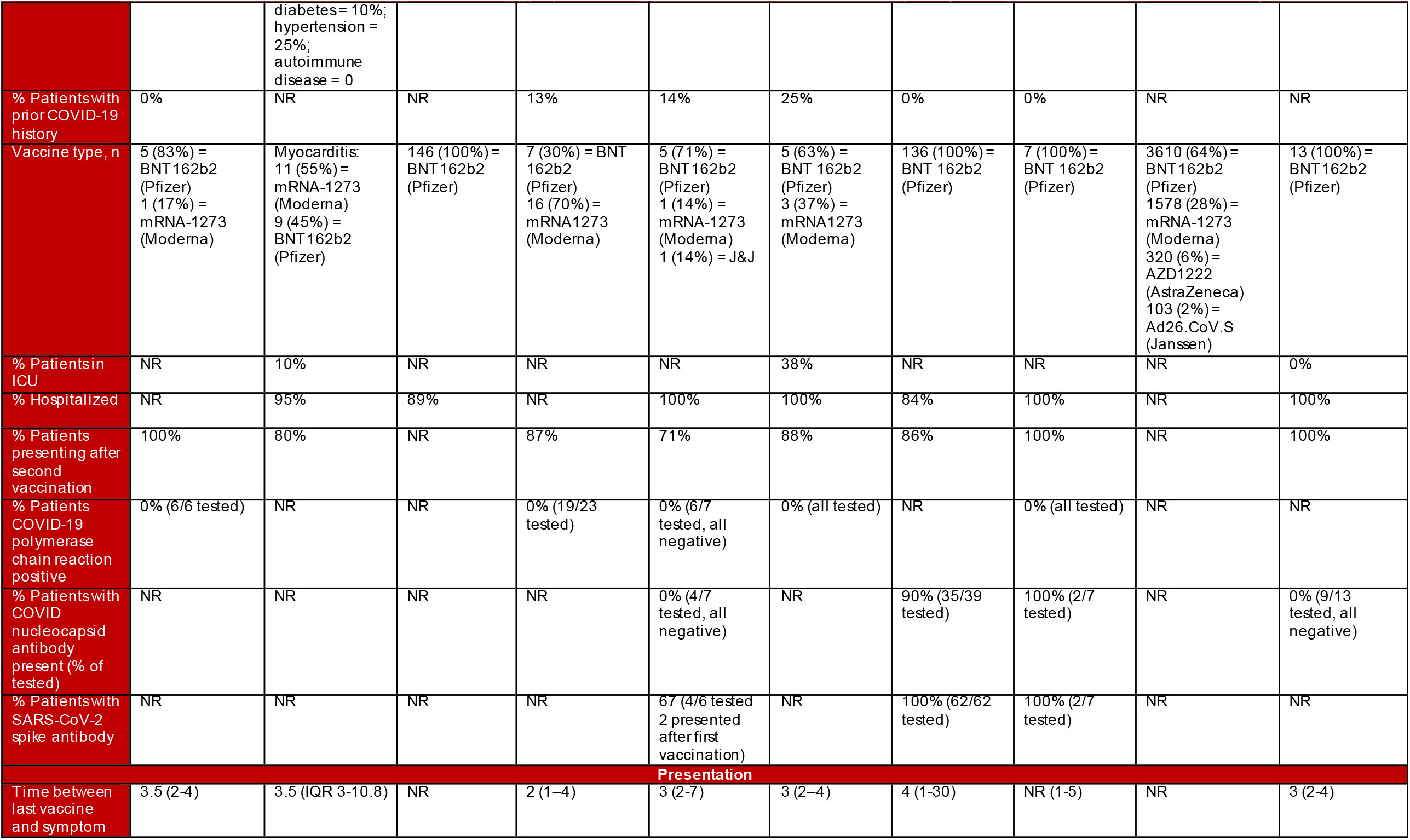
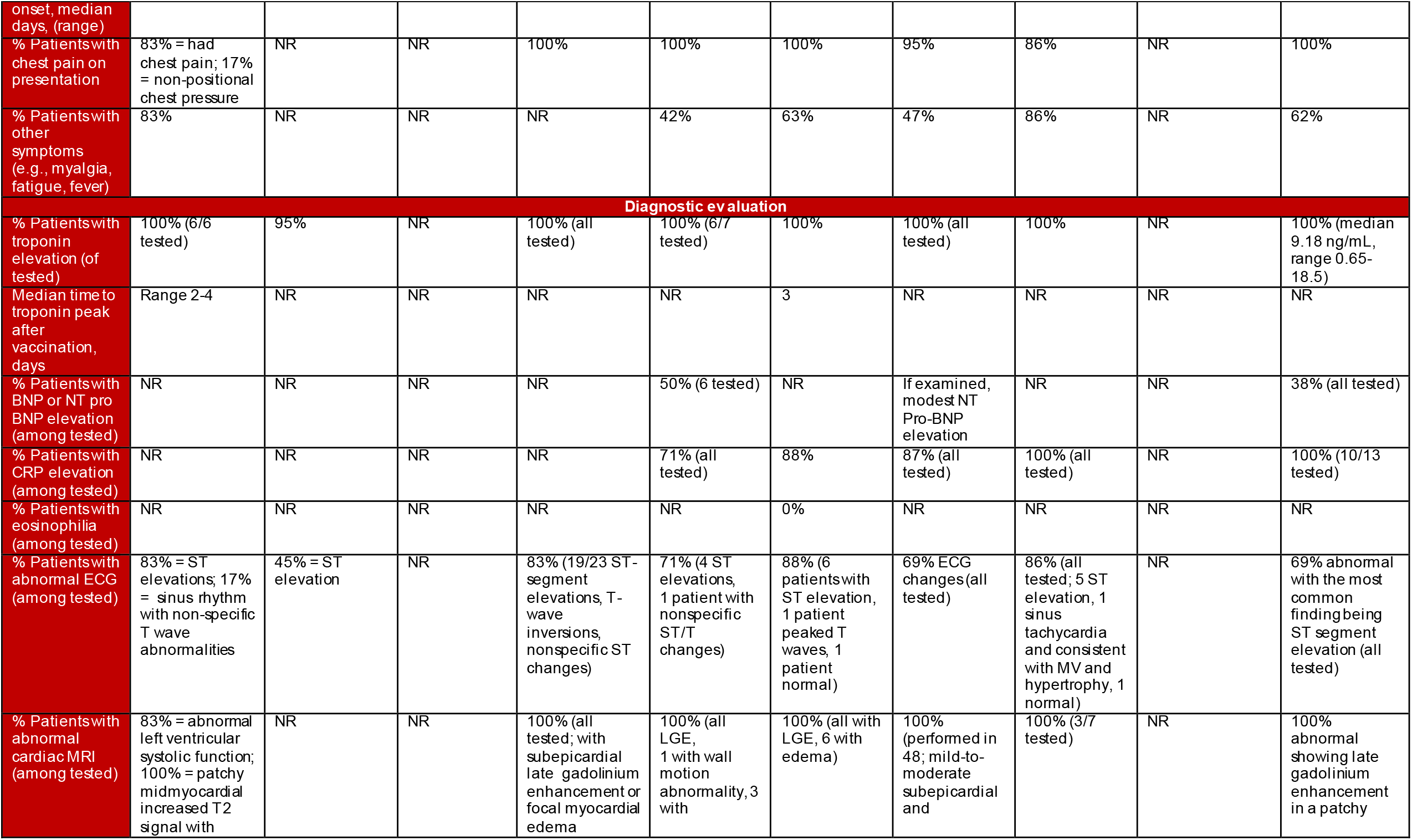
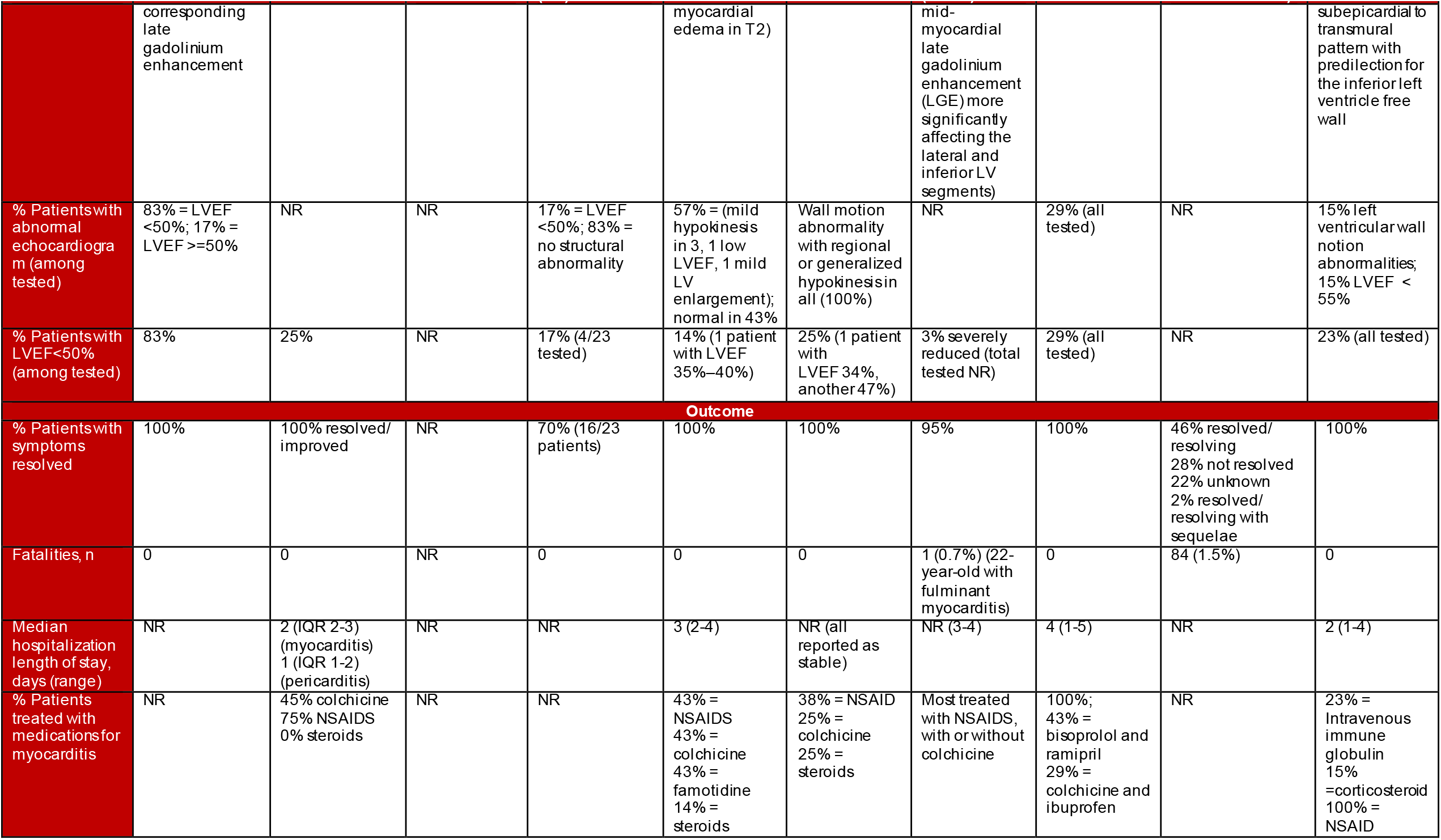
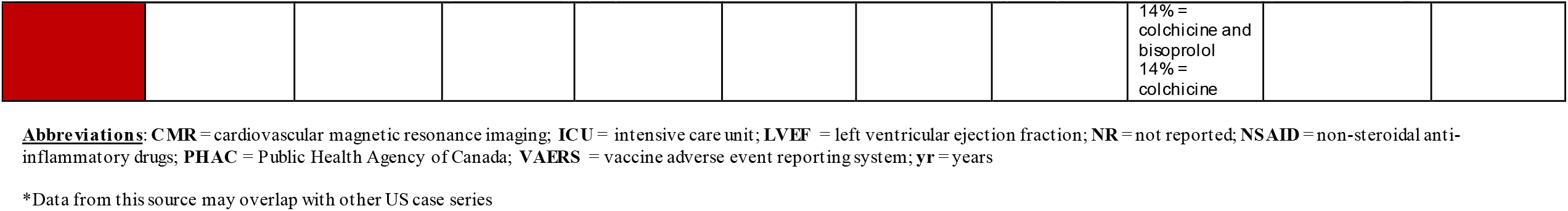
Results from Case Series of Myocarditis and Myopericarditis after COVID-19 Vaccination.

| Case series (Country) | Dickey 2021 (US) | Diaz 2021 (US) | Hoeg 2021* (US) | Montgomery 2021 (US) | Rosner 2021 (US) | Larson 2021 (Italy & US) | Mevorach 2021 (Israel) | Levin 2021 (Israel) | EudraVigilance (European Economic Area) | Schauer 2021 (US) |
| --- | --- | --- | --- | --- | --- | --- | --- | --- | --- | --- |
| Date of cases last updated | Sep 2021 | 25 May 2021 | 18 Jun 2021 | 30 Apr 2021 | 10 Aug 2021 (publication date) | 10 Aug 2021 (date published) | 31 May 2021 | 1 Mar 2021 | 25 Oct 2021 | 21 Jun 2021 |
| Cases of myocarditis, n | 6 | 20 | 124 | 23 | 7 | 8 | 136 | 7 | 5611 | 13 |
| Confirmed cases | All CMR-proven myocarditis | Clinically confirmed with troponin values or cardiac MRI | Inclusion criteria aligned with CDC working case definition for probable myocarditis, pericarditis, or myopericarditis | Diagnoses reviewed and met the CDC case definition criteria for probable myocarditis | Confirmed via diagnostic testing | Diagnosed with myocarditis by laboratory and cardiac MRI | Brighton Collaboration case definition | Based on clinical records | Reported by healthcare or non-healthcare professionals | All CMR-proven myopericarditis |
| Case source | University of Massachusetts Memorial Health | 40 hospitals that were part of Providence health care system and use same electronic medical record (Washington, Oregon, Montana, Los Angeles County, California) | VAERS | US Military Health System | Two US medical centers in Falls Church, Virginia, and Dallas, Texas | Hospitalized patients in Italy and USA | Israeli MOH | Hospitalized patients in Israeli Defense Forces Medical Corps | EudraVigilance | Seattle Children's Hospital |
| Male, % | 100% | 75% | 91% | 100% | 100% | 100% | 87% | 100% | 72% | 92% |
| Median age (range), y | NR (17-37) | 36 (26.3-48.3) | NR (12-17) | 25 (20-51) | 24 (19-30) | 29 (21-56) | NR (76% under the age of 30) | 20 (18-24) | NR | 15 (12-17) |
| Ages included | All ages included | All ages included | Search limited to 12-17 years | All ages included | All ages included | All ages included | All ages included | All ages included | All ages included | Search limited to 12-17 years |
| % Patients with pre-existing conditions (excluding prior COVID-19 infection, see below) | All patients were previously healthy | Alcohol or drug dependence = 20%; coronary artery disease = 5%; cancer = 10%; heart failure = 0; cirrhosis = 0; chronic kidney disease = 5%; COPD = 0; | NR | Patients had no history of cardiac disease, significant cardiac risk factors, or exposure to cardiotoxic agents | No patients had evidence of an autoimmune disease | All patients were otherwise healthy | NR | ADHD = 14%; celiac disease = 14%; allergic asthma = 14%; myocarditis 5 years earlier = 14%; none = 43% | NR | Two cases (15%) had history of myocarditis in first degree relative |
|  |  | diabetes = 10%; hypertension = 25%; autoimmune disease = 0 |  |  |  |  |  |  |  |  |
| % Patients with prior COVID-19 history | 0% | NR | NR | 13% | 14% | 25% | 0% | 0% | NR | NR |
| Vaccine type, n | 5 (83%) = BNT162b2 (Pfizer)<br>1 (17%) = mRNA-1273 (Moderna) | Myocarditis: 11 (55%) = mRNA-1273 (Moderna)<br>9 (45%) = BNT 162b2 (Pfizer) | 146 (100%) = BNT162b2 (Pfizer) | 7 (30%) = BNT 162b2 (Pfizer)<br>16 (70%) = mRNA1273 (Moderna) | 5 (71%) = BNT162b2 (Pfizer)<br>1 (14%) = mRNA-1273 (Moderna)<br>1 (14%) = J&J | 5 (63%) = BNT 162b2 (Pfizer)<br>3 (37%) = mRNA1273 (Moderna) | 136 (100%) = BNT 162b2 (Pfizer) | 7 (100%) = BNT 162b2 (Pfizer) | 3610 (64%) = BNT162b2 (Pfizer)<br>1578 (28%) = mRNA-1273 (Moderna)<br>320 (6%) = AZD1222 (AstraZeneca)<br>103 (2%) = Ad26.CoV.S (Janssen) | 13 (100%) = BNT162b2 (Pfizer) |
| % Patients in ICU | NR | 10% | NR | NR | NR | 38% | NR | NR | NR | 0% |
| % Hospitalized | NR | 95% | 89% | NR | 100% | 100% | 84% | 100% | NR | 100% |
| % Patients presenting after second vaccination | 100% | 80% | NR | 87% | 71% | 88% | 86% | 100% | NR | 100% |
| % Patients COVID-19 polymerase chain reaction positive | 0% (6/6 tested) | NR | NR | 0% (19/23 tested) | 0% (6/7 tested, all negative) | 0% (all tested) | NR | 0% (all tested) | NR | NR |
| % Patients with COVID nucleocapsid antibody present (% of tested) | NR | NR | NR | NR | 0% (4/7 tested, all negative) | NR | 90% (35/39 tested) | 100% (2/7 tested) | NR | 0% (9/13 tested, all negative) |
| % Patients with SARS-CoV-2 spike antibody | NR | NR | NR | NR | 67 (4/6 tested 2 presented after first vaccination) | NR | 100% (62/62 tested) | 100% (2/7 tested) | NR | NR |
| **Presentation** |  |  |  |  |  |  |  |  |  |  |
| Time between last vaccine and symptom | 3.5 (2-4) | 3.5 (IQR 3-10.8) | NR | 2 (1-4) | 3 (2-7) | 3 (2-4) | 4 (1-30) | NR (1-5) | NR | 3 (2-4) |
| onset, median days, (range) |  |  |  |  |  |  |  |  |  |  |
| % Patients with chest pain on presentation | 83% = had chest pain; 17% = non-positional chest pressure | NR | NR | 100% | 100% | 100% | 95% | 86% | NR | 100% |
| % Patients with other symptoms (e.g., myalgia, fatigue, fever) | 83% | NR | NR | NR | 42% | 63% | 47% | 86% | NR | 62% |
| **Diagnostic evaluation** |  |  |  |  |  |  |  |  |  |  |
| % Patients with troponin elevation (of tested) | 100% (6/6 tested) | 95% | NR | 100% (all tested) | 100% (6/7 tested) | 100% | 100% (all tested) | 100% | NR | 100% (median 9.18 ng/mL, range 0.65-18.5) |
| Median time to troponin peak after vaccination, days | Range 2-4 | NR | NR | NR | NR | 3 | NR | NR | NR | NR |
| % Patients with BNP or NT pro BNP elevation (among tested) | NR | NR | NR | NR | 50% (6 tested) | NR | If examined, modest NT Pro-BNP elevation | NR | NR | 38% (all tested) |
| % Patients with CRP elevation (among tested) | NR | NR | NR | NR | 71% (all tested) | 88% | 87% (all tested) | 100% (all tested) | NR | 100% (10/13 tested) |
| % Patients with eosinophilia (among tested) | NR | NR | NR | NR | NR | 0% | NR | NR | NR | NR |
| % Patients with abnormal ECG (among tested) | 83% = ST elevations; 17% = sinus rhythm with non-specific T wave abnormalities | 45% = ST elevation | NR | 83% (19/23 ST-segment elevations, T-wave inversions, nonspecific ST changes) | 71% (4 ST elevations, 1 patient with nonspecific ST/T changes) | 88% (6 patients with ST elevation, 1 patient peaked T waves, 1 patient normal) | 69% ECG changes (all tested) | 86% (all tested; 5 ST elevation, 1 sinus tachycardia and consistent with MV and hypertrophy, 1 normal) | NR | 69% abnormal with the most common finding being ST segment elevation (all tested) |
| % Patients with abnormal cardiac MRI (among tested) | 83% = abnormal left ventricular systolic function; 100% = patchy midmyocardial increased T2 signal with | NR | NR | 100% (all tested; with subepicardial late gadolinium enhancement or focal myocardial edema) | 100% (all LGE, 1 with wall motion abnormality, 3 with | 100% (all with LGE, 6 with edema) | 100% (performed in 48; mild-to-moderate abnormality and | 100% (3/7 tested) | NR | 100% abnormal showing late gadolinium enhancement in a patchy |
|  | corresponding late gadolinium enhancement |  |  |  | myocardial edema in T2) |  | mid-myocardial late gadolinium enhancement (LGE) more significantly affecting the lateral and inferior LV segments) |  |  | subepicardial to transmural pattern with predilection for the inferior left ventricle free wall |
| % Patients with abnormal echocardiogram (among tested) | 83% = LVEF <50%; 17% = LVEF ≥50% | NR | NR | 17% = LVEF <50%; 83% = no structural abnormality | 57% = (mild hypokinesis in 3, 1 low LVEF, 1 mild LV enlargement); normal in 43% | Wall motion abnormality with regional or generalized hypokinesis in all (100%) | NR | 29% (all tested) | NR | 15% left ventricular wall motion abnormalities; 15% LVEF < 55% |
| % Patients with LVEF <50% (among tested) | 83% | 25% | NR | 17% (4/23 tested) | 14% (1 patient with LVEF 35%–40%) | 25% (1 patient with LVEF 34%, another 47%) | 3% severely reduced (total tested NR) | 29% (all tested) | NR | 23% (all tested) |
| **Outcome** |  |  |  |  |  |  |  |  |  |  |
| % Patients with symptoms resolved | 100% | 100% resolved/improved | NR | 70% (16/23 patients) | 100% | 100% | 95% | 100% | 46% resolved/resolving<br>28% not resolved<br>22% unknown<br>2% resolved/resolving with sequelae | 100% |
| Fatalities, n | 0 | 0 | NR | 0 | 0 | 0 | 1 (0.7%) (22-year-old with fulminant myocarditis) | 0 | 84 (1.5%) | 0 |
| Median hospitalization length of stay, days (range) | NR | 2 (IQR 2-3) (myocarditis)<br>1 (IQR 1-2) (pericarditis) | NR | NR | 3 (2-4) | NR (all reported as stable) | NR (3-4) | 4 (1-5) | NR | 2 (1-4) |
| % Patients treated with medications for myocarditis | NR | 45% colchicine<br>75% NSAIDS<br>0% steroids | NR | NR | 43% = NSAIDS<br>43% = colchicine<br>43% = famotidine<br>14% = steroids | 38% = NSAID<br>25% = colchicine<br>25% = steroids | Most treated with NSAIDS, with or without colchicine | 100%;<br>43% = bisoprolol and ramipril<br>29% = colchicine and ibuprofen | NR | 23% = Intravenous immune globulin<br>15% = corticosteroid<br>100% = NSAID |
|  |  |  |  |  |  |  |  | 14% = colchicine and bisoprolol<br>14% = colchicine |  |  |

**Abbreviations:** CMR = cardiovascular magnetic resonance imaging; ICU = intensive care unit; LVEF = left ventricular ejection fraction; NR = not reported; NSAID = non-steroidal anti-inflammatory drugs; PHAC = Public Health Agency of Canada; VAERS = vaccine adverse event reporting system; yr = years
\*Data from this source may overlap with other US case series

**Table 5.** Results from Case Series of Pericarditis or Mixed Diagnoses after COVID-19 Vaccination.

| Case series | Diaz 2021 (US) | Hoeg 2021* (US) | EudraVigilance (European Economic Area) | Gargano 2021* (US) | Klein 2021 (US) | Yellow Card (UK) | PHAC 2021 (Canada) |
| --- | --- | --- | --- | --- | --- | --- | --- |
| Date of cases last updated | 25 May 2021 | 18 Jun 2021 | 25 Oct 2021 | 6 Jul 2021 | 21 Aug 2021 | 13 Oct 2021 | 15 Oct 2021 |
| Cases, n | 37 | 22 | 4250 | 323 | 56 | 1037 | 956 |
| Confirmed cases | Clinically confirmed with troponin values or cardiac MRI | Inclusion criteria aligned with CDC working case definition for probable myocarditis, pericarditis, or myopericarditis | Reported by healthcare or non-healthcare professionals | All met CDC criteria for myocarditis, pericarditis, or myopericarditis | ICD-10 used then diagnoses confirmed by medical record review | NR | Brighton Collaboration case definition for myocarditis/pericarditis (inflammation of the heart muscle and lining around heart) |
| Myocarditis, % | 0% | 0% | 0% | NR | 30% | 52% | NR |
| Pericarditis, % | 100% | 100% | 100% | NR | 56% | 48% | NR |
| Myopericarditis, % | 0% | 0% | 0% | NR | 14% | 0% | 0% |
| Case source | 40 hospitals that were part of Providence health care system and use same electronic medical record (Washington, Oregon, Montana, Los Angeles County, California) | VAERS | EudraVigilance | VAERS | Kaiser Permanente: Colorado, Northern and Northwest, Southern California, Washington; Marshfield Clinic; Health Partners; and Denver Health (from Vaccine Safety Datalink) | Yellow Card Scheme | Canadian Adverse Events Following Immunization Surveillance System and Health Canada's Canada Vigilance System |
| Male, % | 73% | 91% | 54% | 90% | 84% | NR | 62% Pfizer, 73% Moderna |
| Median age (range), y | 59 (46-69) | NR (12-17) | NR | 19 (12-29) | 24 (13-39) | NR | 27 (12-87 Pfizer), (17-95 Moderna) |
| Ages included | All ages included | Search limited to 12-17 years | All ages included | Limited to those under 30 years | Medical record search limited to 12-39 years | All ages included | All ages included |
| % Patients with pre-existing conditions (excluding prior COVID-19 infection, see below) | Alcohol or drug dependence = 14%; coronary artery disease = 11%; cancer = 14%; heart failure = 5%; cirrhosis = 3%; chronic kidney disease = 11%; COPD = 11; diabetes = 11%; hypertension = 49%; autoimmune disease = 8% | NR | NR | NR | History of myo/pericarditis = 7% | NR | NR |
| % Patients with prior COVID-19 history | NR | NR | NR | NR | 5% | NR | NR |
| Vaccine type, n | 12 (32%) = mRNA-1273<br>23 (62%) = BNT162b2 (Pfizer)<br>2 (5%) = Ad26.CoV.S (Janssen) | 146 (100%) = BNT162b2 (Pfizer) | 2883 (68%) = BNT162b2 (Pfizer)<br>850 (20%) = mRNA-1273 (Moderna)<br>416 (10%) = AZD1222 (AstraZeneca)<br>101 (2%) = Ad26.CoV.S (Janssen) | NR | 14 (41%) = BNT162b2 (Pfizer)<br>20 (59%) = mRNA1273 (Moderna)* | Myocarditis:<br>337 (56%) = BNT162b2 (Pfizer)<br>86 (62%) = mRNA-1273 (Moderna)<br>120 (40%) = AZD1222 (AstraZeneca)<br>Pericarditis:<br>265 (44%) = BNT162b2 (Pfizer)<br>52 (38%) = mRNA-1273 (Moderna)<br>177 (60%) = AZD1222 (AstraZeneca) | 573 (60%) = BNT162b2 (Pfizer)<br>357 (37%) = mRNA-1273 (Moderna)<br>21 (2%) = AZD1222 (AstraZeneca)<br>5 (5%) = unspecified vaccine |
| % Patients in ICU | 2.7% | NR | NR | NR | 13% | NR | NR |
| % Hospitalized | 35% | 89% (myocarditis)<br>73% (pericarditis) | NR | 96% | 71% | NR | NR |
| % Patients presenting after second vaccination | 60% | NR | NR | NR | 71%* | NR | 45% Pfizer, 68% Moderna |
| % Patients COVID-19 polymerase chain reaction positive | NR | NR | NR | NR | NR | NR | NR |
| % Patientswith COVID nucleocapsid antibody present (% of tested) | NR | NR | NR | NR | NR | NR | NR |
| % Patientswith SARS-CoV-2 spike antibody | NR | NR | NR | NR | NR | NR | NR |
| **Presentation** |  |  |  |  |  |  |  |
| Time between last vaccine and symptom onset, median days, (range) | 20 (IQR 6-41) | NR | NR | 2 (0-40) | 12-17 yr= 2 (1-20)<br>18-29 yr= 1 (0-11)<br>30-39 yr= 5 (1-20) | NR | NR (5 min-92 d Pfizer), (17 min-69 d Moderna) |
| % Patientswith chest pain on presentation | NR | NR | NR | NR | 100% | NR | NR |
| % Patientswith other symptoms(e.g., myalgia, fatigue, fever) | NR | NR | NR | NR | 59% | NR | NR |
| **Diagnostic evaluation** |  |  |  |  |  |  |  |
| % Patientswith troponin elevation (of tested) | 0% | NR | NR | NR | 84% abnormal troponin level (all tested) | NR | 100% |
| Median time to troponin peakafter vaccination, days | NR | NR | NR | NR | NR | NR | NR |
| % Patientswith BNP or NT pro BNP elevation (among tested) | NR | NR | NR | NR | NR | NR | NR |
| % Patientswith CRP elevation (among tested) | NR | NR | NR | NR | NR | NR | NR |
| % Patientswith eosinophilia (among tested) | NR | NR | NR | NR | NR | NR | NR |
| % Patientswith abnormal ECG (among tested) | 38%= ST elevation | NR | NR | NR | 84% (all tested) | NR | NR |
| % Patientswith abnormal cardiac MRI (among tested) | NR | NR | NR | NR | 92% (12/56 tested) | NR | NR |
| % Patientswith abnormal echocardiogram (among tested) | NR | NR | NR | NR | 46% (52/56 tested) | NR | NR |
| % Patientswith LVEF<50% (among tested) | 8% | NR | NR | NR | NR | NR | NR |
| **Outcome** |  |  |  |  |  |  |  |
| % Patientswith symptoms resolved | 81% resolved/ improved; 5% | NR | 49% resolved/ resolving<br>31% not resolved | 95% (at time of publication) | 100% | NR | NR |
|  | persistent; 14% unknown |  | 19% unknown<br>1% resolved/<br>resolving with sequelae |  |  |  |  |
| Fatalities, n | 0 | NR | 15 (0.4%) | 0 | NR | 5 total (0.5%)<br>3 = BNT162b2 (Pfizer)<br>0 = mRNA-1273 (Moderna)<br>2 = AZD1222 (AstraZeneca) | NR |
| Median hospitalization length of stay, days (range) | 1 (IQR 1-2) | NR | NR | NR | 0 days = 14%<br>1 day = 38%<br>2 days = 23%<br>3 days = 13%<br>4 days = 5%<br>5 days = 4%<br>≥6 days = 4% | NR | NR |
| % Patients treated with medications for myocarditis | 54% colchicine<br>49% NSAIDs<br>11% steroids | NR | NR | NR | NR | NR | NR |

**Abbreviations:** CMR = cardiovascular magnetic resonance imaging; ICU = intensive care unit; LVEF = left ventricular ejection fraction; NR = not reported; NSAID = non-steroidal anti-inflammatory drugs.
\*Data from this source may overlap with other US case series.

Most case series focused on myocarditis (10 reports, n=5,955). The majority of cases (often >90%) involved males with reported median age most often between 20 and 29 years; confirmed cases ranged from 12 to 56 years. Time between last vaccine and symptom onset was on average 2 to 4 days, and the majority (71-100%) presented after a second dose. Most cases presented with chest pain or pressure, and troponin elevation; a minority (<30%) showed left ventricular dysfunction (i.e., LVEF<50%).

The majority of myocarditis cases were hospitalized (≥84%) with few admitted to ICU; average length of hospital stay was 2-4 days. NSAIDs were most often used as treatment; other interventions included bisoprolol, ramipril, colchicine, famotidine, steroids (for myocarditis) and intravenous immune globulin (for myopericarditis). Among the series of confirmed cases of myocarditis that reported on fatalities (N=220), one fatality was reported in Israel in a 22-year-old with fulminant myocarditis. The unconfirmed series from the EEA reported 84 fatalities among 5,611 cases (1.5%), though cause of death was not confirmed.

Three reports provided data for pericarditis (n=4,309; almost 99% unconfirmed). The majority involved males; however, there was variation across reports (54 to 91%). Median age (59 years) was only reported in one small series (n=37). The same series reported a median interval of 20 days between last vaccine and symptom onset, with 60% presenting after the second vaccination. Hospitalization varied (35% and 73% reported in two series with a total of 59 cases); one series (n=37) reported 3% admitted to ICU and 1 day for median length of stay. One series (n=37) reported 0 fatalities, and a larger series of unconfirmed cases (n=4,250) reported 15 deaths (0.4%).

Four reports (n=2,372) included a mix of diagnoses. The majority of patients were males. Median age was 27 years in the report that included all ages (n=956). Time between last vaccine and symptom onset was median 2 days (based on 323 cases). The majority presented after second vaccination (based on 56 cases). The majority were hospitalized (based on 379 cases), with a minority admitted to ICU (13% of n=56). Length of stay was ≤2 days in 75% of patients hospitalized (based on 56 cases). No fatalities were reported in two series (total 379 cases); a third report identified 5 fatalities among 1,037 unconfirmed cases (0.5%).

## Discussion

The incidence of myocarditis following mRNA vaccines is low but probably highest in males 12-29 years old, with lower incidences in older ages. In females, the incidence may be very low (12-29 y) or little-to-none (≥30 y). The incidence of either myocarditis or pericarditis may be highest in adolescent males; among adults under 40 years there may be few cases but the broad age range may not have detected any variation by age. Among adult males under 40 years, Moderna compared with Pfizer may be associated with a small increase (<20 cases per million) in risk for myocarditis or (one of) myocarditis or pericarditis following vaccination; the evidence for youth under 18 years was very uncertain. This evidence does not strongly support that one mRNA vaccine should be preferred over the other, even in young males. Although not systematically reviewed in this report, the incidence of myocarditis after vaccination appears to be much lower than that reported after becoming infected with COVID-19 (450^57^ to 1,200^58^ per million in young males).

From the case series, the majority of myocarditis cases involved males (often >90%) in their 20s, with a short symptom onset of 2 to 4 days after a second dose (71-100%). The large majority of cases presented with chest pain or pressure and troponin elevation, and a minority (<30%) also had left ventricular dysfunction. Most were hospitalized (≥84%), without ICU stays, for a short duration (2-4 d) and treated with anti-inflammatory and/or other standard supportive therapies. Among the series of confirmed cases that reported on fatalities (n=220), one fatality was reported in a 22-year old with fulminant myocarditis. Within the EudraVigilance dataset (n=5,611 unconfirmed cases), 84 (1.5%) fatalities were reported, though cause of death is not confirmed. Importantly, the vast majority of affected individuals appear to make a complete recovery based on short-term follow-up. Most cases of pericarditis were unconfirmed; for this outcome there appears to be more variation in age, sex, onset timing and rate of hospitalization across cases. The findings across cases appear to be similar to non-vaccine related myo- and pericarditis whereby young males are at highest risk and mortality is very rare.^13^

Although the mechanism(s) through which the mRNA vaccines may cause myocarditis or pericarditis are not known, several findings in the studies in this review support a causal association, including large IRRs and excess risks compared with controls, highly significant clustering effects showing onset soon after vaccine receipt,^40 47^ and higher risk for younger males as seen with other causative agents of myocarditis or pericarditis. Further, an immune-mediated mechanism is suspected, given the much higher incidence of myocarditis after the second compared with first dose of mRNA vaccine..^40 45 47 49-51.^

The key messages from the patient/parent perspective, co-developed with our patient partners, include:

▪ A risk for myocarditis after COVID-19 vaccination seems to exist for young males (<29y) although is likely very small and occurrences appear to be quite mild with full recovery.
▪ Clear and effective communication of the risks (rates of myocarditis and likely clinical course), benefits from vaccination, and the availability of good alternatives will be critical for young males and their parents.
▪ Getting vaccinated is important for young males (for their contacts and themselves) and recommendations against vaccines, even if cautionary or against a single vaccine, may lead to unnecessary harm from increasing hesitancy; recommendations could focus on positive guidance about awareness and identification of symptoms for these rare and relatively mild risks.
▪ More research on what personal risk factors (e.g., pre-existing conditions) may put someone at higher risk would be very useful.

### Strengths and limitations of the review

There are several strengths of this review. A comprehensive, peer-reviewed search strategy was used and inclusion of gray literature captured in several cases very recent data. A second reviewer screened the most relevant (based on machine learning) citations, and verified all data and risk of bias assessments. GRADE assessments were based on team consensus including clinical experts. Patient partners reviewed the evidence and developed interpretations from the patient perspective. The main limitation is that in the era of COVID-19 the literature base is evolving with incredible rapidity and new evidence will emerge; nevertheless, there was some moderate certainty evidence found in this review. Because many reports used overlapping populations and reported findings based on different methods of case ascertainment (e.g., risk interval, whether cases were verified) a quantitative synthesis was not undertaken and some of the descriptive summary statements may not fully account for this. We also avoided making any conclusions about any one possible estimate of average incidence and instead relied on ranges.

## Conclusions

Policy and decision making about vaccination for adolescent and young adult males will need to weigh the possibility of myocarditis with various factors such as the i) societal benefits from preventing COVID-19 transmission, ii) individual, age-relevant benefits from prevention or mitigating severe COVID-19 related illness and iii) availability and efficacy of the non-mRNA vaccines. Although the short-term course of myocarditis after an mRNA vaccine is likely mild and self-limiting in the majority of cases, the potential for long-term sequelae such as recurrent disease and/or heart failure are not known. Continued active surveillance of myocarditis/myopericarditis incidence out to 30 days from dosing is recommended with respect to i) new populations (i.e., vaccinated children <12y), ii) the impact of 3rd and subsequent doses, and iii) affected individuals receiving subsequent mRNA vaccine doses. Future research is also needed for determining other risk factors for myocarditis after vaccination apart from age and sex.

## Data Availability

All data produced in the present work are contained in the manuscript and supplement

## Abbreviations

CAEFISS: Canadian Adverse Events Following Immunization Surveillance System
CVP: Canada Vigilance Program
CDC: Centers for Disease Control and Prevention
EEA: European Economic Area
ICU: Intensive care unit
IRR: Incidence rate ratio
KPSC: Kaiser Permanente Southern California
MOH: Ministry of Health
PHAC: Public Health Agency of Canada
RCT: randomized controlled trial
VSD: Vaccine Safety Datalink
VAERS: Vaccine Adverse Events Reporting System.

## Contributors

JP, LH, AM and IP designed the study. JP, LH, LB, LG, and AW screened citations for inclusion and were involved in data extraction and interpretation. JP, LB, LG and AW were involved is risk of bias assessments. All authors were involved in interpretation of the data. JP wrote the draft manuscript with input from all authors. All authors approved the final version of the manuscript. LH and JP are the guarantors of this manuscript and accept full responsibility for the work and the conduct of the study, had access to the data, and controlled the decision to publish. The corresponding author attests that all listed authors meet authorship criteria and that no others meeting the criteria have been omitted.

## Acknowledgements

The authors thank the patient/public partners Linda Wilhelm (New Brunswick) and Natasha Trehan (Toronto, Ontario) for contributing key message from their perspective; Susanna Ogunnaike-Cooke and Natalia Abraham (Knowledge Users; Public Health Agency of Canada) and Dr. Andrea Tricco (Unity Health Toronto, Ontario and SPOR Evidence Alliance) for reviewing the protocol and draft report from which this article stems; and Kaitryn Campbell, MLS, MSc (St. Joseph’s Healthcare Hamilton/McMaster University) for peer reviewing the search strategy.

## Funding Acknowledgement(s) & Disclaimer

This project was funded by the Canadian Institutes of Health Research (CIHR) through the COVID-19 Evidence Network to support Decision-making (COVID-END) at McMaster University and the SPOR Evidence Alliance. The funders provided input on the design of the study, but did not take part in the collection, analysis, or interpretation of data. They reviewed a report generated from the findings but did not make decisions about submission of the article for publication.

This review was developed through the analysis, interpretation and synthesis of scientific research and/or systematic reviews published in peer-reviewed journals, institutional websites and other distribution channels. This document may not fully reflect all the scientific evidence available at the time this report was prepared. Other relevant scientific findings may have been reported since completion of this synthesis report.

SPOR Evidence Alliance, COVID-END and the project team at ARCHE make no warranty, express or implied, nor assume any legal liability or responsibility for the accuracy, completeness, or usefulness of any information, data, product, or process disclosed in this report. Conclusions drawn from, or actions undertaken on the basis of, information included in this report are the sole responsibility of the user.

Dr. Hartling is supported by a Canada Research Chair in Knowledge Synthesis and Translation, and is a Distinguished Researcher with the Stollery Science Lab supported by the Stollery Children’s Hospital Foundation.

## Ethics approval

This research did not involve human research participants and thus did not require ethics approval.

## Transparency declaration

The lead authors of this work, JP and LH, affirm that the manuscript is an honest, accurate, and transparent account of the study being reported; that no important aspects of the study have been omitted; and that any discrepancies from the study as originally planned in the protocol have been explained.

## Data sharing

All of the data extracted for this review are included in the manuscript and associated supplementary files.

## References

1. Bautista García J, Peña Ortega P, Bonilla Fernández JA, Cárdenes León A, Ramírez Burgos L, Caballero Dorta E. [Acute myocarditis after administration of the BNT162b2 vaccine against COVID-19]. Rev Esp Cardiol. 2021;74(9):812–4.

2. Reuters. Israel examining heart inflammation cases in people who received Pfizer COVID shot. 2021 Oct 28, 2021. Available from: https://www.reuters.com/world/middle-east/israel-examining-heart-inflammation-cases-people-who-received-pfizer-covid-shot-2021-04-25/.

3. Marshall M, Ferguson ID, Lewis P, Jaggi P, Gagliardo C, Collins JS, et al. Symptomatic acute myocarditis in 7 aAdolescents after Pfizer-BioNTech COVID-19 vaccination. Pediatrics. 2021;148(3).

4. Israel Ministry of Health. Press Release: Surveillance of Myocarditis (Inflammation of the Heart Muscle) Cases Between December 2020 and May 2021 (Including) 2021 Oct 28, 2021. Available from: https://www.gov.il/en/departments/news/01062021-03.

5. Willame C SM, Weibel, D.. ACCESS Background Rates of Adverse Events of Special Interest for Monitoring COVID-19 Vaccine 2021 Oct 28, 2021. Available from: http://www.encepp.eu/documents/DraftReport.pdf.

6. Abara WE, Gee J, Mu Y, Deloray M, Ye T, Shay DK, et al. Expected rates of select aAdverse eEvents following immunization for COVID-19 vaccine safety monitoring. MedRxiv. 2021:2021.08.31.21262919.

7. Kytö V, Sipilä J, Rautava P. The effects of gender and age on occurrence of clinically suspected myocarditis in adulthood. Heart. 2013;99(22):1681–4.

8. Bozkurt B, Kamat I, Hotez PJ. Myocarditis With COVID-19 mRNA Vaccines. Circulation. 2021;144(6):471–84.

9. Gargano JW, Wallace M, Hadler SC, Langley G, Su JR, Oster ME, et al. Use of mRNA COVID-19 vaccine after reports of myocarditis among vaccine recipients: Update from the Advisory Committee on Immunization Practices - United States, June 2021. MMWR Morb Mortal Wkly Rep. 2021;70(27):977–82.

10. Cooper LT. Myocarditis. N Engl J Med. 2009;360(15):1526–38.

11. Fung G, Luo H, Qiu Y, Yang D, McManus B. Myocarditis. Circ Res. 2016;118(3):496–514.

12. Chiabrando JG, Bonaventura A, Vecchié A, Wohlford GF, Mauro AG, Jordan JH, et al. Management of acute and recurrent pericarditis: JACC State-of-the-Art Review. J Am Coll Cardiol. 2020;75(1):76–92.

13. Imazio M, Brucato A, Barbieri A, Ferroni F, Maestroni S, Ligabue G, et al. Good prognosis for pericarditis with and without myocardial involvement: results from a multicenter, prospective cohort study. Circulation. 2013;128(1):42–9.

14. Shea BJ, Reeves BC, Wells G, et al. AMSTAR 2: a critical appraisal tool for systematic reviews that include randomised or non-randomised studies of healthcare interventions, or both. BMJ 2017;358:j4008. doi: 10.1136/bmj.j4008

15. Moher D, Shamseer L, Clarke M, et al. Preferred reporting items for systematic review and meta-analysis protocols (PRISMA-P) 2015 statement. Syst 2015;4(1):1. doi: 10.1186/2046-4053-4-1

16. McGowan J, Sampson M, Salzwedel DM, et al. PRESS Peer Review of Electronic Search Strategies: 2015 Guideline Statement. J Clin Epidemiol 2016;75:40-6. doi: 10.1016/j.jclinepi.2016.01.021 [published Online First: 2016/03/24]

17. Hamel C, Kelly SE, Thavorn K, Rice DB, Wells GA, Hutton B. An evaluation of DistillerSR’s machine learning-based prioritization tool for title/abstract screening – impact on reviewer-relevant outcomes. BMC Medical Research Methodology. 2020;20(1):256.

18. Sterne JAC, Savović J, Page MJ, Elbers RG, Blencowe NS, Boutron I, et al. RoB 2: a revised tool for assessing risk of bias in randomised trials. BMJ. 2019;366:l4898.

19. Joanna Briggs Institute (JBI). Checklist for Cohort Studies 2017. Available from: https://jbi.global/sites/default/files/2019-05/JBI_Critical_Appraisal-Checklist_for_Cohort_Studies2017_0.pdf.

20. Guyatt GH, Oxman AD, Schunemann HJ, Tugwell P, Knottnerus A. GRADE guidelines: a new series of articles in the Journal of Clinical Epidemiology. J Clin Epidemiol. 2011;64(4):380–2.

21. Hultcrantz M, Rind D, Akl EA, Treweek S, Mustafa RA, Iorio A, et al. The GRADE Working Group clarifies the construct of certainty of evidence. J Clin Epidemiol. 2017;87:4–13.

22. Ali K, Berman G, Zhou H, Deng W, Faughnan V, Coronado-Voges M, et al. Evaluation of mRNA-1273 SARS-CoV-2 vaccine in adolescents. N Engl J Med. 2021;11:11.

23. Baden LR, El Sahly HM, Essink B, Kotloff K, Frey S, Novak R, et al. Efficacy and safety of the mRNA-1273 SARS-CoV-2 vaccine. N Engl J Med. 2021;384(5):403–16.

24. Falsey AR, Sobieszczyk ME, Hirsch I, Sproule S, Robb ML, Corey L, et al. Phase 3 safety and efficacy of AZD1222 (ChAdOx1 nCoV-19) Covid-19 vaccine. N Engl J Med. 2021;29:29.

25. Frenck RW, Jr., Klein NP, Kitchin N, Gurtman A, Absalon J, Lockhart S, et al. Safety, immunogenicity, and efficacy of the BNT162b2 Covid-19 vaccine in adolescents. N Engl J Med. 2021;385(3):239–50.

26. Polack FP, Thomas SJ, Kitchin N, Absalon J, Gurtman A, Lockhart S, et al. Safety and efficacy of the BNT162b2 mRNA Covid-19 vaccine. N Engl J Med. 2020;383(27):2603–15.

27. Sadoff J, Gray G, Vandebosch A, Cardenas V, Shukarev G, Grinsztejn B, et al. Safety and efficacy of single-dose Ad26.COV2.S vaccine against Covid-19. N Engl J Med. 2021;384(23):2187–201.

28. Voysey M, Clemens SAC, Madhi SA, Weckx LY, Folegatti PM, Aley PK, et al. Safety and efficacy of the ChAdOx1 nCoV-19 vaccine (AZD1222) against SARS-CoV-2: an interim analysis of four randomised controlled trials in Brazil, South Africa, and the UK. Lancet. 2021;397(10269):99–111.

29. El Sahly HM, Baden LR, Essink B, Doblecki-Lewis S, Martin JM, Anderson EJ, et al. Efficacy of the mRNA-1273 SARS-CoV-2 vaccine at completion of blinded phase. N Engl J Med. 2021.

30. Thomas SJ, Moreira ED, Jr., Kitchin N, Absalon J, Gurtman A, Lockhart S, et al. Safety and efficacy of the BNT162b2 mRNA Covid-19 vaccine through 6 months. N Engl J Med. 2021;15:15.

31. Voysey M, Costa Clemens SA, Madhi SA, Weckx LY, Folegatti PM, Aley PK, et al. Single-dose administration and the influence of the timing of the booster dose on immunogenicity and efficacy of ChAdOx1 nCoV-19 (AZD1222) vaccine: a pooled analysis of four randomised trials. Lancet. 2021;397(10277):881–91.

32. European Medicines Agency. EudraVigilance - European database of suspected adverse drug reaction reports Amsterdam, the Netherlands: European Medicines Agency; 2021 [Available from: https://www.adrreports.eu/en/eudravigilance.html.

33. Israel Ministry of Health. Surveillance of Mycocarditis (Inflammation of the Heart Muscle) Cases Between December 2020 and May 2021 (Including) Israel 2021 [updated June 2, 2021. Available from: https://www.gov.il/en/departments/news/01062021-03.

34. Høeg TB, Krug A, Stevenson J, Mandrola J. SARS-CoV-2 mRNA vaccination-associated myocarditis in children ages 12-17: a stratified national database analysis. 2021. MedRxiv. https://dx.doi.org/10.1101/2021.08.30.21262866

35. Lane S, Shakir S. Reports of myocarditis and pericarditis following mRNA COVID-19 vaccines: A review of spontaneously reported data from the UK, Europe, and the U. 2021. MedRxiv. https://dx.doi.org/10.1101/2021.09.09.21263342

36. Lazaros G, Anastassopoulou C, Hatziantoniou S, Kalos T, Soulaidopoulos S, Lazarou E, et al. A case series of acute pericarditis following COVID-19 vaccination in the context of recent reports from Europe and the United States. Vaccine. 2021; 39:6585–6590.

37. Medicine & Healthcare Products Regulatory Agency. Coronavirus vaccine - weekly summary of Yellow Card reporting London: Government of the United Kingdom; 2021 [updated October 21, 2021. Available from: https://www.gov.uk/government/publications/coronavirus-covid-19-vaccine-adverse-reactions/coronavirus-vaccine-summary-of-yellow-card-reporting.

38. Public Health Agency of Canada. Canadian COVID-19 vaccination safety report. Ottawa: Public Health Agency of Canada; 2021 October 18, 2021. Available from: https://health-infobase.canada.ca/covid-19/vaccine-safety/#a4.

39. Rosenblum HG, Hadler SC, Moulia D, Shimabukuro TT, Su JR, Tepper NK, et al. Use of COVID-19 vaccines after reports of adverse events among adult recipients of Janssen (Johnson & Johnson) and mRNA COVID-19 vaccines (Pfizer-BioNTech and Moderna): Update from the Advisory Committee on Immunization Practices - United States, July 2021. MMWR Morb Mortal Wkly Rep. 2021;70(32):1094–9.

40. Su JR. Myopericarditis following COVID-19 vaccination: Updates from the Vaccine Adverse Event Reporting System (VAERS). Powerpoint Presentation. CDC; 2021 October 21, 2021. Availabe at: https://www.cdc.gov/vaccines/acip/meetings/downloads/slides-2021-10-20-21/07-COVID-Su-508.pdf

41. Barda N, Dagan N, Ben-Shlomo Y, Kepten E, Waxman J, Ohana R, et al. Safety of the BNT162b2 mRNA Covid-19 vaccine in a nationwide setting. N Engl J Med. 2021;385(12):1078–90.

42. Bardenheier BH, Gravenstein S, Blackman C, Gutman R, Sarkar IN, Feifer RA, et al. Adverse events following one dose of mRNA COVID-19 vaccination among US nursing home residents with and without a previous SARS-CoV-2 infection. J Am Med Dir Assoc. 2021;28:28.

43. Bardenheier BH, Gravenstein S, Blackman C, Gutman R, Sarkar IN, Feifer RA, et al. Adverse events following mRNA SARS-CoV-2 vaccination among U.S. nursing home residents. Vaccine. 2021;39(29):3844–51.

44. Diaz GA, Parsons GT, Gering SK, Meier AR, Hutchinson IV, Robicsek A. Myocarditis and pericarditis after vaccination for COVID-19. JAMA. 2021;326(12):1210–2.

45. Klein NP. Myocarditis Analysis in the Vaccine Safety Datalink: Rapid Cycle Analyses and “Head-to-Head” Product Comparisons. 2021 21 October 2021. Available at: https://www.cdc.gov/vaccines/acip/meetings/downloads/slides-2021-10-20-21/08-COVID-Klein-508.pdf

46. Klein NP. Rapid Cycle Analysis to Monitor the Safety of COVID-19 Vaccines in Near Real-Time within the Vaccine Safety Datalink: Myocarditis and Anaphylaxis. Aug 30 Advisory Committee on Immunization Practices (ACIP) Meeting presentation. 2021. Available from: https://www.cdc.gov/vaccines/acip/meetings/downloads/slides-2021-08-30/04-COVID-Klein-508.pdf

47. Klein NP, Lewis N, Goddard K, Fireman B, Zerbo O, Hanson KE, et al. Surveillance for adverse events after COVID-19 mRNA vaccination. JAMA. 2021;03:03.

48. Levin D, Shimon G, Fadlon-Derai M, Gershovitz L, Shovali A, Sebbag A, et al. Myocarditis following COVID-19 vaccination - A case series. Vaccine. 2021;39(42):6195–200.

49. Mevorach D, Anis E, Cedar N, Bromberg M, Haas EJ, Nadir E, et al. Myocarditis after BNT162b2 mRNA vaccine against Covid-19 in Israel. N Engl J Med. 2021. DOI: 10.1056/NEJMoa2109730

50. Montgomery J, Ryan M, Engler R, Hoffman D, McClenathan B, Collins L, et al. Myocarditis following immunization with mRNA COVID-19 vaccines in members of the US Military. JAMA Cardiol. 2021;29:29.

51. Simone A, Herald J, Chen A, Gulati N, Shen AY, Lewin B, et al. Acute myocarditis following COVID-19 mRNA vaccination in adults aged 18 years or oOlder. JAMA Intern Med. 2021;04:04.

52. Witberg G, Barda N, Hoss S, Richter I, Wiessman M, Aviv Y, et al. Myocarditis after Covid-19 vaccination in a large health care rrganization. N Engl J Med. 2021. DOI: 10.1056/NEJMoa2110737

53. Dickey JB, Albert E, Badr M, Laraja KM, Sena LM, Gerson DS, et al. A series of patients with myocarditis following SARS-CoV-2 vaccination with mRNA-1279 and BNT162b2. JACC Cardiovasc Imaging. 2021;14(9):1862–3.

54. Larson KF, Ammirati E, Adler ED, Cooper LT, Hong KN, Saponara G, et al. Myocarditis after BNT162b2 and mRNA-1273 vaccination. Circulation. 2021;144(6):506–8.

55. Rosner CM, Genovese L, Tehrani BN, Atkins M, Bakhshi H, Chaudhri S, et al. Myocarditis temporally associated with COVID-19 vaccination. Circulation. 2021;144(6):502–5.

56. Schauer J, Buddhe S, Colyer J, Sagiv E, Law Y, Chikkabyrappa SM, et al. Myopericarditis after the Pfizer mRNA COVID-19 vaccine in adolescents. J Pediatr. 2021;03:03.

57. Singer ME, Taub IB, Kaelber DC. Risk of myocarditis from COVID-19 infection in people under age 20: a population-based analysis. medRxiv 2021 doi: 10.1101/2021.07.23.21260998

58. Boehmer TK, Kompaniyets L, Lavery AM, et al. Association between COVID-19 and myocarditis using hospital-based administrative data - United States, March 2020-January 2021. MMWR Morb Mortal Wkly Rep 2021;70(35):1228–32. doi: 10.15585/mmwr.mm7035e5

