## Appendices for "Myocarditis and Pericarditis following COVID-19 Vaccination: Rapid Systematic Review of Incidence, Risk Factors, and Clinical Course"

**Appendix 1. Search strategies**
Ovid Multifile

Database: Ovid MEDLINE(R) and Epub Ahead of Print, In-Process, In-Data-Review & Other Non-Indexed Citations and Daily <1946 to October 05, 2021>, Embase <1974 to 2021 October 05>
Search Strategy:
--------------------------------------------------------------------------------
1 COVID-19 Vaccines/ [COVID VACCINES PT 1] (11038)
2 COVID-19/ (152175)
3 SARS-CoV-2/ (101136)
4 Coronavirus/ (13257)
5 Betacoronavirus/ (40795)
6 Coronavirus Infections/ (56640)
7 (COVID-19 or COVID19).tw,kf. (324030)
8 ((coronavirus* or corona virus*) and (hubei or wuhan or beijing or shanghai)).tw,kf. (10884)
9 (wuhan adj5 virus*).tw,kf. (569)
10 (2019-nCoV or 19nCoV or 2019nCoV).tw,kf. (3694)
11 (nCoV or n-CoV or "CoV 2" or CoV2).tw,kf. (124619)
12 (SARS-CoV-2 or SARS-CoV2 or SARSCoV-2 or SARSCoV2 or SARS2 or SARS-2 or severe acute respiratory syndrome coronavirus 2).tw,kf. (126706)
13 (2019-novel CoV or Sars-coronavirus2 or Sars-coronavirus-2 or SARS-like coronavirus* or ((novel or new or nouveau) adj2 (CoV or nCoV or covid or coronavirus* or corona virus or Pandemi*2)) or (coronavirus* and pneumonia)).tw,kf. (41877)
14 (novel coronavirus* or novel corona virus* or novel CoV).tw,kf. (20703)
15 ((coronavirus* or corona virus*) adj2 "2019").tw,kf. (71612)
16 ((coronavirus* or corona virus*) adj2 "19").tw,kf. (11500)
17 ("coronavirus 2" or "corona virus 2").tw,kf. (38199)
18 (OC43 or NL63 or 229E or HKU1 or HCoV* or Sars-coronavirus*).tw,kf. (8144)
19 COVID-19.rx,px,ox. or severe acute respiratory syndrome coronavirus 2.os. (9120)
20 (coronavirus* or corona virus*).ti. (48015)
21 COVID.ti. (254131)
22 ("B.1.1.7" or "B.1.351" or "B.1.617" or "B.1.427" or "B.1.429").tw,kf,rx,px,ox. (1406)
23 ("P.1" and (Brazil* or variant?)).tw,kf,rx,px,ox. (3731)
24 (((alpha or beta or delta or eta or gamma or iota or kappa or lambda) adj3 variant?) and (coronavirus* or corona virus* or covid*)).tw,kf. (459)
25 or/2-24 [COVID-19] (397323)
26 exp Vaccination/ (278048)
27 ((COVID or COVID-19 or COVID19) adj5 (immunis* or immuniz* or inoculat* or vaccin*)).tw,kf. (15689)
28 ((coronavirus* or corona virus*) adj5 (immunis* or immuniz* or inoculat* or vaccin*)).tw,kf. (3065)

29 ((2019-nCoV or nCoV or n-CoV or SARS-CoV-2 or SARS-CoV2 or SARSCoV-2 or SARSCoV2 or SARS2 or SARS-2 or OC43 or NL63 or 229E or HKU1 or HCoV*) adj5 (immunis* or immuniz* or inoculat* or vaccin*)).tw,kf. (8201)
30 (BNT162 or BNT162-01 or BNT162a1 or BNT162b1 or BNT162b2 or BNT162c2 or N38TVC63NU).tw,kf. (1584)
31 (AZD1222 or ChAdOx1 or Covishield$2 or B5S3K2V0G8).tw,kf. (967)
32 (mRNA-1273 or EPK39PL4R4).tw,kf. (596)
33 (mRNA adj5 (immunis* or immuniz* or inoculat* or vaccin*)).tw,kf. (4901)
34 (messenger RNA adj5 (immunis* or immuniz* or inoculat* or vaccin*)).tw,kf. (462)
35 LV-SMENP-DC.tw,kf. (6)
36 (Ad5-nCoV or hAdOx1 nCoV-19).tw,kf. (39)
37 ("Ad26.COV2.S" or Ad26COVS1 or JNJ 78436735 or JNJ-78436735 or JT2NS6183B).tw,kf. (195)
38 (Janssen adj5 (immunis* or immuniz* or inoculat* or vaccin*)).tw,kf. (142)
39 CX-024414.tw,kf. (2)
40 (Moderna adj5 (immunis* or immuniz* or inoculat* or vaccin*)).tw,kf. (470)
41 Spikevax$2.tw,kf. (11)
42 ((Pfizer or Pfizer-BioNTech) adj5 (immunis* or immuniz* or inoculat* or vaccin*)).tw,kf. (1249)
43 (Comirnaty$2 or Tozinameran$2).tw,kf. (202)
44 ((AstraZeneca or AZ or Oxford) adj5 (immunis* or immuniz* or inoculat* or vaccin*)).tw,kf. (731)
45 Vaxzevria$2.tw,kf. (119)
46 or/26-45 [VACCINES] (295717)
47 25 and 46 [COVID-VACCINES PT 2] (26788)
48 1 or 47 [COVID-VACCINES PTS 1-2] (28932)
49 COVID-19 Vaccines/ae [adverse events] (933)
50 Viral Vaccines/ae [adverse events] (2911)
51 Adverse Drug Reaction Reporting Systems/ (10804)
52 ((immunis* or immuniz* or inoculat* or vaccin*) adj10 (adverse* or ADE or ADEs or ADR or ADRs or complication* or harm* or safe or safety or side effect? or undesirable effect? or undesirable consequence? or undesirable outcome? or unintended effect? or unintended consequence? or unintended outcome?)).tw,kf. (76675)
53 ((post-vaccin* or post-immuni#ation* or post-innoculat* or after vaccin* or after immuni#ation* or after innoculat* or (following adj3 vaccin*) or (following adj3 immuni#ation*) or (following adj3 innoculat*)) adj10 (adverse* or ADE or ADEs or ADR or ADRs or complication* or harm* or safe or safety or side effect? or undesirable effect? or undesirable consequence? or undesirable outcome? or unintended effect? or unintended consequence? or unintended outcome?)).tw,kf. (6413)
54 ((post-vaccin* or post-immuni#ation* or post-innoculat* or after vaccin* or after immuni#ation* or after innoculat* or (following adj3 vaccin*) or (following adj3 immuni#ation*) or (following adj3 innoculat*)) adj10 (complication? or consequenc* or effect? or event? or harm* or outcome?)).tw,kf. (7716)
55 (vaccine-associated or vaccine-induced or vaccine-related).tw,kf. (20509)
56 (immuni#ation-associated or immuni#ation-induced or immuni#ation-related).tw,kf. (1855)
57 ((data or vaccin*) adj3 (monitor* or surveillance*)).tw,kf. (73396)
58 Pharmacovigilance/ (5248)
59 (pharmacovigilan* or pharmaco-vigilan*).tw,kf. (18894)
60 (Canada Vigilance or Eudravigilance* or FAERS or VAERS).tw,kf. (2780)
61 Product Surveillance, Postmarketing/ (19287)
62 ((postmarket* or post-market*) adj3 surveillance*).tw,kf. (9589)
63 "Clinical Trial, Phase IV".pt. (2193)
64 ((Phase 4 or Phase IV) adj3 (evaluation? or study or studies or trial?)).tw,kf. (4910)
65 Cardiomyopathies/ (54974)
66 (cardiomyopath* or cardio-myopath* or myocardiopath* or myo-cardiopath*).tw,kf. (209222)

67 Myocarditis/ (42633)

68 myocarditis.tw,kf. (41845)

69 ((myocard* or myo-card) adj3 inflam*).tw,kf. (10214)

70 carditis.tw,kf. (4148)

71 exp Pericarditis/ (31618)

72 pericarditis.tw,kf. (27519)

73 ((pericard* or peri-card*) adj3 inflam*).tw,kf. (1118)

74 epicarditis.tw,kf. (249)

75 ((epicard* or epi-card*) adj3 inflam*).tw,kf. (260)

76 (myopericarditis or myo-pericarditis).tw,kf. (1763)

77 ((myopericard* or myo-pericard*) adj inflam*).tw,kf. (13)

78 (pleuropericarditis or pleuro-pericarditis).tw,kf. (412)

79 ((pleuropericard* or pleuropericard*) adj3 inflam*).tw,kf. (12)

80 or/49-79 [AEs, MYOCARDITIS, PERICARDITIS] (535683)

81 48 and 80 [COVID-VACCINES - AEs, MYOCARDITIS, PERICARDITIS] (6406)

82 exp Animals/ not Humans/ (16924647)

83 81 not 82 [ANIMAL-ONLY REMOVED] (4653)

84 (comment or editorial or news or newspaper article or (letter not (letter and randomized controlled trial))).pt. (4104337)

85 83 not 84 [OPINION PIECES REMOVED] (4141)

86 limit 85 to yr="2020-current" (4046)

87 (202010* or 202011* or 202012* or 2021*).dt. (1600708)

88 86 and 87 [RECORDS SINCE 1 OCT 2020] (2589)

89 88 use ppez [MEDLINE RECORDS] (2589)

90 SARS-CoV-2 vaccine/ [COVID VACCINES - PT 1] (11038)

91 coronavirus disease 2019/ (261337)

92 severe acute respiratory syndrome coronavirus 2/ (130655)

93 Coronavirinae/ (5218)

94 Betacoronavirus/ (40795)

95 coronavirus infection/ (57662)

96 (COVID-19 or COVID19).tw,kw,kf. (324030)

97 ((coronavirus* or corona virus*) and (hubei or wuhan or beijing or shanghai)).tw,kw,kf. (10884)

98 (wuhan adj5 virus*).tw,kw,kf. (584)

99 (2019-nCoV or 19nCoV or 2019nCoV).tw,kw,kf. (3694)

100 (nCoV or n-CoV or "CoV 2" or CoV2).tw,kw,kf. (124619)

101 (SARS-CoV-2 or SARS-CoV2 or SARSCoV-2 or SARSCoV2 or SARS2 or SARS-2 or severe acute respiratory syndrome coronavirus 2).tw,kw,kf. (126706)

102 (2019-novel CoV or Sars-coronavirus2 or Sars-coronavirus-2 or SARS-like coronavirus* or ((novel or new or nouveau) adj2 (CoV or nCoV or covid or coronavirus* or corona virus or Pandemi*2)) or (coronavirus* and pneumonia)).tw,kw,kf. (41884)

103 (novel coronavirus* or novel corona virus* or novel CoV).tw,kw,kf. (20703)

104 ((coronavirus* or corona virus*) adj2 "2019").tw,kw,kf. (71614)

105 ((coronavirus* or corona virus*) adj2 "19").tw,kw,kf. (11502)

106 ("coronavirus 2" or "corona virus 2").tw,kw,kf. (38199)

107 (OC43 or NL63 or 229E or HKU1 or HCoV* or Sars-coronavirus*).tw,kw,kf. (8144)

108 (coronavirus* or corona virus*).ti. (48015)

109 COVID.ti. (254131)

110 ("B.1.1.7" or "B.1.351" or "B.1.617" or "B.1.427" or "B.1.429").tw,kw,kf. (1406)

111 ("P.1" and (Brazil* or variant?)).tw,kw,kf. (3704)

112 (((alpha or beta or delta or eta or gamma or iota or kappa or lambda) adj3 variant?) and (coronavirus* or corona virus* or covid*)).tw,kw,kf. (461)

113 or/91-112 [COVID-19] (404957)
114 vaccination/ (246289)
115 ((COVID or COVID-19 or COVID19) adj5 (immunis* or immuniz* or inoculat* or vaccin*)).tw,kw,kf. (17544)
116 ((coronavirus* or corona virus*) adj5 (immunis* or immuniz* or inoculat* or vaccin*)).tw,kw,kf. (3913)
117 ((2019-nCoV or nCoV or n-CoV or SARS-CoV-2 or SARS-CoV2 or SARSCoV-2 or SARSCoV2 or SARS2 or SARS-2 or OC43 or NL63 or 229E or HKU1 or HCoV*) adj5 (immunis* or immuniz* or inoculat* or vaccin*)).tw,kw,kf. (9069)
118 (BNT162 or BNT162-01 or BNT162a1 or BNT162b1 or BNT162b2 or BNT162c2 or N38TVC63NU).tw,kw,kf. (1584)
119 (AZD1222 or ChAdOx1 or Covishield$2 or B5S3K2V0G8).tw,kw,kf. (967)
120 (mRNA-1273 or EPK39PL4R4).tw,kw,kf. (596)
121 (mRNA adj5 (immunis* or immuniz* or inoculat* or vaccin*)).tw,kw,kf. (4944)
122 (messenger RNA adj5 (immunis* or immuniz* or inoculat* or vaccin*)).tw,kw,kf. (475)
123 LV-SMENP-DC.tw,kw,kf. (6)
124 (Ad5-nCoV or hAdOx1 nCoV-19).tw,kw,kf. (39)
125 ("Ad26.COV2.S" or Ad26COVS1 or JNJ 78436735 or JNJ-78436735 or JT2NS6183B).tw,kw,kf. (195)
126 (Janssen adj5 (immunis* or immuniz* or inoculat* or vaccin*)).tw,kf. (142)
127 CX-024414.tw,kw,kf. (2)
128 (Moderna adj5 (immunis* or immuniz* or inoculat* or vaccin*)).tw,kw,kf. (483)
129 Spikevax$2.tw,kw,kf. (11)
130 ((Pfizer or Pfizer-BioNTech) adj5 (immunis* or immuniz* or inoculat* or vaccin*)).tw,kf,kw. (1264)
131 (Comirnaty$2 or Tozinameran$2).tw,kw,kf. (202)
132 ((AstraZeneca or AZ or Oxford) adj5 (immunis* or immuniz* or inoculat* or vaccin*)).tw,kw,kf. (733)
133 Vaxzevria$2.tw,kw,kf. (119)
134 or/114-133 [VACCINES] (265426)
135 113 and 134 [COVID VACCINES - PT 2] (27369)
136 90 or 135 [COVID VACCINES - PTS 1 & 2] (29374)
137 virus vaccine/ae [adverse drug event] (999)
138 exp severe acute respiratory syndrome vaccine/ae [adverse drug event] (470)
139 ((immunis* or immuniz* or inoculat* or vaccin*) adj10 (adverse* or ADE or ADEs or ADR or ADRs or complication* or harm* or safe or safety or side effect? or undesirable effect? or undesirable consequence? or undesirable outcome? or unintended effect? or unintended consequence? or unintended outcome?)).tw,kw,kf. (76966)
140 ((post-vaccin* or post-immuni#ation* or post-innoculat* or after vaccin* or after immuni#ation* or after innoculat* or (following adj3 vaccin*) or (following adj3 immuni#ation*) or (following adj3 innoculat*)) adj10 (adverse* or ADE or ADEs or ADR or ADRs or complication* or harm* or safe or safety or side effect? or undesirable effect? or undesirable consequence? or undesirable outcome? or unintended effect? or unintended consequence? or unintended outcome?)).tw,kw,kf. (6417)
141 ((post-vaccin* or post-immuni#ation* or post-innoculat* or after vaccin* or after immuni#ation* or after innoculat* or (following adj3 vaccin*) or (following adj3 immuni#ation*) or (following adj3 innoculat*)) adj10 (complication? or consequenc* or effect? or event? or harm* or outcome?)).tw,kw,kf. (7718)
142 (vaccine-associated or vaccine-induced or vaccine-related).tw,kw,kf. (20509)
143 (immuni#ation-associated or immuni#ation-induced or immuni#ation-related).tw,kw,kf. (1855)
144 exp pharmacovigilance/ (5254)
145 (pharmacovigilan* or pharmaco-vigilan*).tw,kw,kf. (18894)
146 (Canada Vigilance or Eudravigilance* or FAERS or VAERS).tw,kw,kf. (2780)

147 exp postmarketing surveillance/ (37209)
148 ((postmarket* or post-market*) adj3 surveillance*).tw,kw,kf. (9603)
149 phase 4 clinical trial/ (4491)
150 ((Phase 4 or Phase IV) adj3 (evaluation? or study or studies or trial?)).tw,kw,kf. (4910)
151 cardiomyopathy/ (90014)
152 (cardiomyopath* or cardio-myopath* or myocardiopath* or myo-cardiopath*).tw,kw,kf. (209222)
153 exp myocarditis/ (45666)
154 myocarditis.tw,kw,kf. (41845)
155 ((myocard* or myo-card) adj3 inflam*).tw,kw,kf. (12899)
156 carditis.tw,kw,kf. (4148)
157 exp pericarditis/ (31618)
158 pericarditis.tw,kw,kf. (27519)
159 ((pericard* or peri-card*) adj3 inflam*).tw,kw,kf. (1222)
160 epicarditis.tw,kw,kf. (249)
161 ((epicard* or epi-card*) adj3 inflam*).tw,kw,kf. (391)
162 (myopericarditis or myo-pericarditis).tw,kw,kf. (1763)
163 ((myopericard* or myo-pericard*) adj inflam*).tw,kw,kf. (25)
164 (pleuropericarditis or pleuro-pericarditis).tw,kw,kf. (412)
165 ((pleuropericard* or pleuropericard*) adj3 inflam*).tw,kw,kf. (13)
166 or/137-165 [MYOCARDITIS, PERICARDITIS] (483566)
167 136 and 166 [COVID-VACCINES - MYOCARDITIS, PERICARDITIS] (6014)
168 exp animal/ or exp animal experimentation/ or exp animal model/ or exp animal experiment/ or nonhuman/ or exp vertebrate/ (54214081)
169 exp human/ or exp human experimentation/ or exp human experiment/ (42550990)
170 168 not 169 (11664830)
171 167 not 170 [ANIMAL-ONLY REMOVED] (5784)
172 editorial.pt. (1287357)
173 letter.pt. not (letter.pt. and randomized controlled trial/) (2336030)
174 171 not (172 or 173) [OPINION PIECES REMOVED] (5259)
175 limit 174 to yr="2020-current" (5170)
176 (202010* or 202011* or 202012* or 2021*).dc. (2457999)
177 175 and 176 [RECORDS PUBLISHED/ADDED SINCE OCT 2020] (2500)
178 177 use oemezd [EMBASE RECORDS] (2500)
179 89 or 178 [BOTH DATABASES] (5089)
180 remove duplicates from 179 (3235) [TOTAL UNIQUE RECORDS]
181 180 use ppez [MEDLINE UNIQUE RECORDS] (2530)
182 180 use oemezd [EMBASE UNIQUE RECORDS] (705)

***************************

### Cochrane Library

| | | | |
|---|---|---|---|
| #1 | [mh "COVID-19 Vaccines"] | Limits | 55 |
| #2 | [mh "COVID-19"] | Limits | 657 |
| #3 | [mh "SARS-CoV-2"] | Limits | 479 |
| #4 | [mh ^Coronavirus] | Limits | 4 |
| #5 | [mh ^Betacoronavirus] | Limits | 127 |
| #6 | [mh ^"Coronavirus Infections"] | Limits | 657 |
| #7 | ("COVID-19" or COVID19):ti,ab,kw | Limits | 7299 |
| #8 | ((coronavirus* or corona virus*) and (hubei or wuhan or beijing or shanghai)):ti,ab,kw | Limits | 219 |
| #9 | (wuhan NEAR/5 virus*):ti,ab,kw | Limits | 9 |
| #10 | ("2019-nCoV" or 19nCoV or 2019nCoV):ti,ab,kw | Limits | 10 |
| #11 | (nCoV or "n-CoV" or "CoV 2" or CoV2):ti,ab,kw | Limits | 690 |
| #12 | ("SARS-CoV-2" or "SARS-CoV2" or "SARSCoV-2" or SARSCoV2 or SARS2 or "SARS-2" or "severe acute respiratory syndrome coronavirus 2"):ti,ab,kw | Limits | 3039 |
| #13 | ("2019-novel CoV" or "Sars-coronavirus2" or "Sars-coronavirus-2" or "SARS-like coronavirus" or "SARS-like coronaviruses" or ((novel or new or nouveau) NEAR/2 (CoV or nCoV or covid or coronavirus* or "corona virus" or Pandemi*2)) or (coronavirus* and pneumonia)):ti,ab,kw | Limits | 1367 |
| #14 | ("novel coronavirus" or "novel coronaviruses" or "novel corona virus" or "novel corona viruses" or "novel CoV"):ti,ab,kw | Limits | 478 |
| #15 | ((coronavirus* or "corona virus" or "corona viruses") NEAR/2 "2019"):ti,ab,kw | Limits | 2503 |
| #16 | ((coronavirus* or "corona virus" or "corona viruses") NEAR/2 "19"):ti,ab,kw | Limits | 299 |
| #17 | ("coronavirus 2" or "corona virus 2"):ti,ab,kw | Limits | 765 |
| #18 | (OC43 or NL63 or 229E or HKU1 or HCoV* or (Sars NEXT coronavirus*)):ti,ab,kw | Limits | 84 |
| #19 | (coronavirus* or "corona virus" or "corona viruses"):ti | Limits | 736 |
| #20 | COVID:ti | Limits | 5641 |

| | | | |
|---|---|---|---|
| #21 | ("B.1.1.7" or "B.1.351" or "B.1.617" or "B.1.427" or "B.1.429"):ti,ab,kw | Limits | 16 |
| #22 | ("P.1" and (Brazil* or variant*)):ti,ab,kw | Limits | 175 |
| #23 | (((alpha or beta or delta or eta or gamma or iota or kappa or lambda) NEAR/3 variant*) and (coronavirus* or corona virus* or covid*)):ti,ab,kw | Limits | 7 |
| #24 | {or #2-#23} | Limits | 7898 |
| #25 | [mh Vaccination] | Limits | 2680 |
| #26 | ((COVID or "COVID-19" or COVID19) NEAR/5 (immunis* or immuniz* or inoculat* or vaccin*)):ti,ab,kw | Limits | 493 |
| #27 | ((coronavirus* or "corona virus" or "corona viruses") NEAR/5 (immunis* or immuniz* or inoculat* or vaccin*)):ti,ab,kw | Limits | 100 |
| #28 | (("2019-nCoV" or nCoV or "n-CoV" or "SARS-CoV-2" or "SARS-CoV2" or "SARSCoV-2" or SARSCoV2 or SARS2 or "SARS-2" or OC43 or NL63 or 229E or HKU1 or HCoV*) NEAR/5 (immunis* or immuniz* or inoculat* or vaccin*)):ti,ab,kw | Limits | 221 |
| #29 | (BNT162 or "BNT162-01" or BNT162a1 or BNT162b1 or BNT162b2 or BNT162c2 or N38TVC63NU):ti,ab,kw | Limits | 38 |
| #30 | (AZD1222 or ChAdOx1 or Covishield* or B5S3K2V0G8):ti,ab,kw | Limits | 63 |
| #31 | ("mRNA-1273" or EPK39PL4R4):ti,ab,kw | Limits | 24 |
| #32 | (mRNA NEAR/5 (immunis* or immuniz* or inoculat* or vaccin*)):ti,ab,kw | Limits | 100 |
| #33 | ("messenger RNA" NEAR/5 (immunis* or immuniz* or inoculat* or vaccin*)):ti,ab,kw | Limits | 24 |
| #34 | "LV-SMENP-DC":ti,ab,kw | Limits | 0 |
| #35 | ("Ad5-nCoV" or "hAdOx1 nCoV-19"):ti,ab,kw | Limits | 0 |
| #36 | ("Ad26.COV2.S" or Ad26COVS1 or "JNJ 78436735" or "JNJ-78436735" or JT2NS6183B):ti,ab,kw | Limits | 22 |
| #37 | (Janssen NEAR/5 (immunis* or immuniz* or inoculat* or vaccin*)):ti,ab,kw | Limits | 18 |
| #38 | "CX-024414":ti,ab,kw | Limits | 2 |
| #39 | (Moderna NEAR/5 (immunis* or immuniz* or inoculat* or vaccin*)):ti,ab,kw | Limits | 20 |
| #40 | Spikevax*:ti,ab,kw | Limits | 0 |
| #41 | ((Pfizer or "Pfizer-BioNTech") NEAR/5 (immunis* or immuniz* or inoculat* or vaccin*)):ti,ab,kw | Limits | 47 |
| #42 | (Comirnaty* or Tozinameran*):ti,ab,kw | Limits | 10 |
| #43 | ((AstraZeneca or AZ or Oxford) NEAR/5 (immunis* or immuniz* or inoculat* or vaccin*)):ti,ab,kw | Limits | 34 |
| #44 | Vaxzevria*:ti,ab,kw | Limits | 6 |
| #45 | {or #25-#44} | Limits | 3419 |
| #46 | #24 AND #45 | Limits | 638 |
| #47 | #1 OR #46 | Limits | 638 |
| #48 | [mh "COVID-19 Vaccines"/AE] | Limits | 30 |
| #49 | [mh ^"Viral Vaccines"/AE] | Limits | 221 |
| #50 | [mh "Adverse Drug Reaction Reporting Systems"] | Limits | 90 |
| #51 | ((immunis* or immuniz* or inoculat* or vaccin*) NEAR/10 (adverse* or ADE or ADEs or ADR or ADRs or complication* or harm* or safe or safety or "side effect" or "side effects" or "undesirable effect" or "undesirable effects" or "undesirable consequence" or "undesirable consequences" or "undesirable outcome" or "undesirable outcomes" or "unintended effect" or "unintended effects" or "unintended consequence" or "unintended consequences" or "unintended outcome" or "unintended outcome"s)):ti,ab,kw | Limits | 12385 |
| #52 | (((post NEXT vaccin*) or (post NEXT immunisation*) or (post NEXT immunization*) or (post NEXT inoculat*) or (after NEXT vaccin*) or (after NEXT immunisation*) or (after NEXT immunization*) or (after NEXT inoculat*) or (following NEAR/3 vaccin*) or (following NEAR/3 immunisation*) or (following NEAR/3 immunization*) or (following NEAR/3 inoculat*)) NEAR/10 (adverse* or ADE or ADEs or ADR or ADRs or complication* or harm* or safe or safety or "side effect" or "side effects" or "undesirable effect" or "undesirable effects" or "undesirable consequence" or "undesirable consequences" or "undesirable outcome" or "undesirable outcomes" or "unintended effect" or "unintended effects" or "unintended consequence" or "unintended consequences" or "unintended outcome" or "unintended outcome"s)):ti,ab,kw | Limits | 1493 |
| #53 | (((post NEXT vaccin*) or (post NEXT immunisation*) or (post NEXT immunization*) or (post NEXT inoculat*) or (after NEXT vaccin*) or (after NEXT immunisation*) or (after NEXT immunization*) or (after NEXT inoculat*) or (following NEAR/3 vaccin*) or (following NEAR/3 immunisation*) or (following NEAR/3 immunization*) or (following NEAR/3 inoculat*)) NEAR/10 (complication* or consequenc* or effect or effects or event or events or harm* or outcome*)):ti,ab,kw | Limits | 1214 |
| #54 | ("vaccine-associated" or "vaccine-induced" or "vaccine-related"):ti,ab,kw | Limits | 1451 |
| #55 | ("immunisation-associated" or "immunisation-induced" or "immunisation-related" or "immunization-associated" or "immunization-induced" or "immunization-related"):ti,ab,kw | Limits | 38 |
| #56 | ((data or vaccin*) NEAR/3 (monitor* or surveillance*)):ti,ab,kw | Limits | 4798 |
| #57 | [mh Pharmacovigilance] | Limits | 18 |

| | | # | Search | | Results |
|---|---|---|---|---|---|
| − | + | #58 | (pharmacovigilan* or (pharmaco NEXT vigilan*)):ti,ab,kw | Limits | 318 |
| − | + | #59 | ("Canada Vigilance" or Eudravigilance* or FAERS or VAERS):ti,ab,kw | Limits | 19 |
| − | + | #60 | [mh ^"Product Surveillance, Postmarketing"] | Limits | 100 |
| − | + | #61 | ((postmarket* or (post NEXT market*)) NEAR/3 surveillance*):ti,ab,kw | Limits | 732 |
| − | + | #62 | "Clinical Trial, Phase IV":pt | Limits | 1201 |
| − | + | #63 | (("Phase 4" or "Phase IV") NEAR/3 (evaluation* or study or studies or trial or trials)):ti,ab,kw | Limits | 2770 |
| − | + | #64 | [mh ^Cardiomyopathies] | Limits | 677 |
| − | + | #65 | (cardiomyopath* or (cardio NEXT myopath*) or myocardiopath* or (myo NEXT cardiopath*)):ti,ab,kw | Limits | 4440 |
| − | + | #66 | [mh Myocarditis] | Limits | 93 |
| − | + | #67 | myocarditis:ti,ab,kw | Limits | 1208 |
| − | + | #68 | ((myocard* or (myo NEXT card*)) NEAR/3 inflam*):ti,ab,kw | Limits | 268 |
| − | + | #69 | carditis:ti,ab,kw | Limits | 36 |
| − | + | #70 | [mh Pericarditis] | Limits | 63 |
| − | + | #71 | pericarditis:ti,ab,kw | Limits | 349 |
| − | + | #72 | ((pericard* or (peri NEXT card*)) NEAR/3 inflam*):ti,ab,kw | Limits | 22 |
| − | + | #73 | epicarditis:ti,ab,kw | Limits | 2 |
| − | + | #74 | ((epicard* or (epi NEXT card*)) NEAR/3 inflam*):ti,ab,kw | Limits | 5 |
| − | + | #75 | (myopericarditis or "myo-pericarditis"):ti,ab,kw | Limits | 12 |
| − | + | #76 | ((myopericard* or (myo NEXT pericard*)) NEAR/3 inflam*):ti,ab,kw | Limits | 1 |
| − | + | #77 | (pleuropericarditis or "pleuro-pericarditis"):ti,ab,kw | Limits | 3 |
| − | + | #78 | ((pleuropericard* or (pleuro NEXT pericard*)) NEAR/3 inflam*):ti,ab,kw | Limits | 0 |
| − | + | #79 | {or #48-#78} | Limits | 27721 |
| − | + | #80 | #47 AND #79<br>with Cochrane Library publication date from Oct 2020 to Oct 2021 | Limits | 339 |

*6-Oct-21*

| | Original | Deduped |
|---|---|---|
| MEDLINE | 2530 | 2511 |
| Embase | 705 | 675 |
| CDSR | 2 | 0 |
| CENTRAL | 337 | 253 |
| TOTAL | 3574 | 3439 |

**Appendix 2. Literature flow**

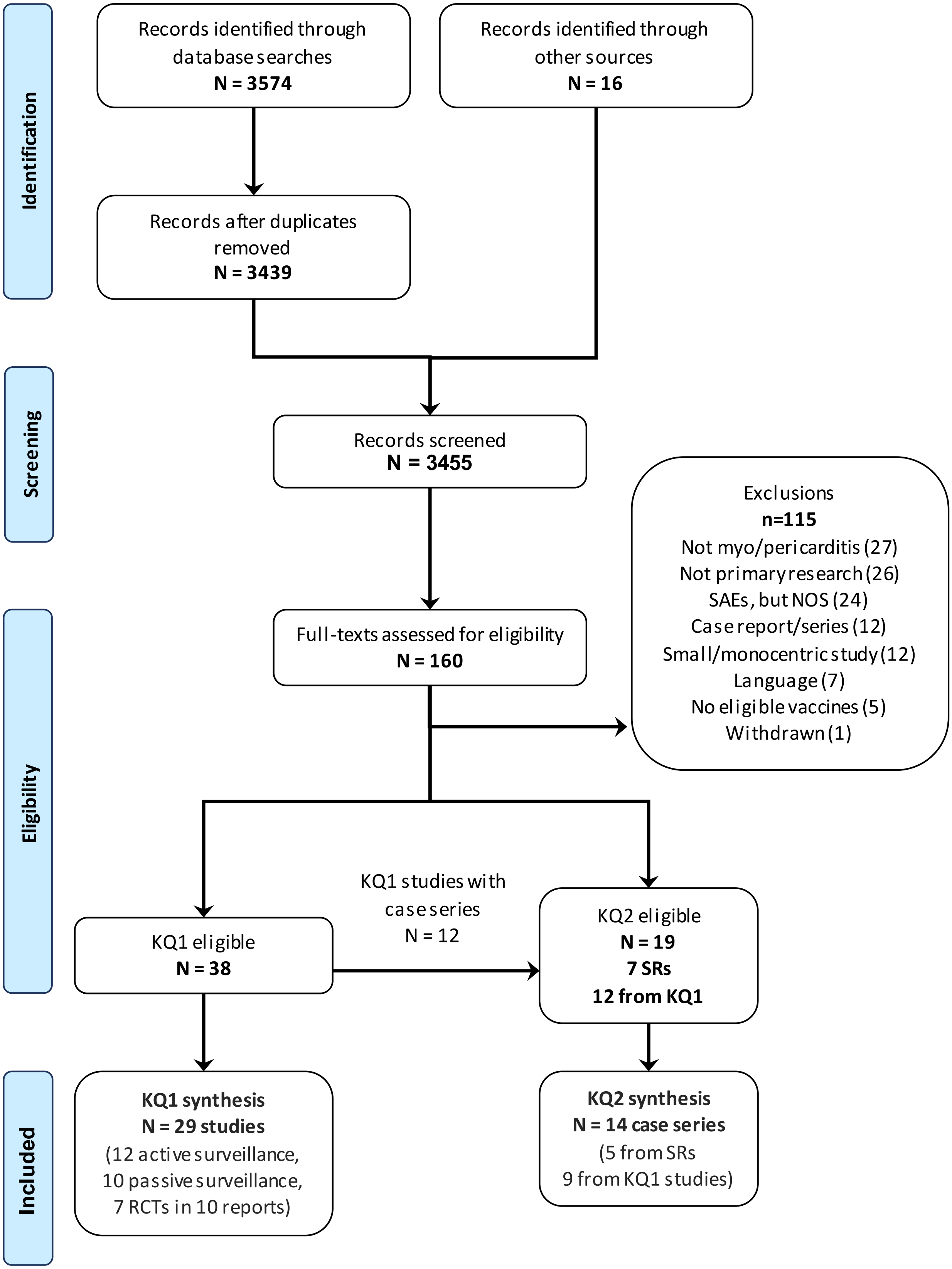

### Appendix 3. Supplements to Tables 2 and 3

#### Descriptions of surveillance systems used in studies within this report

CAEFISS: Managed by PHAC, monitors the safety of marketed vaccines in Canada through both passive and active reporting strategies. Reports are submitted by public health authorities in provinces and territories, who in turn receive them from local public health units. Provincial and territorial authorities also receive reports from federal authorities that provide immunization in their jurisdictions. Active surveillance is also carried out by 12 pediatric centres across Canada, which screen all hospital admissions for potential vaccine-related adverse events.

CVP: Passive surveillance program that collects reports of suspected adverse reactions to health products. Reports are submitted by health professionals and consumers on a voluntary basis either directly to Health Canada or to the market authorization holder, who in turn is required to submit reports of adverse events to Health Canada through the Canada Vigilance Program.

EudraVigilance: Passive surveillance system for the European Economic Area. Healthcare providers and patients can report any adverse effects from medical products, including vaccines. Patients, consumers and healthcare professionals report suspected side effects to either the national medicines regulatory authority or the pharmaceutical company that holds the marketing authorisation for the medicine. These reports are then transmitted electronically to EudraVigilance.

VAERS: Passive surveillance system for the United States, to which healthcare providers and patients can report adverse events from medical products, including vaccines. Providers are required to report to VAERS adverse events (including administration errors, serious adverse events, cases of multisystem inflammatory syndrome, and cases of COVID-19 that result in hospitalization or death) that occur after receipt of any COVID-19 vaccine.

Yellow Card: The Yellow Card scheme in the United Kingdom is a passive surveillance system to which anybody can voluntarily report any suspected adverse reactions or side effects to the vaccine. The reports are continually reviewed to detect possible new side effects that may require regulatory action, and to differentiate these from things that would have happened regardless of the vaccine or medicine being administered, for instance due to underlying or undiagnosed illness.

**References for included studies in question 1 labeled based on data sources**

| Study | Reference(s) |
|---|---|
| **Active Surveillance** | |
| Clalit Health May 24a | Witberg G, Barda N, Hoss S, et al. Myocarditis after Covid-19 Vaccination in a Large Health Care Organization. N Engl J Med. 2021. doi:https://dx.doi.org/10.1056/NEJMoa2110737. |
| Clalit Health May 24b | Barda N, Dagan N, Ben-Shlomo Y, et al. Safety of the BNT162b2 mRNA Covid-19 Vaccine in a Nationwide Setting. N Engl J Med. 2021;385(12):1078-1090. doi:https://dx.doi.org/10.1056/NEJMoa2110475. |
| Genesis Healthcare Jan 3 | Bardenheier BH, Gravenstein S, Blackman C, et al. Adverse events following mRNA SARS-CoV-2 vaccination among U.S. nursing home residents. Vaccine. 2021;39(29):3844-3851. doi:https://dx.doi.org/10.1016/j.vaccine.2021.05.088. |
| Genesis HealthcareFeb 14 | Bardenheier BH, Gravenstein S, Blackman C, et al. Adverse Events Following One Dose of mRNA COVID-19 Vaccination Among US Nursing Home Residents With and Without a Previous SARS-CoV-2 Infection. J Am Med Dir Assoc. 2021;28:28. doi:https://dx.doi.org/10.1016/j.jamda.2021.08.024. |

| Study | Reference(s) |
|---|---|
| Israel MOH May 31 | Mevorach D, Anis E, Cedar N, et al. Myocarditis after BNT162b2 mRNA Vaccine against Covid-19 in Israel. N Engl J Med. 2021. doi:https://dx.doi.org/10.1056/NEJMoa2109730. |
| Israeli Defense Forces May 7 | Levin D, Shimon G, Fadlon-Derai M, et al. Myocarditis following COVID-19 vaccination - A case series. Vaccine. 2021;39(42):6195-6200. doi:https://dx.doi.org/10.1016/j.vaccine.2021.09.004. |
| KPSC Jul 20 | Simone A, Herald J, Chen A, et al. Acute Myocarditis Following COVID-19 mRNA Vaccination in Adults Aged 18 Years or Older. JAMA Intern Med. 2021;04:04. doi:https://dx.doi.org/10.1001/jamainternmed.2021.5511. |
| Providence Health May 25 | Diaz GA, Parsons GT, Gering SK, Meier AR, Hutchinson IV, Robicsek A. Myocarditis and Pericarditis After Vaccination for COVID-19. Jama. 2021;326(12):1210-1212. doi:https://dx.doi.org/10.1001/jama.2021.13443. |
| US Military Apr 30 | Montgomery J, Ryan M, Engler R, et al. Myocarditis Following Immunization With mRNA COVID-19 Vaccines in Members of the US Military. JAMA Cardiol. 2021;29:29. doi:https://dx.doi.org/10.1001/jamacardio.2021.2833. |
| VSD Jun 26 | Klein NP, Lewis N, Goddard K, et al. Surveillance for Adverse Events After COVID-19 mRNA Vaccination. Jama. 2021;03:03. doi:https://dx.doi.org/10.1001/jama.2021.15072. |
| VSD Aug 21 | Klein NP. Rapid Cycle Analysis to Monitor the Safety of COVID-19 Vaccines in Near Real-Time within the Vaccine Safety Datalink: Myocarditis and Anaphylaxis. Aug 30 Advisory Committee on Immunization Practices (ACIP) https://www.cdc.gov/vaccines/acip/meetings/downloads/slides-2021-08-30/04-COVID-Klein-508.pdf |
| VSD Oct 6 | Klein NP. Myocarditis Analysis in the Vaccine Safety Datalink: Rapid Cycle Analyses and "Head-to-Head" Product Comparisons. 21 October 2021 2021. Advisory Committee on Immunization Practices (ACIP). Available from: https://www.cdc.gov/vaccines/acip/meetings/downloads/slides-2021-10-20-21/08-COVID-Klein-508.pdf. |
| **Passive Surveillance** | |
| CAEFISS & CVP | Public Health Agency of Canada. Canadian COVID-19 vaccination safety report. Ottawa: Public Health Agency of Canada; October 18, 2021 2021. Available from: https://health-infobase.canada.ca/covid-19/vaccine-safety/#a4. |
| EudraVigilance Oct 19 | European Medicines Agency. EudraVigilance - European database of suspected adverse drug reaction reports. European Medicines Agency. Available from: https://www.adrreports.eu/en/eudravigilance.html. Published 2021. Accessed October 26, 2021, 2021. |
| Israel MOH May 30 | Israel Ministry of Health. Surveillance of Mycocarditis (Inflammation of the Heart Muscle) Cases Between December 2020 and May 2021 (Including). Available from: https://www.gov.il/en/departments/news/01062021-03. Published 2021. Updated June 2, 2021. Accessed. |

| Study | Reference(s) |
|---|---|
| VAERS Jun 11 | Gargano JW, Wallace M, Hadler SC, et al. Use of mRNA COVID-19 Vaccine After Reports of Myocarditis Among Vaccine Recipients: Update from the Advisory Committee on Immunization Practices - United States, June 2021. MMWR Morb Mortal Wkly Rep. 2021;70(27):977-982. doi:https://dx.doi.org/10.15585/mmwr.mm7027e2. |
| VAERS Jun 18a | Høeg TB, Krug A, Stevenson J, Mandrola J. SARS-CoV-2 mRNA Vaccination-Associated Myocarditis in Children Ages 12-17: A Stratified National Database Analysis. 2021. doi:https://dx.doi.org/10.1101/2021.08.30.21262866. |
| VAERS Jun 18b | Lazaros G, Anastassopoulou C, Hatziantoniou S, et al. A case series of acute pericarditis following COVID-19 vaccination in the context of recent reports from Europe and the United States. Vaccine. 2021. doi:https://dx.doi.org/10.1016/j.vaccine.2021.09.078. |
| VAERS Jun 30 | Rosenblum HG, Hadler SC, Moulia D, et al. Use of COVID-19 Vaccines After Reports of Adverse Events Among Adult Recipients of Janssen (Johnson & Johnson) and mRNA COVID-19 Vaccines (Pfizer-BioNTech and Moderna): Update from the Advisory Committee on Immunization Practices - United States, July 2021. MMWR Morb Mortal Wkly Rep. 2021;70(32):1094-1099. doi:https://dx.doi.org/10.15585/mmwr.mm7032e4. |
| VAERS Aug 6 | Lane S, Shakir S. Reports of myocarditis and pericarditis following mRNA COVID-19 vaccines: A review of spontaneously reported data from the UK, Europe, and the U. 2021. doi:https://dx.doi.org/10.1101/2021.09.09.21263342. |
| VAERS Oct 6 | Su JR. Myopericarditis following COVID-19 vaccination: Updates from the Vaccine Adverse Event Reporting System (VAERS). CDC October 21, 2021 2021. Available from: https://www.cdc.gov/vaccines/acip/meetings/downloads/slides-2021-10-20-21/07-COVID-Su-508.pdf. |
| Yellow Card Oct 13 | Medicine & Healthcare products Regulatory Agency. Coronavirus vaccine - weekly summary of Yellow Card reporting. Government of the United Kingdom. Available from: https://www.gov.uk/government/publications/coronavirus-covid-19-vaccine-adverse-reactions/coronavirus-vaccine-summary-of-yellow-card-reporting. Published 2021. Updated October 21, 2021. Accessed October 21, 2021, 2021. |
| **Randomized Controlled Trials** | |
| AZD1222 Trial, Phase 3 (AstraZeneca) | Falsey AR, Sobieszczyk ME, Hirsch I, et al. Phase 3 Safety and Efficacy of AZD1222 (ChAdOx1 nCoV-19) Covid-19 Vaccine. N Engl J Med. 2021;29:29. doi:https://dx.doi.org/10.1056/NEJMoa2105290. |
| AZD1222 Trial, Pooled Analysis (AstraZeneca) | Voysey M, Clemens SAC, Madhi SA, et al. Safety and efficacy of the ChAdOx1 nCoV-19 vaccine (AZD1222) against SARS-CoV-2: an interim analysis of four randomised controlled trials in Brazil, South Africa, and the UK. Lancet. 2021;397(10269):99-111. doi:https://dx.doi.org/10.1016/S0140-6736(20)32661-1. |
| | Voysey M, Costa Clemens SA, Madhi SA, et al. Single-dose administration and the influence of the timing of the booster dose on immunogenicity and efficacy of ChAdOx1 nCoV-19 (AZD1222) vaccine: a pooled analysis of four randomised trials. Lancet. 2021;397(10277):881-891. doi:https://dx.doi.org/10.1016/S0140-6736(21)00432-3. |

| Study | Reference(s) |
|---|---|
| C4591001 Trial, Adolescent cohort (Pfizer) | Frenck RW, Jr., Klein NP, Kitchin N, et al. Safety, Immunogenicity, and Efficacy of the BNT162b2 Covid-19 Vaccine in Adolescents. N Engl J Med. 2021;385(3):239-250. doi:https://dx.doi.org/10.1056/NEJMoa2107456. |
| C4591001 Trial, Adult cohort (Pfizer) | Polack FP, Thomas SJ, Kitchin N, et al. Safety and Efficacy of the BNT162b2 mRNA Covid-19 Vaccine. N Engl J Med. 2020;383(27):2603-2615. doi:https://dx.doi.org/10.1056/NEJMoa2034577. |
| | Thomas SJ, Moreira ED, Jr., Kitchin N, et al. Safety and Efficacy of the BNT162b2 mRNA Covid-19 Vaccine through 6 Months. N Engl J Med. 2021;15:15. doi:https://dx.doi.org/10.1056/NEJMoa2110345. |
| COVE trial (Moderna) | Baden LR, El Sahly HM, Essink B, et al. Efficacy and Safety of the mRNA-1273 SARS-CoV-2 Vaccine. N Engl J Med. 2021;384(5):403-416. doi:https://dx.doi.org/10.1056/NEJMoa2035389. |
| | El Sahly HM, Baden LR, Essink B, et al. Efficacy of the mRNA-1273 SARS-CoV-2 Vaccine at Completion of Blinded Phase. N Engl J Med. 2021. doi:https://dx.doi.org/10.1056/NEJMoa2113017. |
| Teen COVE trial (Moderna) | Ali K, Berman G, Zhou H, et al. Evaluation of mRNA-1273 SARS-CoV-2 Vaccine in Adolescents. N Engl J Med. 2021;11:11. doi:https://dx.doi.org/10.1056/NEJMoa2109522. |
| ENSEMBLE Trial (Janssen) | Sadoff J, Gray G, Vandebosch A, et al. Safety and Efficacy of Single-Dose Ad26.COV2.S Vaccine against Covid-19. N Engl J Med. 2021;384(23):2187-2201. doi:https://dx.doi.org/10.1056/NEJMoa2101544. |

### Appendix 4: Study Characteristics Tables and Risk of Bias Tables

#### Table A1. Study characteristics of observational studies using data from active surveillance systems (question 1)

| Dataset<br>Dates<br>Country | Vaccines Studied | Sample Size; Demographics; Previous Covid-19 diagnoses | Study Group(s) | Outcome(s); Risk Interval; Case Ascertainment | Analysis | Results |
|---|---|---|---|---|---|---|
| VSD[†] Oct 9<br><br>Dec 14 2020 to Oct 9<br><br>USA<br><br>(Klein, 2021a) | Pfizer-BioNTech (60% of dose 2) or Moderna | Total doses 14,214,955 (71.5% eligible people fully vaccinated) to 7.5 million people<br><br>NR<br><br>Excluded from analysis if COVID-19 diagnosis ≤30 d before vaccination | 1. Vaccinated in previous 0-21 d<br>2. Concurrent previous vaccinees (22 to 42 d previously for similar individuals after vaccine dose 1 or 2 [most dose 2])<br><br>Interval between doses: majority 21 d (Pfizer) and 28 d Moderna) | Myocarditis/pericarditis/ myopericarditis (combined & separate for head-to-head comparison)<br><br>ICD-10 using inclusion and exclusionary codes via clinical expert consultation; cases among 12-39 y confirmed by medical record review<br><br>ED and in-patient records used<br><br>Blinding to vaccine status NR<br><br>Risk interval: 0-21 d & 0-7 d | Weekly, rapid cycle analysis<br><br>Adjusted incident rate ratio (aIRR) estimated by Poisson regression adjusted for age, sex, race and ethnicity, health plan, and calendar day; 1-sided P < .0048 for statistical signal<br><br>Excess risk/incidence per million doses (IR – [IR/IRR])<br><br><u>Exploratory analyses</u> (no prespecified level of significance): by dose and vaccine type; for 12-39 y by dose, vaccine type and with shorter risk intervals; head-to-head Moderna vs Pfizer in 18-39 y (also excluded pericarditis) | Events: 138 (not confirmed) after either dose<br>Incidence: 138/14,214,955 = 9.7 per million doses<br>aIRR: 1.72 p <.001 (statistical signal)<br><br><u>Exploratory analyses</u>:<br><br>12-39 y (all confirmed cases)<br><br>Group 1: 74 cases in 0-21 d interval (59 [80%] after dose 2; 39 [53% in 0-7 d)); 44 (59%) in 18-39 y and 30 (40.5% in 12-17 y<br><br>Clustering at 0-5 d after vaccine (p<.0001)<br><br>**0-7 d risk interval**<br>aIRR (95% CI) — Excess risk per million doses<br>Both doses 21.73 (9.53 to 57.34) 11.8<br>Dose 2 31.28 (12.83 to 91.40) 21.1<br><br>Pfizer (<u>12-39y</u>) 19.31 (7.66 to 57.79) 11.5<br>Dose 2 30.27 (11.01 to -104.4) 21.5<br><br>Moderna (<u>18-39y</u>) 37.51 (6.69 to 803.00) 12.8<br>Dose 2 NE (11.75 to NE) 21.0<br><br>(For dose 1: aIRR range 8.7-10.5, all significant; excess cases 2.3-5.2)<br><br>**0-7 d risk interval (*18-39 y only*)**<br>aIRR (95% CI) — Excess risk per million doses<br>Both doses 12.61 (5.27 to 34.47) 8.4<br>Dose 2 14.33 (5.50 to 43.88) 13.1<br><br>Pfizer 7.98 (2.72 to 26.50) 5.7<br>Dose 2 8.77 (2.56 to 35.34) 8.5<br><br>Moderna 37.42 (6.68 to 801.33) 12.8<br>Dose 2 NE (11.70 – NE) 21.0 |

| Dataset<br>Dates<br>Country | Vaccines Studied | Sample Size; Demographics; Previous Covid-19 diagnoses | Study Group(s) | Outcome(s); Risk Interval; Case Ascertainment | Analysis | Results |
|---|---|---|---|---|---|---|
| | | | | | | (For dose 1: aIRR range 8.05 to 10.47; all significant; excess cases 3.3 to 5.2)<br><br>**0-7 d risk interval (*12-17 y only)***<br>aIRR (95% CI) Excess risk per million doses<br>Pfizer NE (16.88 to NE) 29.6<br>Dose 2 NE (28.83 to NE) 54.0<br><br>**0-21 d risk interval (*12-17 y only*)**<br>aIRR (95% CI) Excess risk per million doses<br>Pfizer NE (5.68 to NE) 29.6<br>Dose 2 NE (9.09 to NE) 56.7<br><br>**Head-to-head Moderna vs Pfizer in 18-39 y**<br><br>(0-7 d) aIRR Excess risk per million doses<br>Both doses 2.56 (1.32 to 5.03) 8.0<br>Dose 2 2.72 (1.25 to 6.05) 13.3<br>Dose 2 (males) 2.26 (1.00 to 5.19) 21.5<br><br>(0-21 d) aIRR Excess risk per million doses<br>Both doses 2.05 (1.11 to 3.83) 7.1<br>Dose 2 2.19 (1.05 to 4.65) 11.5<br><br>Dose 1 all nonsignificant<br><br>**Head-to-head Moderna vs Pfizer in 18-39 y (without pericarditis)**<br>aIRR Excess cases per million doses<br>(0-7 d)<br>Both doses 2.24 (1.09 to 4.63) 5.6<br>Dose 2 2.28 (1.25 to 6.05) 13.3<br>Dose 2 (males) 2.14 (0.93 to 4.98) 19.1<br><br>(0-21 d)<br>Both doses 1.96 (**0.99** to 3.91) 5.3<br>Dose 2 2.19 (**0.98** to 4.97) 9.4<br>Dose 1 all nonsignificant |
| VSD[†] Aug 21<br><br>Dec 14 2020 to Aug 21 2021 | Pfizer-BioNTech (57% overall) or Moderna | Total doses 13,334,831 (66.5% fully vaccinated) to 7.1 million people | 1. Vaccinated in previous 0-21 d<br>2. Concurrent previous vaccinees (22 to 42 d previously for similar | Myocarditis/pericarditis/ myopericarditis (combined)<br>Risk interval: 0-21 d | | Events: 115<br>aIRR: 1.57 p=0.010 (not meeting p<0.0048 signal)<br><br><u>Subgroups</u>: |

| Dataset Dates Country | Vaccines Studied | Sample Size; Demographics; Previous Covid-19 diagnoses | Study Group(s) | Outcome(s); Risk Interval; Case Ascertainment | Analysis | Results |
|---|---|---|---|---|---|---|
| USA<br><br>(Klein 2021b) | | | individuals after vaccine dose 1 or 2 [most dose 2]) | | | *12-39 y*<br>56 cases in 0-21 d risk interval<br>aIRR / Excess risk per million doses<br>Both doses 5.63 (2.31 to 16.44) 8.9<br>Dose 2 8.31 (3.07 to 28.28) 15.5<br><br>Pfizer 3.62 (1.39 to 11.11) 7.2<br>Dose 2 5.74 (1.98 to 20.52) 13.6<br><br>Moderna NE (3.32 to NE) 12.7<br>Dose 2 NE (3.79 to NE) 19.8<br><br>Dose 1 aIRR 2.3 to 3.8 and imprecise for separate vaccines<br><br>**For 0-7 risk interval – see more recent data in Klein Oct 21, 2021** |
| VSD† Jun 26<br><br>Dec 14 2020 to Jun 26 2021<br><br>USA<br><br>Klein 2021c | Pfizer-BioNTech (57% overall) or Moderna | 10,162,227 doses to 6.2 million vaccine-eligible members (≥12 y) (60.7% received at least 1 dose)<br><br>Females 52.4%<br>Mean age 49 y; 5.5% 12-15 y; 2.8%16-17 y; 50.4% 18-49 y<br>White non-Hispanic 40.6%<br><br>Excluded from analysis if COVID-19 diagnosis ≤30 d before vaccination | 1. Vaccinated in previous 0-21 d<br>2. Concurrent previous vaccinees (22 to 42 d previously for similar individuals after vaccine dose 1 or 2 [most dose 2])<br>3. Unvaccinated concurrent comparators (same day)<br><br>Interval between doses NR | Myocarditis/pericarditis/ myopericarditis (combined)<br><br>Risk interval: 0-21 d and 0-7 d<br><br>ICD-10 using inclusion and exclusionary codes via clinical expert consultation; cases among 12-39 y confirmed by medical record review<br><br>ED and in-patient records used<br><br>Blinding to vaccine status NR | Weekly, rapid cycle analysis<br><br>Incidence after any dose, after dose 1 and after dose 2<br><br>Incident rate ratio (IRR) estimated by Poisson regression adjusted for age, sex, race and ethnicity, health plan, and calendar day; 1-sided P < .0048 for statistical signal<br><br>Excess risk/incidence per million doses (IR – [IR/IRR])<br><br><u>Exploratory analyses</u> (no prespecified level of significance): by dose and vaccine type; for 12-39 y by dose, vaccine type and with shorter risk intervals | **Group 1 vs. 2**<br>Events: 87 vs 39 events<br>Incidence: 131.1 vs. 106.9 per million person-years<br>aIRR: 1.18 (95% CI, 0.79 to 1.79)<br>Excess risk/cases per million doses: 1.2 (95% CI, –2.1 to 3.3)<br><br><u>Exploratory analyses</u>:<br><br>*By dose and vaccine type:* p > 0.0048 for all<br><br>*12-39y*:<br>34 cases (53% 12-24y; 85% male; 82% hospitalized; 71% after dose 2; 7 myocarditis; 6 pericarditis; 21 myopericarditis)<br>Clustering at 0-5 d after vaccine (p<.001)<br>During 0-21 d after vaccination (either dose):<br>Incidence: 141.2 vs. 35.0 cases per million person-years<br>aIRR: 3.74 (95% CI, 1.38 to 12.84)<br>Similar results for each dose but most precise for dose 2 having more data<br>During 0-7 d after vaccination (either dose):<br>Incidences: 321 vs 35 cases per million person-years<br>aIRR: 9.83 (95% CI, 3.35 to 35.77)<br>Excess risk per million doses: 6.3 (95% CI, 4.9 to 6.8)<br>Similar results for each dose but most precise for dose 2 having more data<br>Similar findings in analyses by vaccine type |

| Dataset<br>Dates<br>Country | Vaccines Studied | Sample Size; Demographics; Previous Covid-19 diagnoses | Study Group(s) | Outcome(s); Risk Interval; Case Ascertainment | Analysis | Results |
|---|---|---|---|---|---|---|
| | | | | | | **Group 1 vs 3:**<br>Events: 87 vs. 293<br>Incidence 132 vs. 83 cases per million person-years<br>aIRR: 1.39 (95% CI, 1.05 to 1.82) |
| US Military Apr 30<br><br>Jan 1 to Apr 30 2021<br><br>USA<br><br>Montgomery 2021 | Pfizer-BioNTech or Moderna | 2,810,00 doses (38% dose 2)<br><br>Males 100%<br>Median age 25 (20-51)<br><br>Tested cases for Covid-19 n=0 but all cases after dose 2 (n=3) had previous Covid-19 | 1. Vaccinated<br>2. Expected numbers within 30 d after vaccination | Myocarditis<br><br>Cases identified via referrals to Defense Health Agency clinical specialists and through review of VAERS reports; each cases adjudicated using CDC definition for probable<br><br>Risk interval: all presented within 4 d | Incidence in vaccinated<br><br>Observed vs expected cases: expected number based on an expected annual incidence ranging from 1-10 per 100 000 person-years (US) to 22 per 100 000 person-years (internationally); presenting within a 30-day period after vaccination. | Events: 23 (20 after dose 2)<br><br>Observed vs expected:<br>Total doses: 23 v vs 2 to 52<br>Dose 2: 20 vs 1 to 20<br>Dose 2 to military members: 19 vs 0 to 10<br>Dose 2 to male military members: 19 vs 0 to 8<br><br>Incidence:<br>Total doses: 0.8 per 100,000 doses<br>Dose 2: 1.9 per 100,000 doses<br>Dose 2 to military members: 3.5 per 100,000 doses<br>Dose 2 to male military members: 4.4 per 100,000 doses |
| Providence Health May 25<br><br>Up to May 25 2021<br><br>USA<br><br>Diaz 2021 | Pfizer, Moderna, Janssen<br><br>At least one dose (23.5% more than 1) | N=2,000,287<br><br>Female 58.9%<br>Median age 57 y (IQR 40-70) | 1. Individuals vaccinated with at least 1 dose<br>2. Unvaccinated individuals Jan 19 – Jan21 | Myocarditis, Pericarditis w/o myocarditis<br><br>Emergency department or inpatient encounters with diagnoses of myocarditis/ myopericarditis, or pericarditis<br><br>Patients with previous diagnoses (cardiac, immunological, infectious) excluded<br><br>Risk interval: 30d | Change/excess in incidence between periods | Excess incidence of myocarditis/myopericarditis: 1.0 (0.61-1.54) per 100,000 vaccinees<br>Excess incidence of pericarditis: 1.8 (1.30-2.55) per 100,000 vaccinees |
| KPSC Jul 20<br><br>Dec 14 2021 to Jul 20 2021<br><br>USA<br><br>Simone 2021 | BNT162b2 (Pfizer) or mRNA-1273 (Moderna) (93.5% fully vaccinated) | KPSC members ≥18y<br>Vaccinated group n=2,392,924<br><br>Median age 49y (IQR: 34-64y)<br>Age <40 y: 35.7%<br><br>Females 54.0% | 1. Vaccinated<br>2. Concurrent unvaccinated<br>3. Vaccinated individuals during a 10-day period 1 year prior to vaccination | Myocarditis<br><br>Reports from clinicians to the KPSC Regional Immunization Practice Committee and by identifying hospitalization within 10 days of vaccine administration with a | Incidence rates and 95% confidence intervals (CIs)<br><br>IRR 1 vs 2, 1 vs 3 | Observed incidence:<br>Dose 1: 0.8 (0.2-3.3) cases per 1 million first doses<br>Dose 2: 5.8 (3.4-10) cases per 1 million second doses<br>Unvaccinated: 2.2 (1.7-2.7) cases per million people<br><br>100% cases men<br><br>IRR (95% CI): 1 v 2<br>Dose 1: 0.38 (0.05-1.40)<br>Dose 2: 2.7 (1.4-4.8) |

| Dataset<br>Dates<br>Country | Vaccines Studied | Sample Size; Demographics; Previous Covid-19 diagnoses | Study Group(s) | Outcome(s); Risk Interval; Case Ascertainment | Analysis | Results |
|---|---|---|---|---|---|---|
| | | Black 6.7%; Hispanic 37.8%; Asian 14.3%<br><br>Unvaccinated group: n=1 577 741, median (IQR) age was 39 (28-53) years<br>Age <40y: 53.7%<br><br>Females: 49.1% Black 8.8%; Hispanic 39.2%; Asian 6.6% | | discharge diagnosis of myocarditis<br><br>All cases were independently adjudicated by at least 2 cardiologists.<br><br>Risk interval: 10d | | IRR 1 v 3<br>Dose 1: 1.0 (0.1-13.8)<br>Dose 2: 3.3 (1.0-13.7) |
| Israel MOH*<br>May 31<br><br>Dec 20 2020 to May 31 2021<br><br>Israel<br><br>Mevorach, 2021 | Pfizer-BioNTech<br><br>2 doses to 5.12 million people | Surveillance population 9,289,765 Israeli residents ≥16 y<br><br>29 of 98 cases in unvaccinated population had confirmed Covid-19 (timing NR); NR for vaccinated<br><br>Age, sex, race/ethnicity NR<br><br>None of the post-vaccination cases had concurrent Covid-19 via symptoms or PCR; 35/39 had negative serology tests | 1. Received dose 1 (n=5,442,696)<br>2. Received dose 2 (n=5,125,635)<br>3. Concurrent unvaccinated persons matched by date<br>4. Historical controls from 2017-2019<br><br>Interval between doses 21 d | Myocarditis/myopericarditis<br><br>ICD-9 codes 422.0-9x and 429.0x; cases 12-29 yrs old confirmed by medical record review using Brighton Criteria via consensus<br><br>Assessors not blinded to vaccine status<br><br>Cases of pericarditis without myocarditis were excluded<br><br>Risk interval: 21 d for dose 1 and 30d for dose 2 | Incidence in groups 1 and 2 (cumulative risks)<br>Risk difference (incidence) between 1 vs 2<br>Standardized incidence ratio of observed-to-expected (historical controls)<br>Attributable risk to dose 2<br>IRR group 2 vs 3 (weighted by age and sex)<br><br>Subgroups: sex, 5-yr age categories <30; 10 yr after 30; shorter risk interval 0-7 d | Total events in groups 1 and 2: 142 (136 definitive or probable [117 after dose 2], used for analysis except for standardized IR); group 3: 101<br><br>Incidence: dose 1: 0.35 per 100,000 persons; dose 2: 2.28 per 100,000 persons<br><br>Risk difference dose 2 – dose 1: 1.76 per 100,000 persons (95% CI, 1.33 to 2.19)<br><br>Standardized incidence ratio: group 1 vs 4 1.42 (95% CI, 0.92 to 2.10); group 2 vs 4: 5.34 (95% CI, 4.48 to 6.40)<br><br>aIRR group 2 vs 3: 2.35 (95% CI, 1.10 to 5.02)<br><br><u>Subgroup analyses</u><br>Males vs females:<br>Incidence:<br>dose 1: 0.64 vs 0.07 per 100,000 persons<br>dose 2: 3.83 vs. 0.46 per 100,000 persons<br>Risk difference dose 2 – dose 1: 3.19 (95% CI, 2.37 to 4.02) vs. 0.39 (95% CI, 0.10 to 0.68)<br>IRR: NR<br><br>By age categories:<br>Males<br>Dose 1 incidence per 100,000 persons<br>16-19 y 1.34<br>20-24 y 1.91<br>25-29y 1.22<br>30-39 y 0.41 |

| Dataset<br>Dates<br>Country | Vaccines Studied | Sample Size; Demographics; Previous Covid-19 diagnoses | Study Group(s) | Outcome(s); Risk Interval; Case Ascertainment | Analysis | Results |
|---|---|---|---|---|---|---|
| | | | | | | 40-49y 0.65<br>≥50y 0.10<br>Dose 2 incidence<br>16-19 y 15.07<br>20-24 y 10.86<br>25-29 y 6.99<br>30-39 y 3.69<br>40-49 y 1.15<br>≥50 y 0.21<br>aIRR 16-19 y 8.96 (95% CI, 4.50 to 17.83); 20-24 y 6.13 (95% CI 3.16 to 11.88); 25-29 y 3.58 (95% CI 1.82 to 7.01); ≥30 y 1.00 (95% CI, 0.61 to 1.64)<br>Females<br>Dose 1 incidence<br>16-39 y 0<br>40-49 y 0.21<br>≥50 y 0.09<br>Dose 2 incidence<br>16-19 y 1.00<br>20-24 y 2.16<br>25-29 y 0<br>30-39 y 0.22<br>40-49 y 0.45<br>≥50 y 0.19<br>IRR 20-24 y 7.56 (95 CI, 1.47 to 8.96); others imprecise<br><br>Shorter risk interval (7 d after dose 2):<br>Males 16-19 y:<br>Risk difference: 13.62 per 100,000 persons (95% CI, 8.31 to 19.03)<br>IRR: 31.90 (95% CI, 15.88 to 64.08) |
| Clalit Health**<br>May 24a<br><br>Dec 20 2020 to May 24 2021<br><br>Israel<br><br>Witberg, 2021 | Pfizer-BioNTech<br><br>94% received 2 doses | 2,558,421 ≥16 y receiving at least 1 dose (94% 2 doses)<br><br>2 cases had Covid-19 (125 and 186 d) prior to vaccine but NR if any others and NR if active testing of cases | 1. Received 1 dose (n=2,558,421)<br><br>Interval between doses 21 d | Myocarditis<br><br>Risk interval: 42 d after dose 1<br><br>ICD-9 (codes 422, 429.0, 398.0, and 391.2 and their respective subcodes), with adjudication by cardiologists using medical and CDC case definition, with consensus | Incidence after dose 1 or 2 using Kaplan–Meier analysis<br><br>Subgroups: Sex; Ages 16-29 y; ≥30 y | Events: 54 (of 159 potential cases; 37 after dose 2; with 17/37 >7 days post dose 2)<br><br>Incidence: 2.13 cases per 100,000 persons (95% CI: 1.56 to 2.70)<br><br><u>Subgroup analysis</u><br>Males vs females:<br>Incidence: 4.12 (95% CI: 2.99 to 5.26) vs. 0.23 (95% CI: 0 to 0.49)<br><br><u>By age categories</u>:<br>Overall 16-29 y: 5.49 per 100,000 persons (95% CI: 3.59 to 7.39)<br>Overall ≥30 y: 1.13 per 100,000 persons (95% CI: 0.66 to 1.60)<br>Males 16-29 y: 10.69 per 100,000 persons (95% CI: 6.93 to 14.46) |

| Dataset<br>Dates<br>Country | Vaccines Studied | Sample Size; Demographics; Previous Covid-19 diagnoses | Study Group(s) | Outcome(s); Risk Interval; Case Ascertainment | Analysis | Results |
|---|---|---|---|---|---|---|
| | | | | Unblinded to vaccine status (all vaccinated and author adjudicating) | | Males ≥30 y: 2.11 per 100,000 persons (95% CI, 1.19 to 3.04)<br>Females 16-29 y: 0.34 per 100,000 persons (95% CI, 0 to 1)<br>Females ≥30 y: 0.20 per 100,000 persons (95% CI, 0 to 0.48) |
| Clalit Health** May 24b<br><br>Dec 20 2020 to May 24 2021<br><br>Israel<br><br>Barda, 2021 | Pfizer-BioNTech<br><br>NR how may had 1 vs 2 doses | Surveillance population 1,877,624 ≥16 y<br><br>Excluded people with previous events, residing in nursing homes, healthcare workers, no BMI or residential area data<br><br>Mean age 38 y<br>Females 48%<br><br>Previous Covid-19 cases excluded | 1. Received 1 dose (n=884,828)<br>2. Concurrent unvaccinated, matched 1:1 by age, sex, place of residence, socioeconomic status, population sector (general Jewish, Arab, or ultra-Orthodox Jewish), number of preexisting conditions at risk for severe Covid-19. (n=884,828)<br><br>Interval between doses 21 d | Myocarditis & pericarditis after at least 1 dose<br><br>Risk interval: 42 d after dose 1<br><br>ICD-9 codes (422, 429.0, 398.0, 391.2, 420) after the index date<br><br>Blinding NR | Incidence using Kaplan–Meier estimator<br><br>Risk difference<br>Risk Ratio | Events in group 1 vs 2: myocarditis 21 (91% male; median age 25y [IQR 20-34]) vs 6; pericarditis: 27 vs 18<br><br>Risk difference (all ages): myocarditis: 2.7 events per 100,000 persons (95% CI, 1.0 to 4.6); pericarditis: 1.0 events per 100,000 persons (95% CI, −1.6 to 3.4)<br><br>RR: myocarditis: 3.24 (95% CI, 1.55 to 12.44); pericarditis: 1.27 (95% CI, 0.68 to 2.31) |
| Israeli Defense Forces May 7<br><br>Dec 28 2020 to Mar 7 2021<br><br>Israel<br><br>Levin, 2021 | Pfizer-BioNTech<br><br>138,000 military personnel receiving 2 doses | 138,000<br><br>NR<br><br>NR | 1. Vaccinated with 2 doses (n=138,000)<br><br>Interval between doses NR | Myocarditis<br><br>Medical record review, requiring ECG, echocardiography, or MRI findings<br><br>Risk interval: 7 d after dose 2<br><br>Not blinded | Crude cumulative incidence | Events: 7 confirmed in risk interval (100% male; Age 18-24)<br><br>Incidence: 5.07 per 100,000 people |
| Genesis Healthcare*** Feb 14<br><br>Dec 18 2020 to Feb 14 2021<br><br>USA<br><br>Bardenheier, 2021a | Moderna or Pfizer-BioNTech<br><br>All had 1 dose | 20,918 nursing home residents<br><br>Females 61.9%<br><65 y 18.5%<br>African American or Latinx 16.9%<br><br>Excluded if Covid-19 +ve within 20 d of vaccination, or treated | 1. No prior infection at vaccination (n=13.163)<br>2. Symptomatic infection >20 days before vaccination (n=5,617)<br>3. Asymptomatic infection >20 days before vaccination (n=2,138)<br>4. Unvaccinated (from Bardenheier, 2021b)<br><br>Interval between doses NA | Myocarditis/pericarditis<br><br>Risk interval: 15 days after dose 1<br><br>Active, prospective surveillance; medical record review for ICD-10-CM codes, confirmed by physician chart review, using Brighton Collaboration Criteria | Incidence (if events)<br><br>Comparisons between groups by multilevel logistic regression that adjusted for clustering and applied propensity scoring for age, sex, pre-existing conditions | Events: group 1 vs 2 vs 3: 0 vs. 0 vs. 0 |

| Dataset<br>Dates<br>Country | Vaccines Studied | Sample Size; Demographics; Previous Covid-19 diagnoses | Study Group(s) | Outcome(s); Risk Interval; Case Ascertainment | Analysis | Results |
|---|---|---|---|---|---|---|
| | | with SARS-CoV-2 monoclonal antibodies within 90 days before vaccination | | Blinding NR | | |
| Genesis Healthcare***<br>Jan 3<br><br>Dec 18 2020 to Jan 3 2021<br><br>USA<br><br>Bardenheier, 2021b | Moderna or Pfizer-BioNTech (70.6%)<br><br>1st dose or 2nd dose | 21,222 nursing home residents<br><br>Females 61.9%<br><65 y 18.8<br>African American or Latinx 18%<br><br>Excluded if Covid-19 +ve within 20 d of vaccination, or treated with SARS-CoV-2 monoclonal antibodies within 90 days before vaccination | 1. After dose 1 (n=8553)<br>2. After dose 2 (n=8371)<br>3. Unvaccinated (n=11072)<br><br>Interval between doses 3 to 6 weeks | Myocarditis<br><br>Active, prospective surveillance; medical record review for ICD-10 codes, confirmed by physician chart review, using Brighton Collaboration Criteria<br><br>Blinding NR | Incidence<br><br>IRR, adjusted for age, sex, pre-existing conditions | Events: 0 vs. 0 vs. 0 |

**Table A2. Study characteristics of observational studies using data from passive surveillance systems (question 1)**

| Dataset<br>Dates of data<br>Country of Data | Vaccines Studied | Outcome(s); Case Ascertainment & Risk Interval | Analysis | Results |
|---|---|---|---|---|
| VAERS* Oct 6<br><br>Up to Oct 6, 2021<br><br>USA<br><br>Su, 2021 | Pfizer-BioNTech, Moderna and Janssen | Myopericarditis (myocarditis +/- pericardtis); pericarditis<br><br>Screening via 30 MedDRA terms and ICD-10; all analyzed reports verified to meet CDC case definition by provider interview or medical record review<br><br>Risk interval: 7 d | Crude reporting rates of confirmed cases of myopericarditis after each dose | Total events: 2,459 myopericarditis (1,181 have been verified and 935 occurred in 0-6 d after vaccination; 877 pericarditis (366,062,239 doses of mRNA vaccines)<br><br>For myopericarditis: 67% Pfizer; 29% Moderna; 76% after dose 2 (50 preliminary reports after Janssen not in analysis)<br><br><u>Reporting rates of myopericarditis per 1 million doses administered (N=935 verified cases):</u><br>**Pfizer** **Moderna**<br>Ages Dose 1 Dose 2 Dose 1 Dose 2<br>12-15 **2.3*** **21.5** 0.0 not calculated<br>16-17 **2.8** **37.4** 0.0 not calculated<br>18-24 1.2 **18.1** **3.1** **20.7**<br>25-29 0.7 **5.7** 1.8 **11.2**<br>30-39 0.6 **2.8** 1.4 **3.6**<br>40-49 0.2 1.5 0.2 **2.1**<br>50-64 0.3 0.4 0.5 0.5<br>65+ 0.1 0.2 0.0 0.3<br><br><u>Males (N=797):</u><br>Ages Dose 1 Dose 2 Dose 1 Dose 2<br>12-15 **4.2** **39.9** 0.0 not calculated<br>16-17 **5.7** **69.1** 0.0 not calculated<br>18-24 **2.3** **36.8** **6.1** **38.5**<br>25-29 1.3 **10.8** **3.4** **17.2**<br>30-39 0.5 **5.2** **2.3** **6.7**<br>40-49 0.3 2.0 0.2 **2.9**<br>50-64 0.2 0.3 0.5 0.6<br>65+ 0.2 0.1 0.1 0.3<br><br><u>Females (N=138):</u><br>Ages Dose 1 Dose 2 Dose 1 Dose 2<br>12-15 0.4 **3.9** 0.0 0.0<br>16-17 0.0 **7.9** 0.0 0.0<br>18-24 0.2 **2.5** 0.6 **5.3**<br>25-29 0.2 1.2 0.4 **5.7**<br>30-39 0.6 0.7 0.5 0.4<br>40-49 0.1 1.1 0.2 1.4<br>50-64 0.3 0.5 0.5 0.4<br>65+ 0.1 0.3 0.0 0.3<br><br>*Reporting rates exceed background incidence (bolded data above))<br>An estimated 1–10 cases of myocarditis per 100,000 person years occurs among people in the United States, regardless of vaccination status; adjusted for the 7-day risk period, this estimated background is 0.2 to 1.9 per 1 million person 7-day risk period |
| VAERS* Jun 30<br><br>Up to Jun 30 2021 | Pfizer-BioNTech or Moderna | Myocarditis | Crude reporting rates after dose 2<br><br>Subgroups by age and sex | Overall ≥18 yrs: 3.5 cases per million second doses<br><br><u>Subgroups (all after dose 2):</u> |

| Dataset<br>Dates of data<br>Country of Data | Vaccines Studied | Outcome(s); Case Ascertainment & Risk Interval | Analysis | Results |
|---|---|---|---|---|
| USA<br><br>Rosenblum, 2021 | After dose 2, timing NR | Cases in persons aged 18–29 yrs were individually reviewed and confirmed to meet CDC case definitions<br><br>Risk interval: 42 d | | Females<br>18-29 yrs: 3-4 cases per million second doses<br>30-49 yrs:1-2 cases per million second doses<br>50-64 yrs: 1 case per million second doses<br>≥65 yrs: <1 case per million second doses<br><br>Males<br>18-29 yrs: 22-27 cases per million second doses<br>30-49 yrs: 5-6 cases per million second doses<br>50-64 yrs: 1 case per million second doses<br>≥65 yrs: 1 case per million second doses |
| VAERS* Jun 11<br><br>Up to Jun 11 2021<br><br>USA<br><br>Gargano 2021 | Pfizer-BioNTech or Moderna<br><br>After dose 2, timing NR | Myocarditis<br><br>Note: Subset of confirmed cases described and included in KQ2<br><br>Risk interval: 7 d | Crude reporting rates (i.e., using confirmed and unconfirmed cases), after dose 2 | Males 12-29 yrs: 40.6 cases per million second doses<br>Males 12-17 yrs: 62.8 cases per million second doses<br>Males 18-24 yrs: 50.5 cases per million second doses<br>Males ≥30 yrs: 2.4 cases per million second doses<br>Females 12-29 yrs: 4.2 cases per million second doses<br>Females ≥30 yrs: 1.0 cases per million second doses |
| VAERS* Aug 6<br><br>Up to Aug 6 2021<br><br>US<br><br>Lane 2021 | Pfizer-BioNTech, Moderna<br><br>At least one dose (UK, EEA) or 2 doses (US); schedule NR. | Myocarditis and pericarditis<br><br>Events labelled “Myocarditis” or “Pericarditis”<br><br>Risk interval: None applied (date of vaccine launch to datalock point) | Crude reporting rates per million people receiving at least one dose (UK, EEA) or receiving both doses (US) | UK<br>Pfizer/BioNTech: 7.93 cases of myocarditis and 6.73 cases pericarditis per million vaccinees<br>Moderna: 2.07 cases of myocarditis and 1.79 cases of pericarditis per million vaccinees<br><br>EEA<br>Pfizer/BioNTech: 4.23 cases of myocarditis and 2.87 cases of pericarditis per million vaccinees<br>Moderna: 6.15 cases of myocarditis and 3.84 cases of pericarditis per million vaccinees<br><br>US (after dose 2)<br>Pfizer/BioNTech: 6.47 cases of myocarditis and 3.53 cases of pericarditis per million vaccinees<br>Moderna: 3.65 cases of myocarditis and 2.69 cases of pericarditis per million vaccinees |
| VAERS* Jun 18a<br><br>Jan 1 to Jun 18 2021<br><br>USA<br><br>Høeg 2021 | Pfizer-BioNTech or Moderna (but only 1 of 257 cases since not approved for <18y at that time)<br><br>Schedule NR | Myocarditis<br><br>“Myocarditis,” “pericarditis,” “myopericarditis” or “chest pain” in the symptom notes; “troponin” required element in the laboratory data.<br><br>CDC working case definition of probable myocarditis<br><br>Risk interval: Any timing | Crude rates per million vaccinees<br><br>Cases with an unknown dose number were assigned to dose 1 or dose 2 in the same proportion as the known doses: 15% occurred following dose 1 and 85% occurred following dose 2 | Events: 257 (92% within 5 d; 90% males)<br><br>At least Dose 1<br>Males 12-15 yrs: 12.0 per million vaccinees<br>Males 16-17 yrs: 8.2 per million vaccinees<br>Females 12-15 yrs: 0 per million vaccinees<br>Females 16-17 yrs: 2.0 per million vaccinees<br>Dose 2<br>Males 12-15 yrs: 162.2 per million vaccinees<br>Males 16-17 yrs: 94.0 per million vaccinees<br>Females 12-15 yrs: 13.0 per million vaccinees<br>Females 16-17 yrs: 13.4 per million vaccinees |
| VAERS* Jun 18b<br><br>USA<br><br>Lazaros 2021 | AstraZeneca<br>Janssen<br>Moderna<br>Pfizer-BioNTech | Pericarditis<br><br>Ascertainment NR | Crude rate per million vaccine doses<br><br>Raw data available to calculate crude age and sex subgroup rates if needed. | EEA<br>AstraZeneca: 2.604 per million doses<br>Janssen: 2.530 per million doses<br>Total vectored: 2.596 per million doses<br>Moderna: 4.023 per million doses<br>Pfizer-BioNTech: 1.613 per million doses |

| Dataset<br>Dates of data<br>Country of Data | Vaccines Studied | Outcome(s); Case Ascertainment & Risk Interval | Analysis | Results |
|---|---|---|---|---|
| | Dose and schedule not specified. | | | Total mRNA: 1.882 per million doses |
| CAEFISS & CVP<br><br>Up to Oct 8 2021<br><br>Canada<br><br>PHAC 2021 | AstraZeneca, Moderna (≥12 y), Pfizer-BioNTech (≥12 y)<br><br>Dose and schedule not specified. | Myocarditis or pericarditis<br><br>Cases meeting Level 1-4 to Brighton Criteria of Diagnostic Certainty (not all probable or definitive)<br><br>Risk interval: Any timing | Reported rates per 100,000 doses administered | Events: 913<br><br>Moderna: 2.51 cases per 100,000 doses<br>Pfizer-BioNTech: 1.37 cases per 100,000 doses<br>AstraZeneca: 0.72 cases per 100,000 doses |
| Yellow Card Oct 13<br><br>Dec 9 2020 to Oct 13 2021<br><br>UK<br><br>Medicines & Healthcare Products Regulatory Agency 2021 | AstraZeneca, Moderna, Pfizer-BioNTech<br><br>After dose 1 or 2; schedule NR | Myocarditis and pericarditis (including those from viral/other infective causes)<br><br>Ascertainment NR<br><br>Risk interval: NR | Reporting rates per million doses | <u>Overall</u><br>Pfizer-BioNTech: myocarditis 8 cases per million doses; pericarditis 6 cases per million doses<br>Moderna: myocarditis 32 case per million doses; pericarditis 19 cases per million doses<br>AstraZeneca: myocarditis 3 case per million doses; pericarditis 4 cases per million doses<br><br><u>Subgroups:</u><br>Myocarditis/pericarditis<br><18 yrs<br>Pfizer-BioNTech: 10 cases per million doses<br>Moderna: 0 cases<br>AstraZeneca: 0 cases<br><br>18-49 yrs<br>Pfizer-BioNTech: 18 cases per million doses<br>Moderna: 44 cases per million doses<br>AstraZeneca: 8 cases per million doses<br><br>≥50 yrs<br>Pfizer-BioNTech: 5 cases per million doses<br>Moderna: 29 cases per million doses<br>AstraZeneca: 4 cases per million doses |
| Israel MOH* May 30<br><br>Dec 2020 to May 30 2021<br><br>Israel | Pfizer-BioNTech (≥16 y)<br><br>Dose 1 and 2; schedule NR | Myocarditis<br><br>Ascertainment NR<br><br>Risk interval: 30 d after dose 2 | Crude number of cases and doses | Dose 1:<br>27 cases of 5,401,150 vaccinated individuals (5 per million) (11 had pre-existing conditions)<br><br>Within 30 d after dose 2:<br>121 cases of 5,049,424 vaccinated individuals (24 per million) (60 had pre-existing conditions) |
| EudraVigilance Oct 19**<br><br>Up to Oct 19 2021<br><br>EEA<br><br>European Medicines Agency 2021 | AstraZeneca<br>Janssen<br>Moderna<br>Pfizer-BioNTech<br><br>Dose and schedule NR | Myocarditis, pericarditis<br><br>Ascertainment NR<br><br>Risk interval: NR | Crude number of case and doses | <u>Overall</u>:<br>Moderna: 1578 cases of myocarditis and 850 cases of pericarditis across 61,172,753 doses administered† (25.8 per million & 13.9 per million)<br>Pfizer: 3610 cases of myocarditis and 2883 cases of pericarditis across 424,162,518 doses administered† (8.5 per million & 6.8 per million)<br>AstraZeneca: 320 cases of myocarditis and 416 cases of pericarditis across 68,799,957 doses administered† (4.7 per million & 6.0 per million)<br>Janssen: 103 cases of myocarditis and 101 cases of pericarditis across 15,782,596 doses administered† (6.5 per million & 6.4 per million)<br><br><u>Age<18y</u> |

| Dataset<br>Dates of data<br>Country of Data | Vaccines Studied | Outcome(s); Case Ascertainment & Risk Interv al | Analysis | Results |
|---|---|---|---|---|
| | | | | Moderna: 32 reports of myocarditis across 531,050 people receiving at least one dose† (60.3 events per million)<br>Pfizer: 369 reports of myocarditis across 8,874,141 people receiving at least one dose† (41.6 events per million) |

*Indicates passive surveillance system with mandatory/legal reporting requirements for healthcare providers of adverse events after COVID-19 vaccines.

†Number of administered vaccine doses from European Center for Disease Control (EDCD), up to end of Week 41 2021 (Oct 16 2021). Period of vaccine doses is shorter than event reporting to account for time period between receiving vaccine and experiencing the event of interest (i.e., individuals vaccinated on October 19 are unlikely to be reporting myocarditis as an AE on that same day)

### Table A3. Summary of risk of bias assessments for observational studies/surveillance data (question 1)

| Dataset | Were the two groups similar and recruited from the same population? | Was vaccination status measured in a reliable or valid way? | Were confounding factors identified and appropriately addressed in design or analysis? | Were the outcomes measured in a valid and reliable way? | Was the follow up time long enough for outcomes to occur? | Were the large majority of cases likely to have been identified? | Overall assessment of risk of bias |
|---|---|---|---|---|---|---|---|
| **Active Surveillance Studies** | | | | | | | |
| VSD Jun 26 Klein 2021 Ref ID 22 | Y | Y | U | Y | Y | Y | Some concerns |
| Israel MOH May 31 Mevorach 2021 Ref ID 3440 | Y | U | N | Y | Y | Y | Some concerns |
| Clalit Health May 24a Witberg Ref ID 3441 | NA | Y | Y | Y | Y | Y | Low |
| Clalit Health May 24b Barda 2021 Ref ID 1282 | Y | Y | Y | Y | Y | Y | Low |
| Israeli Defense Forces May 7 Levin 2021 Ref ID 512 | NA | Y | N | Y | N | Y | Some concerns |
| Genesis Healthcare Feb 14 Bardenheier 2021a Ref ID 144 | Y | Y | Y | Y | U | Y | Some concerns |
| Genesis Healthcare Jan 3 Bardenheier, 2021b ref ID 1972 | Y | Y | Y | Y | U | Y | Some concerns |
| KPSC Jul 2 Simone 2021 Ref ID 22 | Y | Y | U | Y | U | Y | Some concerns |
| Providence Health May 25 Diaz 2021 Ref ID 525 | Y | Y | N | U | N | Y | High |
| VSD Oct 9 (Klein 2021 Oct ACIP) | Y | Y | U | Y | Y | Y | Some concerns |
| VSD Aug 21 | Y | Y | U | Y | Y | Y | Some concerns |

| | | | | | | | |
|---|---|---|---|---|---|---|---|
| Klein 2021 Aug ACIP) | | | | | | | |
| **Passive Surveillance Studies** | | | | | | | |
| VAERS Oct 6 Su, 2021 (ACIP Oct 20-21 2021 meeting) | NA | N | N | Y | Y | N | High |
| VAERS Jun 30 Rosenblum, 2021 | NA | N | N | U | U | N | High |
| VAERS Jun 11 Gargano 2021 | NA | N | N | U | Y | N | High |
| VAERS Aug 6 Lane 2021 | NA | N | N | N | U | N | High |
| VAERS Jun 18a Høeg 2021 | NA | N | N | U | U | N | High |
| VAERS Jun 18b Lazaros 2021 | NA | U | N | U | U | N | High |
| CAEFISS & CVP PHAC 2021 | NA | U | N | N | N | U | High |
| Yellow Card Oct 13 Medicines & Healthcare Products Regulatory Agency 2021 | NA | U | N | U | U | N | High |
| Israel MOH May 30 Israel Ministry of Health 2021 | NA | U | N | U | Y | U | High |
| EudraVigilance Oct 19 European Medicines Agency 2021 | NA | Y | N | U | U | N | High |

### Table A4. Study characteristics of randomized controlled trials (question 1)

| Trial name; Author (year); Country (number of sites); Dates of enrolment Funding | Eligibility, Randomization & Blinding | Study Arms | | Baseline Demographics | Outcome(s); Safety Assessment | Analysis | Results |
|---|---|---|---|---|---|---|---|
| | | Vaccine arm | Placebo arm | | | | |
| **Teen COVE trial, Phase 2/3,** | Included adolescents 12-17 y | mRNA-1273 (Moderna), 2 doses 28 d apart | Saline placebo 2 doses 28 d apart | Total randomized: 3,732 | Myocarditis & pericarditis | Safety analysis included all | **Myocarditis:** 0<br>**Pericarditis:** 0 |

| Trial name;<br>Author (year);<br>Country (number of sites);<br>Dates of enrolment<br>Funding | Eligibility,<br>Randomization &<br>Blinding | Study Arms | | Baseline Demographics | Outcome(s);<br>Safety Assessment | Analysis | Results |
|---|---|---|---|---|---|---|---|
| | | Vaccine arm | Placebo arm | | | | |
| Ali (2021)<br><br>USA (26)<br><br>Dec 9, 2020 –<br>Feb 28, 2021<br><br>USA (26)<br><br>Industry-funded | Exclusions: travel outside of USA 28 d prior to screening; acute illness or fever 24 h prior to or at screening; previous administration of an investigational SARS-CoV-2 vaccine; pregnant or breastfeeding.<br><br>Randomized 2:1; centralized interactive response technology system<br><br>Study personnel and participants were blinded; pharmacists and vaccine administrators unblinded. | 100 mcg per 0.5 ml dose<br><br>Randomized: 2,489<br>Dose 1: 2,486<br>Dose 2: 2,480<br><br>Safety population: 2,486<br>Withdrawals:<br>After dose 1: 18<br>After dose 2: 188 | 0.5 ml per dose<br><br>Randomized: 1,243<br>Dose 1: 1,240<br>Dose 2: 1,222<br><br>Safety population: 1,240<br><br>Withdrawals:<br>After dose 1: 6<br>After dose 2: 57 | Total safety population: 3,726<br><br>Male 51%<br>Mean age 14.3 y<br>12-15 y: 74%<br>16-17 y: 26%<br>White 84%<br><br>Baseline SARS-CoV-2 status:<br>Positive: 216 (6%), similar between arms;<br>Missing: 268 (7%), similar between arms | Solicited local and systemic adverse reactions during 7 d after each injection;<br>**Unsolicited adverse events (AEs)**, defined as any event not present before exposure to study vaccination or any event already present that worsens in intensity or frequency after exposure, observed or reported during 28 d after each injection;<br>AEs leading to discontinuation from a dose and/or study withdrawal, medically attended (MAAEs) and serious AE (SAEs), throughout study period<br><br>Unblinded safety data reviewed by independent data and safety monitoring board | participants who received at least one injection, according to treatment group | |
| **COVE trial, Phase 3**<br><br>Baden (2021; primary publication)<br>& El-Sahly (2021)<br><br>Jul 27, 2020 – Oct 23, 2020<br><br>USA (99)<br><br>Industry-funded | Included adults ≥18 y at appreciable risk of acquiring infection or high risk of severe COVID-19, or both;<br><br>Exclusions: immunosuppression or received systemic immunosuppressant/ immune-modifying drugs for >14 d total in prior 6 mos; known history of SARS-CoV-2 infection; pregnant or breastfeeding, | mRNA-1273 (Moderna)<br>2 doses 28 d apart<br>100 mcg per 0.5 ml dose<br><br>Randomized: 15,210<br>Dose 1: 15,181<br>Dose 2: 14,711<br><br>Safety population (unsolicited AEs): 15,185<br><br>Withdrawals: 233 (45 discontinued study due to SARS-CoV-2 diagnosis) | Saline placebo<br>2 doses 28 d apart<br>0.5 ml dose<br><br>Randomized: 15,210<br>Dose 1: 15,170<br>Dose 2: 14,617<br><br>Safety population (unsolicited AEs): 15,166<br><br>Withdrawals: 290 (69 discontinued study due to SARS-CoV-2 diagnosis) | Total randomized: 30,420<br>Total safety population: 30,351<br><br>Male 52.6%<br>Mean age 51.4 y<br>White 79.2%<br>18 to <65 y at risk: 16.7%<br>18 to <65 y not at risk: 58.6%<br>≥65 y: 24.8%<br>Significant cardiac disease 4.9%<br>HIV infection 0.6%<br><br>Baseline SARS-CoV-2 status: | Solicited local and systemic adverse reactions during 7 d following each injection;<br>**Unsolicited adverse events (AEs)** for 28 d following each injection, including treatment-emergent AE (any event during study not present before exposure to injection or any event already present that worsens after exposure to study vaccine; | Safety analysis included all participants who received at least one injection, according to product received. | From associated publication:<br><br>**Myocarditis:** 0<br>**Pericarditis:** 2 (68 d & 73 d after dose 2) vaccine vs. 2 placebo |

| Trial name;<br>Author (year);<br>Country (number of sites);<br>Dates of enrolment<br>Funding | Eligibility,<br>Randomization &<br>Blinding | Study Arms | | Baseline Demographics | Outcome(s);<br>Safety Assessment | Analysis | Results |
|---|---|---|---|---|---|---|---|
| | | Vaccine arm | Placebo arm | | | | |
| | Randomized 1:1, stratified by age (≥18 to <65 y vs. ≥65 y) and being at risk for severe Covid-19 based on pre-existing risk factors as per CDC; centralized interactive response technology system<br><br>Study personnel and participants were blinded, but pharmacists and vaccine administrators were unblinded; participants informed of group assignment at end of blinded phase and offered vaccine. | | | Positive: 680 (2.2%), similar between arms;<br>Missing: 523 (1.7%), similar between arms | AEs leading to discontinuation from a dose and/or study withdrawal, MAAEs and SAEs, throughout study period (1-759 d)<br><br>MedDRA classification for all solicited adverse reactions and unsolicited AEs<br><br>Unblinded safety data reviewed by independent data and safety monitoring board | | |
| **C4591001 Trial,<br>Phase 3 - adolescent cohort**<br><br>Frenck (2021)<br><br>Oct 15, 2020 – Jan 12, 2021<br><br>Argentina (NR), Brazil (NR), Germany (NR), South Africa (NR), Turkey (NR), USA (29)<br><br>Industry-funded | Included adolescents 12-15 y and adults 16-25 y who were healthy or with stable pre-existing conditions<br><br>Exclusions: medical history of SARS-CoV-2 infection or Covid-19 diagnosis; treatment with immunosuppressive therapy; diagnosed with immunocompromising condition<br><br>Randomized 1:1; web-based system<br><br>Study personnel evaluating safety | BNT162b2 (Pfizer-BioNTech)<br>2 doses 21 d apart<br>30 mcg per 0.3 ml dose<br><br>Randomized:<br><br>12-15 y: 1,134<br>Dose 1: 1,131<br>Dose 2: 1,124<br><br>16-25 y: 1,875<br>Dose 1: 1,869<br>Dose 2: 1,826<br><br>Withdrawals:<br><br>12-15 y<br>After dose 1: 7<br>After dose 2: 0<br><br>16-25 y<br>After dose 1: 43<br>After dose 2: 20 | Saline placebo<br>2 doses 21 d apart<br>0.3 ml per dose<br><br>Randomized:<br><br>12-15 y: 1,130<br>Dose 1: 1,129<br>Dose 2:1,117<br><br>16-25 y: 1,913<br>Dose 1: 1,906<br>Dose 2: 1,836<br><br>Withdrawals:<br><br>12-15 y<br>After dose 1: 12<br>After dose 2: 1<br><br>16-25 y | Total randomized:<br>12-15 y: 2,264<br>16-25 y: 3,788<br><br>Total safety population:<br>12-15 y: 2,260<br>16-25 y: 1,098 (subset with e-diary data)<br><br>Male 51%<br>White 86%<br><br>Baseline SARS-CoV-2: Positive:<br>12-15 y: 93 (4.1%), similar between arms<br>16-25 y: 64 (5.8%), similar between arms | Solicited local and systemic AEs during 7 d following each injection;<br>**Unsolicited AEs** for 1 mo after dose 1 and SAEs for 6 mos after dose 2.<br><br>MedDRA v23.1 classification.<br><br>Unblinded safety data reviewed by independent data and safety monitoring board; ongoing for 2 y after dose 2. | 12-15 y: Safety analysis included all participants who received at least one injection.<br><br>16-25 y: subset of participants who received an e-diary to record reactogenicity events | Myocarditis/pericarditis: 0 |

| Trial name;<br>Author (year);<br>Country (number of sites);<br>Dates of enrolment<br>Funding | Eligibility,<br>Randomization &<br>Blinding | Study Arms<br>Vaccine arm | <br>Placebo arm | Baseline Demographics | Outcome(s);<br>Safety Assessment | Analysis | Results |
|---|---|---|---|---|---|---|---|
| | were blinded to group assignments | | After dose 1: 70<br>After dose 2: 22 | | | | |
| C4591001 Trial, Phase 2/3<br><br>Polack (2021, primary publication) & Thomas (2021)<br><br>Jul 27, 2020 – October 29, 2020<br><br>Argentina (1), Brazil (2), Germany (6), South Africa (4), Turkey (9), USA (130)<br><br>Industry-funded | Included participants ≥16 y, healthy or with stable pre-existing conditions<br><br>Exclusions: medical history of SARS-CoV-2 infection or Covid-19 diagnosis; treatment with immunosuppressive therapy; diagnosed with immunocompromising condition<br><br>Randomized 1:1; web-based system<br><br>Study personnel evaluating safety were blinded to group assignments; unblinded follow-up started Dec 2020 | BNT162b2 (Pfizer-BioNTech)<br>2 doses 21 d apart<br>30 mcg per 0.3 ml dose<br><br>Randomized: 22,085<br>Dose 1: 22,030<br>Dose 2: 21,759<br><br>Safety population: 21,926<br><br>Withdrawals:<br>After dose 1: 271<br>After dose 2: 167 | Saline placebo<br>2 doses 21 d apart<br>0.3 ml dose<br><br>Randomized: 22,080<br>Dose 1: 22,030<br>Dose 2: 21,650<br><br>Safety population: 21,921<br><br>Withdrawals:<br>After dose 1: 380<br>After dose 2: 273 | Total randomized: 44,165<br><br>Total safety population: 43,847<br><br>Male 50.9%<br>Median age 51.0 y<br>16-55 y 59.4%<br>>55 y 40.6%<br>White 82.0%<br>Congestive heart failure 0.5%<br>Myocardial infarction 1.0%<br>Any malignancy 3.6%<br>Rheumatic disease 0.3%<br>AIDS/HIV 0.5%<br><br>Baseline SARS-CoV-2 status:<br>Positive: 1,405 (3.2%), similar between arms<br>Missing: 277 (0.6%), similar between arms | Solicited local and systemic AEs during 7 d after each injection; **Unsolicited AEs** and SAEs for 1 mo and 6 mos after dose 2, respectively.<br><br>MedDRA v23.1 classification;<br><br>Unblinded safety data reviewed by independent data and safety monitoring board; ongoing for 2 y after second dose. | Safety analysis included all participants who received at least one injection irrespective of follow-up time. | **Myocarditis:** 0 |
| **ENSEMBLE Trial, Phase 3**<br><br>Sadoff (2021)<br><br>Sep 21, 2020 – Jan 22, 2021<br><br>Argentina, Brazil, Chile, Colombia, Mexico, Peru, South Africa, USA<br><br>Industry-funded | Adults ≥18 y, healthy or with stable pre-existing conditions; excluded those who received chronic/recurrent systemic corticosteroids, or antineoplastic and immunomodulating agents or radiotherapy in prior 6 mo before study vaccine and during study, or are pregnant<br><br>Randomized 1:1, stratified by site, age, and being at risk for severe Covid-19 | Ad26.COV2.S (Janssen)<br>1 dose<br>$5 \times 10^{10}$ vp per 0.5 ml dose<br><br>Randomized: 21,895<br><br>Safety population: 21,895<br><br>Withdrawals: 49 | Saline placebo<br>1 dose x 0.5ml<br><br>Randomized: 21,888<br><br>Safety population: 21,888<br><br>Withdrawals: 96 | Total randomized: 44,325<br><br>Total safety population: 43,783<br><br>Male 54.9%<br>Median age 52 y<br>White 58.7%<br>Serious heart condition 2.3%<br>Immunocompromised from blood transplant, immune deficiencies, use of corticosteroids or other immunosuppressing medicines 0.2%<br>Immunocompromised from organ transplant <0.1%<br><br>Baseline SARS-CoV-2 status: | Solicited local and systemic AEs 7 d after injection (for subset of 6,000 participants); **Unsolicited SAEs** up to 28 d after injection, MAAEs until 6 mos after vaccination, and MAAEs leading to study withdrawal<br><br>Independent data and safety monitoring board reviewed unblinded safety data. | Safety analysis included all participants who received injection | **Pericarditis:** 1 (<0.1%) vaccine vs. 0 placebo |

| Trial name;<br>Author (year);<br>Country (number of sites);<br>Dates of enrolment<br>Funding | Eligibility,<br>Randomization &<br>Blinding | Study Arms<br>Vaccine arm | <br>Placebo arm | Baseline Demographics | Outcome(s);<br>Safety Assessment | Analysis | Results |
|---|---|---|---|---|---|---|---|
| | based on pre-existing risk factors; interactive web response system<br><br>Participants and study personnel (including HCW administering vaccine) blinded until either discontinuation or an event | | | Positive: 4,217 (9.6%), similar between arms<br>Missing: 1,271 (2.9%), similar between arms | | | |
| **AZD1222 Trial, Phase 3**<br><br>Falsey (2021)<br><br>Aug 28, 2020 – Jan 15, 2021<br><br>Chile, Peru, USA (88 total)<br><br>Industry-funded | Included adults ≥18 y, with stable pre-existing conditions, at high risk for exposure to SARS-CoV-2 and at increased risk for severe Covid-19<br><br>Exclusions: history of SARS-CoV-2 infection; confirmed or suspected immunodeficiency, or with significant disease; pregnant or breastfeeding<br><br>Randomized 2:1, stratified by age; method NR<br><br>Group assignment unblinded for 7,635 (35.3%) AstraZeneca and 4,157 (38.4%) placebo participants, after the second dose. | AZD1222 (AstraZeneca)<br>2 doses 4 wks apart<br>5 x $10^{10}$ vp per dose<br><br>Randomized: 21,635<br>Dose 1: 21,583<br>Dose 2: 20,769<br><br>Safety population: 21,587<br><br>Withdrawals:<br>After dose 1: 283<br>After dose 2: 228 | Saline placebo<br>2 doses 4 wks apart<br><br><br>Randomized: 10,816<br>Dose 1: 10,769<br>Dose 2: 9951<br><br>Safety population: 10,792<br><br>Withdrawals:<br>After dose 1: 240<br>After dose 2: 258 | Total randomized: 32,451<br><br>Total safety population: 32,379<br><br>Male 55.6%<br>Mean age: 50.2 y<br>≥18-64 y 77.6%<br>≥65 y 22.4%<br>White 79.0%<br>Serious heart condition 3.3%<br>Cancer 6.5%<br>Immunocompromised due to solid organ transplantation <0.1%<br>HIV infection 1.6%<br><br>Baseline SARS-CoV-2 status:<br>Positive: 915 (2.8%), similar between arms<br>Missing/not performed: 575 (1.8%), similar between arms | Solicited local and systemic AEs 7 d after each injection;<br>**Unsolicited AEs** up to 28 d after injection, SAEs, MAAEs, and MEs of special interest up to 730 d after informed consent | Safety analysis included all participants who received at least one injection. | Myocarditis/pericarditis: 0 |
| **AZD1222**<br>**Phase 1/2/3, pooled analysis**<br><br>Voysey (2021a; primary publication) & Voysey (2021b)<br><br><br>Apr 23, 2020 – Dec 6, 2020 | Adults ≥18 y, healthy or with stable pre-existing conditions, or at risk of exposure (health and social care setting workers, 58.5%) | AZD1222 (AstraZeneca)<br>2 doses 4-12 wks apart<br>2.2-6.5x$10^{10}$ vp per 0.5ml dose, 2<br><br><br><br>Randomized: 12,408 | Meningococcal ACWY vaccine (UK)<br>OR<br>Meningococcal ACWY vaccine (dose 1) + saline placebo (dose 2; Brazil)<br>OR | Total randomized: 24,422<br><br>Total safety population: 24,244<br><br>Male 44%<br>Median age NR<br>18-55 y 82.5% | Solicited local and systemic AEs for 7 d after each injection observed by investigator or reported by participant; | Safety analysis included all participants who received at least one injection. | **Pericarditis:** 1 vaccine (0 grade 3 severity) vs. 2 (1 grade 3 severity) controls |

| Trial name;<br>Author (year);<br>Country (number of sites);<br>Dates of enrolment<br>Funding | Eligibility,<br>Randomization &<br>Blinding | Study Arms | | Baseline Demographics | Outcome(s);<br>Safety Assessment | Analysis | Results |
|---|---|---|---|---|---|---|---|
| | | Vaccine arm | Placebo arm | | | | |
| Brazil (6), South Africa (8), UK (5+19)<br><br>Industry-funded | Exclusions: confirmed or suspected immunosuppressive or immunodeficiency state; current diagnosis of cancer; pregnant or breastfeeding<br><br>Randomized 1:1, stratified by study site and study group; secure web platform<br><br>Single blind (Brazil, UK); double blind (South Africa) - study personnel and participants blinded, but pharmacist preparing vaccines unblinded | Safety population: 12,282 (after exclusion of 52 participants in HIV cohort)<br><br>Withdrawals NR | Saline placebo (South Africa)<br><br>For all: 2 doses 4-12 wks apart<br>0.5 ml dose<br><br>Randomized: 12014<br><br>Safety population: 11,962 (after exclusion of 52 participants in HIV cohort)<br><br>Withdrawals NR | 56-69 y 11.3%<br>70+ y 6.1%<br>White 75.2%<br>Cardiovascular disorder (includes cardiac diseases) 12.3%<br><br>Baseline SARS-CoV-2 status:<br>Positive: 731 (3.0%), similar between arms<br>Missing: 200 (0.8%), similar between arms<br><br>Sensitivity analyses for participants positive for SARS-CoV-2 at baseline and using ITT were similar to main results | **Unsolicited AEs** through to 28 d after each injection, and SAEs and AEs of special interest throughout study period from last dose to 364 d.<br><br>MedDRA v23.1 classification, severity classified according to FDA toxicity rating scales.<br><br>Independent data monitoring safety board reviewed safety data on an ongoing basis | | |

AE: adverse event(s); LTFU: lost to follow-up; MAAE: medically-attended adverse event(s); mcg: microgram(s); mo: month(s); no.: number; NR: not reported; SAE: serious adverse event(s); vp: viral particle(s); wks: weeks; y: year(s)

**Table A5. Risk of bias assessments for randomized controlled trials (question 1)**

| Vaccine (Trial name) | Randomization | Deviations from intended interventions | Missing outcome data | Measurement of the outcome | Selection of the reported result | Overall assessment of risk of bias |
|---|---|---|---|---|---|---|
| Moderna (Teen COVE Trial)<br><br>Ali 2021 | Low | Low | Some concerns | Low | Low | Some concerns |
| Moderna (COVE Trial)<br><br>Baden 2021 & El-Sahly 2021 | Low | Low | Low | Low | Low | Low |
| Pfizer-BioNTech (Teen C4591001 Trial)<br><br>Frenck 2021 | Low | Low | Low | Some concerns | Low | Some concerns |
| Pfizer-BioNTech (C4591001 Trial)<br><br>Polack 2021 & Thomas 2021 | Low | Low | Low | Some concerns | Low | Some concerns |
| AstraZeneca (AZD1222 Trial)<br><br>Falsey 2021 | Low | Some concerns | Some concerns | Low | Low | Some concerns |
| AstraZeneca (Oxford Vaccine Trial)<br><br>Voysey 2021a & 2021b | Low | Some concerns | Some concerns | Low | Low | Some concerns |
| Janssen (ENSEMBLE Trial)<br><br>Sadoff 2021 | Low | Low | Low | Low | Low | Low |

**Appendix 4. Summary of Findings from the Randomized Controlled Trials** (question 1)

| Age category | Age, risk status | Country (number of sites) | Vaccine (risk interval) | Number of participants in safety analysis | Number of participants with outcome | Conclusion | GRADE |
|---|---|---|---|---|---|---|---|
| | **Myocarditis** | | | | | | |
| 12-17 y | 12-17 y | USA (26)<br><br>Ali | Moderna (28 d) | 3,726 | 0 | RCTs provided very uncertain evidence about the incidence of myocarditis after receipt of Moderna in youth 12-17 years of age. | Very low [a] |
| Adolescents and young adults | 12-25 y | USA (29)<br><br>Frenck | Pfizer (21 d) | 3,357 | 0 (assumed, NR) | RCTs provided very uncertain evidence about the incidence of myocarditis after receipt of Pfizer in adolescents and young adults. | Very low [a] |
| Adolescents and adults | ≥16 y | Argentina (1), Brazil (2), Germany (6), South Africa (4), Turkey (9), USA (130)<br><br>Polack/Thomas | Pfizer (21 d) | 43,847 | 0 | RCTs provided very uncertain evidence about the incidence of myocarditis after receipt of Pfizer in adolescents and adults. | Very low [b] |
| Adults | ≥18 y, at risk of exposure to SARS-CoV-2 and/or increased risk for severe Covid-19 | USA (99)<br><br>Baden/El-Sahly | Moderna (28 d) | 30,351 | 0 | RCTs provided very uncertain evidence about the incidence of myocarditis after receipt of currently available vaccine by adults | Very low [b] |
| | ≥18 y, at high-risk of exposure to SARS-CoV-2 and increased risk for severe Covid-19 | Chile, Peru, USA (88 altogether)<br><br>Falsey | AstraZeneca (28 d) | 32,379 | 0 (assumed, NR) | | |
| | ≥18 y, 59% at risk of exposure to SARS-CoV-2 | Brazil (6), South Africa (8), UK (24)<br><br>Voysey | AstraZeneca (4-12 wk) | 24,244 | 0 (assumed, NR) | | |
| | ≥18 y | Argentina, Brazil, Chile, Colombia, Mexico, Peru, South Africa, USA (number of sites NR)<br><br>Sadoff | Janssen (single dose) | 43,783 | 0 (assumed, NR) | | |
| | **Pericarditis** | | | | | | |

| Age category | Age, risk status | Country (number of sites) | Vaccine (risk interval) | Number of participants in safety analysis | Number of participants with outcome | Conclusion | GRADE |
| --- | --- | --- | --- | --- | --- | --- | --- |
| 12-17 y | 12-17 y | USA (26)<br><br>Ali | Moderna (28 d) | 3,726 | 0 | RCTs provided very uncertain evidence about the incidence of pericarditis after receipt of Moderna in youth 12-17 years f age. | Very low [a] |
| Adolescents and young adults | 12-25 y | USA (29)<br><br>Frenck | Pfizer (21 d) | 3,357 | 0 (assumed, NR) | RCTs provided very uncertain evidence about the incidence of pericarditis after receipt of Pfizer in adolescents and young adults. | Very low [a] |
| Adolescents and adults | ≥16 y | Argentina (1), Brazil (2), Germany (6), South Africa (4), Turkey (9), USA (130)<br><br>Polack/Thomas | Pfizer (21 d) | 43,847 | 0 (assumed, NR) | RCTs provided very uncertain evidence about the incidence of pericarditis after receipt of Pfizer in adolescents and adults. | Very low [b] |
| Adults | ≥18 y, at risk of exposure to SARS-CoV-2 and/or increased risk for severe Covid-19 | USA (99)<br><br>Baden/El-Sahly | Moderna (28 d) | 30,351 | 2 (68 d & 73 d post-second dose) vs. 2 placebo | RCTs provided very uncertain evidence about the incidence of pericarditis after receipt of currently available vaccine by adults | Very low [b] |
| | ≥18 y, at high-risk of exposure to SARS-CoV-2 and increased risk for severe Covid-19 | Chile, Peru, USA (88 altogether)<br><br>Falsey | AstraZeneca (28 d) | 32,379 | 0 (assumed, NR) | | |
| | ≥18 y, 59% at risk of exposure to SARS-CoV-2 | Brazil (6), South Africa (8), UK (24)<br><br>Voysey | AstraZeneca (4-12 wk) | 24,244 | 1 vs. 2 controls | | |
| | ≥18 y | Argentina, Brazil, Chile, Colombia, Mexico, Peru, South Africa, USA (number of sites NR)<br><br>Sadoff | Janssen (single dose) | 43,783 | 1 (<0.1%) vs. 0 placebo | | |

GRADE Explanations:

[a] Rated down for indirectness since results by sex are expected to differ; rated down twice for high imprecision for these very rare events.

[b] Rated down twice for indirectness and for imprecision for these very rare events.
